# Identifying the best diagnostic test for Ovarian cancer in premenopausal women with non-specific symptoms – results from the ROCkeTS prospective, multicentre, cohort study

**DOI:** 10.1101/2025.10.17.25338220

**Authors:** Sudha Sundar, Ridhi Agarwal, Katie Scandrett, Clare Davenport, Ben Van Calster, Susanne Johnson, Partha Sengupta, Radhika Selvi-Vikram, Fong Lien Kwong, Sue Mallett, Caroline Rick, Sean Kehoe, Dirk Timmerman, Tom Bourne, Hilary Stobart, Richard D Neal, Usha Menon, Aleksandra Gentry-Maharaj, Lauren Sturdy, Ryan Ottridge, Jonathan J Deeks, ROCkeTS collaborators

**Affiliations:** PanBirmingham Gynaecological Cancer Centre, Sandwell and West Birmingham Hospitals NHS trust, Birmingham, UK; Department of Cancer and Genomic Sciences, University of Birmingham, Birmingham, UK; Department of Applied Health Science, University of Birmingham, Birmingham, UK; Department of Applied Health Science, Southampton University Hospitals, NHS trust; Durham and Darlington NHS trust; West Hertfordshire Hospitals NHS trust; Centre for Medical Imaging, University College London, London, UK; University of Nottingham; St Peter’s College, University of Oxford, Oxford, UK; Department of Development and Regeneration, KU Leuven, Leuven, Belgium; Department of Obstetrics and Gynecology, University Hospitals KU Leuven, Leuven, Belgium; Faculty of Medicine, Department of Metabolism, Digestion and Reproduction, Imperial College London, London, UK Patient Representative, Birmingham, UK; University of Exeter Medical School, University of Exeter, Exeter, UK; Department of Women’s Cancer, Elizabeth Garrett Anderson Institute for Women’s Health, University College London, London, UK; MRC Clinical Trials Unit, Institute of Clinical Trials and Methodology, University College London, London, UK; Birmingham Clinical Trials unit, University of Birmingham, UK; NIHR Birmingham Biomedical Research Centre, University Hospitals Birmingham NHS Foundation Trust and University of Birmingham, Birmingham, UK; Leuven Unit for Health Technology Assessment Research (LUHTAR), KU Leuven, Leuven, Belgium

## Abstract

**Objective:** Diagnosing ovarian cancer in premenopausal women is challenging due to the rarity of cancer and the ubiquity of symptoms, ovarian cysts on ultrasound, and raised serum CA125 tumour marker levels. We investigated the accuracy of risk prediction models and scores for diagnosing ovarian cancer in premenopausal women presenting to secondary care with symptoms and abnormal tests.

**Methods:** A cohort of premenopausal women presenting with non-specific symptoms, and raised CA125 or abnormal imaging, were prospectively recruited in 23 hospitals in the UK between June 2015 and March 2023, predominantly referred through the NHS urgent suspected cancer pathway from primary to secondary care. A head-to-head comparison of the accuracy of the six risk prediction models and scores was conducted using donated blood and ultrasound scans performed by NHS staff trained in the use of IOTA imaging terminology. Index tests (at pre-stated thresholds) were: RMI1 (200, 250); ROMA (7.4%, 11.4%, 12.5%, 13.1%); IOTA ADNEX (3%, 10%); IOTA SRRisk (3%, 10%); IOTA simple rules; and CA125 (87 IU/ml). Participants were classified as having primary invasive ovarian cancer versus having benign or normal pathology according to the reference standard determined from surgical specimens, biopsies or cytology, by histology if undertaken, or else by 12-month follow-up. After June 2018, because of COVID restrictions and concerns about sample size, ongoing recruitment was restricted to only women undergoing surgery within 3 months of presentation (a selected group in whom ovarian cancer was more likely).

**Results:** Of 1,211 premenopausal recruited women 88 were diagnosed with primary OC, 857 in the pre-June 2018 cohort (prevalence of 5.7% (49/857)) and 354 in the post-June 2018 cohort 11.0% (39/354). For the diagnosis of primary ovarian cancer, (n=799 women after exclusion of n=58 other diagnoses), RMI1 at the 250 threshold had a sensitivity of 42.6%, 95% confidence interval 28.3 to 57.8, and specificity of 96.5%, 94.7 to 97.8. Compared to RMI1/250, CA125 and all other models had higher sensitivity (CA125: 55.1%, 40.2 to 69.3, p=0.06; ROMA/11.4%: 79.2%, 65.0 to 89.5, p<0.0001; IOTA ADNEX/10%: 89.1%, 76.4 to 96.4, p<0.0001; IOTA SRRisk/10%: 83.0%, 69.2 to 92.4, p<0.0001; IOTA simple rules: 75.0%, 56.6 to 88.5, p=0.01) and lower specificity (CA125: 89.0%, 86.5 to 91.2, p<0.0001; ROMA/11.4%: 73.1%, 69.6 to 76.3, p<0.0001; IOTA ADNEX/10%: 75.1%, 71.4 to 78.6, p<0.0001; IOTA SRRisk/10%: 76.0%, 72.4 to 79.3, p<0.0001; IOTA simple rules 95.2%, 93.0 to 96.9, p=0.06). IOTA simple rules have inconclusive results in 120/799 of the participants. Analysis of the complete cohort (n=1,211) including the 354 premenopausal women with a higher likelihood of ovarian cancer, yielded similar results.

**Conclusions:** Compared to RMI 250, the current test used in NHS secondary care to triage women to tertiary care, most tests improve sensitivity but reduce specificity. Ultrasound triage with the IOTA ADNEX model at 10% in secondary care demonstrated the highest sensitivity gain with a comparable decline in specificity to other comparator tests. Ultrasound with the IOTA ADNEX model at 10% should be considered the new standard of care triage test for premenopausal women in secondary care; implementation in practice should incorporate staff training and quality assurance.

**Trial registration** – ROCkeTS is registered ISRCTN17160843

## Section 1: What is already known on this topic

- Diagnosing ovarian cancer in premenopausal women is challenging - OC is rare, while symptoms, elevated CA125, and physiological ovarian cysts are common. Current standard of care risk prediction model used to triage women with ovarian cysts into low or high risk of Ovarian cancer is the Risk of Malignancy index. Several alternate risk prediction models and scores show promise.
- We searched OVID MEDLINE, OVID EMBASE, and Cochrane Library (to 14 July 2025) using terms: ROMA, IOTA ADNEX, ORADS, IOTA simple rules, and RMI to identify optimal risk prediction models for OC in premenopausal women.
- Multiple studies were identified, but no head-to-head prospective comparisons of all tests exist. Studies were predominantly conducted in high-prevalence settings with expert ultrasound operators, limiting generalizability to non-specialist, primary care, or community settings.

## Section 2: What this study adds

- We conducted a prospective head-to-head test accuracy study of common risk prediction models in premenopausal women presenting to secondary care with symptoms, abnormal CA125, and abnormal ultrasound.
- Compared to published literature, our cohort was more representative of real-world populations with lower OC prevalence (5.7%). Ultrasound was performed primarily by NHS sonographers, enhancing applicability to practice.
- RMI at 250 shows poor sensitivity (42.6%) but high specificity (96.5%) in premenopausal women and requires replacement. Alternative tests improved sensitivity at the expense of reduced specificity compared to RMI 250.
- IOTA ADNEX at 10% threshold demonstrates the highest sensitivity (89%) with specificity (75%) comparable to other evaluated tests and is recommended for practice.

## Introduction

Ovarian cancer (OC) is a challenging disease to diagnose, with patients typically visiting General Practitioners (GPs) or primary care multiple times before testing is initiated.^1,2^ Currently, most women are diagnosed at advanced stages due to non-specific symptoms and suboptimal diagnostic pathways. Diagnosis is especially challenging in premenopausal women due to low OC prevalence (approximately 93,000 women diagnosed under 49 years globally, (age-standardized rate of 2.2-3.6/100,000^3^), ubiquitous symptoms, non-specifically elevated CA125, for example, during menses and physiological ovarian cysts on ultrasound. In England and Wales, 40% of women with OC are admitted as an emergency 4 weeks prior to diagnosis and are five times more likely to die within six months than women referred through urgent suspected cancer pathways. OC survival in the UK is significantly lower than in other western countries.

The National Institute for Health and Care Excellence (NICE) guidelines recommend sequential testing using serum CA125 and pelvic ultrasound for women presenting to their GP with symptoms such as persistent abdominal distension, feeling full, pelvic pain, increased urinary urgency, unexplained weight loss, fatigue, or changes in bowel habit.^4^ Women with elevated CA125 or abnormal ultrasound findings are referred to gynaecologists in secondary care hospitals through the urgent suspected cancer pathway in the UK NHS: patients referred to hospital receive a cancer/non-cancer diagnosis within 28 days of referral, and patients diagnosed with cancer receive first treatment within 62 days of referral.^5,6^

Poor performance of current diagnostic testing contributes to the challenge of timely, accurate diagnosis - CA125 identifies only 50% of early-stage OC and can be elevated in other benign conditions, while ultrasound in primary care lacks standardization or quality control and is associated with long waiting times.^7,8^ NICE guidance recommends the Risk of Malignancy Index (RMI) algorithm to triage women referred with suspected OC in hospital, combining age and menopausal status to generate a risk score.^4,9^ Women with an RMI score ≥250 are referred to tertiary care hospitals (gynaecological cancer centres) to be operated on by gynaecological cancer surgeons, while those with an RMI < 250 are managed in the referring secondary care hospital with surgery or surveillance by gynaecologists. The current pathway is summarised in Figure 1. Rates of surgery and additional imaging (MRI/CT) in referred women are high with the current pathway.^10^

Accurate triage is important - triaging a woman as high risk for OC preoperatively enables appropriate surgery at first attempt in a tertiary, specialist cancer centre, which improves survival.^4,11–13^ Improving diagnostic pathways for premenopausal women is a key unmet need. Alternatives (to RMI 250) risk prediction models endorsed variably by professional societies include the Risk of Malignancy Algorithm (ROMA) (combining He4 and CA125 biomarkers, stratified by menopausal status), and several ultrasound based models(the Ovarian-Adnexal Reporting and Data system (ORADS); the Assessment of Different NEoplasias in the adneXa (ADNEX); and the International Ovarian Tumour Analysis (IOTA) IOTA Simple rules).^14–20^

A 2022 Cochrane systematic review highlighted the absence of high-quality, head-to-head comparative test accuracy studies applicable to a primary care referred population with included studies conducted in high-prevalence settings (16-27%).^21^ The Cochrane review also demonstrated variation in performance of risk prediction models between pre- and postmenopausal women, reflecting differences in disease prevalence and spectrum.^21^ Additionally, the trade-offs between identifying true positives (enabling early diagnosis and better survival) versus limiting false positives (reducing unnecessary referral, anxiety and surgery) are different for pre compared to postmenopausal women, where fertility preservation and ovarian function are critical considerations.

The ROCkeTS study aimed to improve on existing evidence by identifying the best diagnostic test for women referred to secondary care hospitals with symptoms and abnormal CA125 and/ or ultrasound. We evaluated the accuracy of alternative risk prediction models: IOTA simple rules, IOTA SRRisk model, ROMA, ORADS, IOTA ADNEX, compared to the RMI 250 standard of care triage test in the UK (Box 1 and Supplementary Appendix C) against a reference standard of histology or follow-up for referral to tertiary care for surgery/biopsy. Results for postmenopausal women have been previously reported.^22^ This manuscript presents results for premenopausal women.

## Methods

Our report adheres to the STARD and TRIPOD checklists (Supplementary Tables 10-11).^23,24^ The trial protocol is on https://www.birmingham.ac.uk/research/bctu/trials/pd/rockets and has previously been published ^25^ Methods for ROCkeTS has been previously described (we reproduce this briefly below) and in the statistical analysis plan (Supplementary Appendix D). ROCkeTS received ethical approval from NHS West Midlands REC (14/WM/1241) and is registered on the controlled trials website (ISRCTN17160843). All participants provided informed consent prior to participation in the study.

## Participants

This prospective cohort study consecutively recruited newly presenting women aged 16-90 years who were referred to hospitals within the UK. Research nurses from the UK national cancer collaborative research network screened patients attending hospital clinics referred from primary care through urgent suspected cancer pathways, ultrasound clinics, gynaecology clinics or presenting through emergency admissions. To be eligible, women needed to have symptoms consistent with NICE guidance for OC (including but not restricted to persistent or frequent abdominal distension, feeling full (early satiety) and/or loss of appetite, pelvic or abdominal pain, increased urinary urgency and/or frequency^4^) as well as elevated CA125 levels or abnormal ultrasound findings. At recruitment, participants self-reported sex and ethnicity and completed a structured questionnaire to obtain medical and gynaecological history. Exclusion criteria comprised pregnancy, existing non-ovarian malignancy, and previous ovarian malignancy.

Participants were categorised as pre- or postmenopausal status at recruitment by their age (<50, 51+ years) and whether they had periods in the last 12 months. Post recruitment, perimenopausal women were re-categorised based on a self-reported history of vaginal bleeding to allow analysis with existing risk prediction models that incorporate different thresholds or covariates based on menopausal status.

The study protocol underwent significant evolution from initial publication.^25^ The ROCkeTS premenopausal study initially recruited consecutive eligible women who were scheduled for surgical intervention within 3 months of referral to hospital or who were being managed through conservative approaches, including surveillance or discharge as clinically indicated, with planned questionnaire-based assessment of wellbeing at 12 months.

At an interim analysis, OC prevalence was observed to be 3%, sufficient for estimating specificity but inadequate to estimate comparisons of sensitivity between tests and models. Consequently, adaptations to the protocol were introduced in March 2018, excluding women with simple ovarian cysts <5cm and normal CA125 levels due to their extremely low cancer risk, and from June 2018 recruitment focused solely on newly presenting premenopausal women scheduled for surgery within 3 months. From June 2018, IOTA ultrasound scan was deemed optional, acknowledging both the scheduling challenges for women undergoing surgery for suspected cancer and ethical concerns about requiring additional hospital visits during the COVID-19 pandemic (2020-2022).

The study cohort comprised two groups: the pre-protocol change cohort (labelled Cohort 1) recruited up until June 2018, which includes both pre-surgical and conservatively managed patients, and the post-protocol change cohort, post June 2018, which comprises only pre-surgical patients (labelled Cohort 2). A sensitivity analysis was undertaken to account for potential changes in spectrum, with the pre-protocol change cohort regarded as most likely to be representative and used in the primary analysis.

Three patients were deemed ineligible after recruitment – one due to a previously unknown pregnancy, one due to being diagnosed with non-ovarian malignancy prior to recruitment and one due to planned surgery not going ahead. No screen eligible and willing patients were excluded. The current NHS standard of care triage test, RMI 250, was used to manage patients.

### Index tests

All patients completed a symptom questionnaire, donated a blood sample and underwent transabdominal and a transvaginal USS scan. Serum samples were collected, processed, and stored according to predefined standard operating procedures detailed in a laboratory manual distributed to participating sites, consistent with consensus guidance from the Early Detection Research Network. (Appendix D). ^26^ Samples were transported and stored at -80°C until analysis at NHS South Tyne and Wear Pathology Services laboratories. For consistent analysis, samples were thawed in batches and tested for CA125 and HE4 using Roche Cobas e802 modules. HE4 and CA125 measurements employed electrochemiluminescence immunoassay (ECLIA) technology, adhering to manufacturer recommendations. Roche Elecsys assay kits were obtained from Roche Diagnostics.

The following index tests were evaluated (Box 1, Supplementary Appendix C):

- ROMA, (a combination of CA125 and He4 tumour markers) at the manufacturer recommended threshold of 11.4% for premenopausal women and at previously reported thresholds of: 7.4%, 12.5%, 13.1%.^27^
- The Risk of Malignancy Index 1 (RMI 1), (a combination of CA125 and limited ultrasound features), at thresholds of 200 and 250. The 250 threshold was chosen as the comparator test, representing the current standard of care diagnostic approach for triaging patients to gynaecological cancer centres within the UK National Health Service.^4,17^
- Serum CA125 measurement at a threshold of 87 IU/ml was selected based on its association with a positive predictive value of 3% in primary care as detailed by Funston et al.^28^
- USS based tests: Three tests developed by the IOTA consortium were evaluated, two models IOTA ADNEX and the IOTA simple rules risk (SRRISK) model at a primary thresholds of 10% and secondary thresholds of 3% and one classifier IOTA simple rules.^16,19,29–31^. IOTA ADNEX is now considered a medical device and is manufactured by Gynaia.^32^
- ORADS was evaluated in a post-hoc analysis using IOTA variables from the ROCkeTS ultrasound case report form retrospectively mapped to the ORADS lexicon 1-3 versus 4-5 using previously described methodological approaches^19,33^ for this analysis.

We define IOTA ultrasound scan as one where IOTA terminology has been used to describe the ultrasound findings.^34^ IOTA terminology is a set of terms, definitions and measurements used to precisely describe ultrasound features seen in adnexal masses.^34^ Ultrasound examinations in ROCkeTS were conducted by sonographers who underwent comprehensive training in IOTA terminology, including one-day in-person and online instruction, followed by formal examination. A mandatory quality assessment process was implemented, with a sample of ultrasound images and reports centrally reviewed by the IOTA team of ultrasound experts has been described previously.^22^ No prior minimum ultrasound experience requirement was stipulated. Scans were primarily performed by level II (non-medical) sonographers which mirrors real-world clinical practice, where pelvic ultrasounds are performed by sonographers with varying levels of experience.

All diagnostic tests and subsequent surgical interventions or biopsies were required to be completed within three months of patient recruitment. All tests were conducted blinded to the reference standard.

## Reference standard

For Cohort 1, the reference standard was histology of surgical specimens, biopsies or cytology or 12-month surveillance for women who did not undergo surgery. In Cohort 2, study participation ended at surgery/biopsy/cytology, which served as the reference standard.

Pathology data were sourced from specialist gynecological pathology reports from the 40 cancer centres in the UK where women undergoing surgery for suspected OC are discussed at specialist gynaecological oncology multidisciplinary team meetings.

For participants under surveillance, 12-month well-being was ascertained through both patient self-completed postal questionnaires and research nurse-completed questionnaires utilizing hospital records from a clinic visit or by contacting patients by telephone. No diagnosis of cancer or histology results were based on self-report alone. Both information sources were cross-referenced to identify any cancer diagnoses occurring within 12 months of study recruitment.

Reference standard results were not available to sonographers or the research team prior to trial entry, although clinical information was available as this is considered usual clinical practice.

### Outcomes

The primary outcome focused on the diagnostic accuracy of index tests for identifying OC. This was defined as a binary outcome: primary invasive malignant neoplasms diagnosed through surgical or biopsy histology, versus benign, normal, or surveillance findings. The definition of primary invasive OC included ovarian, fallopian tube, and primary peritoneal cancers.

The secondary outcome included a broader spectrum of malignancies encompassing primary invasive cancers, secondary malignant neoplasms metastatic to the ovary, borderline neoplasms, and neoplasms of uncertain or unknown behaviour diagnosed through surgery, biopsy, or cytology, versus benign, normal on follow-up findings. Analysis of the secondary outcome was also performed with borderline tumours grouped with benign/ normal follow-up findings rather than with malignancies.

### Statistical analysis

Clinical study data was entered in structured case report forms onto an electronic study management platform hosted by Medscinet, https://rockets.medscinet.com/. Data was entered by research nurses at sites. Data cleaning and addressing data queries was performed by the Birmingham Clinical Trials unit with trial statisticians. The statistical analysis plan is included in the supplementary materials. Diagnostic accuracy was assessed using sensitivity, specificity and the positive and negative predictive values (PPV and NPV). Risk prediction tools were dichotomised at different thresholds. The difference in sensitivity and specificity (and their corresponding 95% confidence intervals) comparing tests was assessed using the exact McNemar’s test with asymptotic confidence intervals. Multiple testing was accounted for by use of the Bonferroni correction (11 pairwise comparisons, p=0.0045). (21, 22). No single measure of accuracy was designated as primary endpoint a priori. This approach was chosen to fully evaluate the trade-offs inherent in performance of diagnostic tests.

Global accuracy performance was further assessed in terms of discrimination, using a c-index and receiver operating characteristic (ROC) plot, and calibration, using calibration plots and calibration slope. We used the ‘pmcalplot’ command in Stata to generate the calibration plots.^35^

Where index tests produced inconclusive results (IOTA Simple rules), we opted not to classify inconclusive results as positive, as this could have led to an overestimation of test performance for sensitivity estimates.

Women with missing index test data were excluded from both primary and secondary analyses. Participants with missing or inconclusive reference standard results were excluded from the primary outcome definition of presence or absence of OC but included in the secondary outcome to make the best use of participant data.

A sensitivity analysis was conducted for both the primary and secondary definitions of OC where missing index test results were imputed using the multiple imputation by chained equations (MICE) for predictors of index test combinations by replacing missing values with plausible values based on the distribution of the observed data.^36^ Multiple imputation was performed using the ‘mi’ package in Stata 17.^37^

Initial analysis was performed for primary and secondary outcomes in the pre-protocol change cohort, in a combined cohort grouping Cohort 1 and Cohort 2 together.

### Sample size

The original sample size was based on local audit data (unpublished) to estimate the performance of RMI in premenopausal women, as previously published systematic reviews did not provide separate estimates for pre- and postmenopausal women. Based on the performance of RMI having a sensitivity of 72% and specificity of 46%, the study was designed to detect increases in sensitivity of 10% and in specificity of 10%, assuming a prevalence of OC in premenopausal women referred to secondary care of 10%.^38^ A sample size of 1000 would provide 100 OC events in which to build new models combining symptom and test data (adequate events to model 10 predictor variables) and will provide 90% power to detect an increase in specificity of 8% (from 46% for RMI to 54%). With a predicted loss to follow-up of up to 5%, the final sample size required is 1050 women.^39^

In an interim analysis in 2018, a much lower prevalence (3%) in premenopausal women was observed than previously assumed (10%). Therefore, both sample size and inclusion criteria were adapted to ensure that the study had adequate power to estimate the difference in sensitivities of the index tests by recruiting 105 women (accounting for 5% dropout) identified as having OC. Following the change of the inclusion criteria, whilst a prevalence of 14.9% was expected in women who undergo surgery, the revised sample size of 880 patients was based on the assumption that only 11.9% (80%) actually undergo surgery.

## Results

### Characteristics of study population

The ROCkETS study recruited 2,268 eligible pre- and postmenopausal women referred to 23 UK hospitals between June 30, 2015 and March 23, 2023, and followed up to March 31, 2023 (Figure 2). Results of the 1,057 post-menopausal cohort have been reported separately ^22^. Here, we report results for the cohort of 1,211 premenopausal participants, comprising 857 women recruited up to June 2018 under the initial protocol (Cohort 1), and 354 women recruited under the adapted protocol restricted to women scheduled for surgery within 3 months (Cohort 2). The majority of the participants in Cohort 1 were recruited from primary to secondary care through the urgent suspected cancer pathway (574/857; 67%). The main analysis focuses on the results of Cohort 1, as it represents ‘real-world’ practice and has low missingness of IOTA USS. Findings of Cohort 2 are included in a sensitivity analysis.

Recruited women in the first cohort had a median age of 44.1 (IQR 35.0-48.7) years. The majority were of white ethnicity (n=732, 85.4%), had never smoked (n=476, 55.5%), and were not using contraception (n=502, 58.6%) (Table 1). Close to over-half of the women (n=444, 51.8%) had comorbidities: 137 (16.0%) with fibroids, 135 (15.8%) with endometriosis and 121 (14.1%) with irritable bowel syndrome. 161 (18.8%) had undergone previous surgery: 44 (5.1%) hysterectomy, 82 (9.6%) cystectomy, 33 (3.9%) salpingectomy and 25 (2.9%) oophorectomy; and 217 (25.3%) reported a family history of cancer: 42 (4.9%) ovarian, 98 (11.4%) breast, 48 (5.6%) colon and 16 (1.9%) uterus (Table 1). We did not observe any systematic differences in patient characteristics between Cohort 1 and Cohort 2.

### Prevalence of ovarian cancer

Eighty-eight of the 1211 women (7.3%) were diagnosed with the primary outcome of ovarian cancer: 47 (53.4%) at FIGO Stage I, 6 (6.9%) at Stage II, 24 (27.2%) at Stage III and one (1.1%) at Stage IV (stage was missing for 10). Five of the cases were diagnosed during the 12-months follow-up, 83 by surgery or biopsy histology. Seventy-three cases had epithelial histology types (n=73, 88.0%) (Table 2).

In the 857 women in Cohort 1, 49 (5.7%) of were diagnosed with OC and 58 (6.8%) were reported as ‘other’ and were excluded from estimation of accuracy for the primary outcome (13 had a missing primary outcome, 22 had borderline neoplasm, 7 had no histology, 10 had secondary malignant neoplasm, 1 had a diagnostic category of ‘Other’ and 5 reported a diagnosis of non-OC identified at 12-months follow up).

### Accuracy of index tests

Of the 799 women included in the analysis for the primary OC classification in Cohort 1, 581 (72.7%) had complete data for all index tests. Results were available for CA125 for 795 (99.5%), ROMA 750 (93.9%), RMI 1 for 672 (84.1%), IOTA SRRisk model 668 (83.6%), IOTA ADNEX 617 (77.2%) (Table 3, Supplementary Table 1). For the IOTA simple rules, results were only available for 553 (69.2%), as 126 had missing and 120 had inconclusive results.

The pattern of values of sensitivity and specificity across the tests showed a threshold effect, with tests with the lowest sensitivity having the highest specificity, and *vice versa* (Table 3). RMI 1 at thresholds of 200 and 250 had the lowest sensitivities of 48.9% (95% CI: 34.1 to 63.9) and 42.6% (95% CI: 28.3 to 57.8), respectively. Compared to RMI 1 at 250, CA125 and all other models had higher sensitivity (CA125 55.1%,, 95% CI 40.2 to 69.3, p=0.06; IOTA simple rules: 75.0%, 95% CI 56.6 to 88.5, p=0.01; ROMA/11.4%: 79.2%, 95% CI 65.0 to 89.5, p<0.0001; IOTA SRRisk/10%: 83.0%, 95% CI 69.2 to 92.4, p<0.0001; IOTA ADNEX/10%: 89.1%, 95% CI 76.4 to 96.4, p<0.0001). RMI 1 at thresholds of 200 and 250 had the highest specificities of 95.4% (95% CI: 93.4 to 96.9) and 96.5% (95% CI: 94.7 to 97.8), respectively. Compared to RMI 1 at 250, all other tests had lower specificity: IOTA simple rules 95.2%, 93.0 to 96.9, p=0.06; CA125/87U/ml: 89.0%, 86.5 to 91.2, p<0.0001; IOTA SRRisk/10%: 76.0%, 72.4 to 79.3, p<0.0001; IOTA ADNEX/10%: 75.1%, 71.4 to 78.6, p<0.0001; ROMA/11.4%: 73.1%, 69.6 to 76.3, p<0.0001).

IOTA ADNEX demonstrated the highest global accuracy with a C-index (area under the curve, AUC) of 0.89 (95% CI: 0.83 to 0.96), followed by IOTA SRRisk 0.86 (0.80 to 0.93); RMI 1 0.85 (0.79 to 0.91); IOTA Simple rules 0.85 (0.77 to 0.93); ROMA 0.84 (0.77 to 0.92) and CA125 0.80 (0.72 to 0.87) (Table 3, Figure 3A). The calibration of the models for ROMA, IOTA ADNEX and IOTA SRRisk model demonstrated under-prediction (Figure 3B).

All index tests had high negative predictive values, ranging from 95.7% (95% CI: 93.8 to 97.2) for RMI1 at 250 to 98.9% (95% CI: 96.7 to 99.8) for IOTA ADNEX at 3%, driven by the low prevalence. Positive predictive values were from 10.4% (95% CI: 7.6 to 13.7) for ROMA at 7.4% to 49.0% (95% CI: 34.4 to 63.7) for IOTA simple rules (Table 3).

### Secondary outcome – presence of any cancer

In Cohort 1, 86/857 (10.0%) women were diagnosed with the secondary outcome definition of the presence of any cancer (Supplementary Table 2). Estimates of sensitivity were lower than for the primary outcome, with sensitivity values being lower by 4% for RMI 1 at 250 and 14% for IOTA ADNEX. RMI 1 of 250 reported the lowest sensitivity of 39.0% (95%: 28.4 to 50.4)), whilst IOTA ADNEX reported the highest sensitivity of 75.3% (95%: 64.5 to 84.2). Specificity values for the secondary outcome were very similar to those for the primary outcome.

RMI 1, ROMA, IOTA ADNEX and IOTA sRisk reported very similar global accuracy C-index values of 0.81, whilst values were lower for IOTA simple rules (0.76) and CA125 (0.73) (Supplementary Table 2, Supplementary Figure 1A). The ROMA model demonstrated good calibration, whilst the IOTA ADNEX and IOTA sRisk probabilities were underpredictions (Supplementary Figure 1B).

For the secondary outcome, negative predictive values ranged from 91.8% (RMI 1 at a threshold of 250) to 97.9% (ROMA at a threshold of 7.4%), lower than for the primary outcome. Positive predictive values ranged from 17.0% (ROMA at a threshold of 7.4%) to 56.9% (IOTA simple rules).

### Sensitivity analyses

For the primary outcome, the accuracy estimates were comparable in sensitivity analyses that combined participants from the Combined Cohorts, with only small changes in point estimates but no changes in the ranking of performance (Supplementary Table 3, Supplementary Figures 2A and 2B), and in analyses using imputation for missing data. (Supplementary Table 4).

Sensitivity analyses of the secondary outcome showed consistent results from the Combined Cohorts (Supplementary Table 5; Supplementary Figures 3A and 3B) and in analyses using imputation for missing data. (Supplementary Table 6).

We also analysed the diagnostic accuracy of the index tests, including borderline tumours with benign tumours as normal in the Combined Cohorts analysis, findings were consistent with main results (Supplementary Table 7).

Results according to quality assurance passed sonographers or high-volume recruiting centres were also consistent with findings of the main primary and secondary analyses (data not submitted but available on request).

### Post-hoc analysis of ORADs

In post-hoc analyses, in a Combined Cohorts analysis we compared the primary outcome (Supplementary Table 8) and secondary outcome (Supplementary Table 9) for ORADS at a 10% threshold with RMI 1 at 250. ORADS demonstrated better sensitivity than RMI 1 250 but with lower specificity for both outcomes. For the primary outcome, ORADS 4 had a sensitivity of 81.3% (95% CI: 69.5 to 89.9) and a specificity of 82.2% (95% CI: 79.5 to 84.6). Similarly for the secondary outcome ORADS had a sensitivity of 68.9% (95% CI: 60.3 to 76.7) with a specificity of 82.2% (95% CI: 79.5 to 84.6).

## Discussion

### Statement of principal findings

The cancer prevalence in pre-menopausal women observed in ROCkeTS is low (5.7% in the pre-protocol Cohort 1, 7% in Combined Cohorts). Only 1% of women referred less than 40 years old for urgent suspected cancer are diagnosed with OC. ^40^ Achieving a high sensitivity (minimising false negative (missed) diagnoses) is therefore important and challenging. Conversely, reducing unnecessary investigations and surgery due to false positive diagnoses (maintaining specificity) is also important. We have previously demonstrated a high level of anxiety in women referred through urgent suspected cancer pathways, which persists at 12 months post-referral, despite a non-cancer diagnosis.^40^ The desire for fertility and ovarian function is also likely to be valuable to this group: 90 women (7.4%) were pursuing fertility, and only 345 (33.0%) were using contraception.

The ROCkeTS study investigated diagnostic tests for OC in premenopausal women with symptoms and abnormal CA125, ultrasound or both. The majority of women were referred from primary care through the urgent suspected cancer pathway to hospital clinics. Results showed that the current UK standard triage test, RMI 1 at a threshold of 250, has a poor sensitivity of 42.6% (95% CI: 28.3 to 57.8), despite a good specificity of 96.5% (95% CI: 94.7 to 97.8). IOTA ADNEX at thresholds of 10% and 3%, ROMA at 11.4% and 7.4%, and IOTA SR Risk models at 10% and 3% significantly improve on RMI 1’s sensitivity but with a significant fall in specificity. Compared to RMI 1, IOTA ADNEX at 10% achieved the highest sensitivity at 89.1% (95% CI: 76.4 to 96.4) with a relatively limited loss of specificity, 75.1% (95% CI: 71.4 to 78.6).

These results were consistent for the detection of primary OC, metastatic cancers, and borderline tumours combined (secondary outcome analysis), and across sensitivity analyses.

Amongst comparator tests, IOTA Simple Rules appeared to offer an improvement in sensitivity compared to RMI 1 to 75.0% (95% CI 56.6 to 88.5) whilst maintaining a high specificity of 95.2% (95% CI 93.0 to 96.9). However, the test classified 120/799 women (15.0%) as inconclusive. Inconclusive results are seldom random; they typically represent hard-to-diagnose, borderline presentations with a higher underlying risk of malignancy.

Excluding them introduces spectrum and missing-not-at-random biases, making sensitivity and specificity appear better than would be observed in practice.

Following the study, we were able to undertake a post-hoc analysis of ORADS at a 10% threshold, showing a sensitivity of 81.3% (95% CI 69.5 to 89.9) and a specificity of 82.2% (95% CI 79.5 to 84.6%), with a lower gain in sensitivity than IOTA ADNEX but higher specificity. Further investigation of the performance of ORADS in a prospective study is needed.

### Comparison with existing literature

A search on 14^th^ July 2025 showed no new prospective head-to-head comparative tests accuracy studies evaluating all relevant models for the diagnosis of OC. Our 2022 Cochrane systematic review of risk prediction models for the diagnosis of OC demonstrated a similar pattern of results in premenopausal women of a higher sensitivity but lower specificity of ROMA (27 studies, 4463 participants) and IOTA ADNEX (4 studies, 1696 participants) compared to RMI 1 (17 studies, 5233 participants).^21^ Differences in accuracy estimates can be explained by the highly selected participants in included studies (mean prevalence OC 16-27%) compared to a prevalence of 5.7% for the ROCkeTS main analysis. Results of ROCkeTS in premenopausal women are also consistent with our previous report from postmenopausal women where we identified IOTA ADNEX ultrasound as the triage test achieving the highest gain in sensitivity over RMI triage with a reduction of specificity comparable to other comparator tests.^22^

### Implications for practice for clinicians and policymakers

Results need to be interpreted in the context of current practice. ROCkeTS recruited predominantly from urgent suspected cancer pathway referrals (67%), and 60% or primary OC diagnoses were stage I/II (pelvis confined) when the cancer is likely curable with standard of care treatment.^41^ A risk prediction model with high sensitivity would improve survival, ensuring women with OC are triaged appropriately to receive surgery from trained specialist gynaecological cancer surgeons whilst enabling women identified as ‘low risk’ to be managed without surgery and with reassurance alone.^13^

Data from ROCkeTS demonstrates high surgery rates in women triaged using RMI 1 (64.7%, 551/857 patients) despite its relatively high specificity, at least in part because current guidance advocates surgery even in pre-menopausal women triaged as ‘low risk’ due to the known low sensitivity of RMI in this group. Balancing gains in sensitivity to detect OC at an early stage against reductions in specificity, and given the limitations of the IOTA simple rules, we recommend IOTA ADNEX at 10% to replace RMI1 at 250 as the UK standard of care OC triage test in secondary care. The sensitivity and specificity of the IOTA ADNEX ultrasound model was achieved using non-specialist, appropriately trained, certified and quality assured sonographers and IOTA ultrasound training resources have been established through ROCkeTS and are available for all NHS staff.^42^

In ROCkeTS, to robustly evaluate algorithm performance, we evaluated IOTA ADNEX model performance in all patients. In practice, a two-step strategy, initially triaging out women with a benign appearances on scan (< 1% risk of cancer over 2 years) and then using IOTA ADNEX to calculate risk of OC in the remaining women has been shown to improve specificity. ^43–45^ For patients with IOTA ADNEX scores of 10-50%, additional MRI imaging can help prevent unnecessary surgery. We have previously discussed strategies needed to successfully implement IOTA ADNEX ultrasound and a flowchart for practice (Supplementary Figure 4) ^46^ Our results also suggest that earlier, accurate OC diagnosis is possible using IOTA ADNEX ultrasound as a first test in primary care, potentially concurrent with CA125 testing and this approach needs evaluating in further research (Supplementary Figure 5)

### Strengths

To our knowledge, ROCkeTS is the first multi-site, blinded, prospective head-to-head comparison study of all commonly used candidate risk prediction models for the diagnosis of OC in newly presenting premenopausal women. Selection bias was minimized by recruiting through the UK National cancer collaborative research network infrastructure. Quality assured index tests were compared against a common reference standard and a predefined statistical analysis plan included appropriate handling of missing data. Differentiation between primary (OC) and secondary (all cancer types including borderline) outcomes allowed investigation of test performance without inflation by inclusion of borderline tumours, which although common in younger women, have little impact on survival. The study mirrored real life and practice where some patients who are referred undergo surgery and others are kept under surveillance or discharged. Inclusion of 12-month follow-up allows us to investigate false negatives of risk prediction models that would be used in clinical care to make triage decisions in women with suspected OC.

In contrast to previous studies the low prevalence of OC in ROCkeTS is applicable to a primary care referred population^21^ and most scans were performed by level II, non-medical NHS sonographers rather than clinically qualified experts (gynaecologists or radiologists) participating from centers of excellence.^47^

### Limitations

Although ROCkets recruitment is consistent with UK census patterns, (mainly white ethnicity), results may be less applicable to more diverse ethnicities. Although we are unable to identify differences in patient characteristics across pre- and post-protocol change cohorts, our decision to restrict recruitment to pre-surgical patients following observation of low OC prevalence in an interim analysis is likely to have led to systematic differences between the two cohorts. Further, not scheduling an additional hospital visit for a transvaginal IOTA ultrasound post-protocol change to reduce risk during COVID and to improve recruitment led to high missingness, particularly of ultrasound variable data, in the cohort of women recruited post-protocol change.

Although we encouraged sites to obtain the IOTA ultrasound scan at the same time as routine scans for clinical care; we did not collect information on how this was delivered across sites. It is possible that the sonographers who completed the scan to calculate RMI were different in some sites from the sonographers who collected the IOTA ultrasound scan.

Despite these issues, the consistency of results across pre-protocol, combined cohort, and imputed analyses across primary and secondary analyses demonstrates the robustness of the study’s findings. The challenges of conducting ROCkeTS and estimating sample size are common to evaluations of test accuracy in low prevalence populations – we believe that our study has learnings for others designing a diagnostic test accuracy study to diagnose rare significant events amongst commonly occurring backgrounds.

### Implications for research

The performance of risk prediction models in ethnically diverse populations needs further research. Alternative methods of evaluating the impact of diagnostic test use on clinical utility or net benefit may be highly relevant and will be conducted as next steps in the ROCkeTS study.^48^ Exploring the effect of threshold on test performance using the area under the curve may play a role in determining optimal trade-offs in sensitivity and specificity. However, limitations of this approach includes not providing the threshold needed to realise the optimal trade-off. This is essential for clinicians and health systems to make decisions about further management. The use of decision curve analysis to compare decisions at the same thresholds across risk prediction models was not possible because RMI 1 does not produce probabilities.

Long waiting times for ultrasound, absence of standardisation and quality assurance are key challenges to achieving IOTA USS implementation at scale in the NHS.^7^ Artificial Intelligence (AI) enabled solutions for ultrasound along with quality assurance and training for sonographers may deliver timely ultrasound availability in practice for women with non-specific symptoms and significantly improve outcomes by early diagnosis. The impact of AI enabled USS with IOTA ADNEX in primary care practice on expediting diagnostic intervals and improving early detection of OC needs investigation. Understanding facilitators and barriers to implementing change in diagnostic pathways, including a one-stop model for IOTA USS, is currently being investigated. (SONATA study, (NCT06129968).

Compared to RMI 1 at 250, in post-hoc analysis, ORADS at 10% achieved a smaller gain in sensitivity but a lower drop in specificity than IOTA ADNEX at 10%. However, the latest version of ORADS v2 with additional variables was not evaluated.^30^ Prospective multicentre research studies investigating the diagnostic accuracy of ORADS for OC is necessary.

## Conclusion

The current triage test for OC, RMI 1, demonstrates poor sensitivity in premenopausal women and should be replaced. IOTA ADNEX at 10% delivered by trained and quality-assured NHS sonographers achieves significantly higher sensitivity with limited specificity reduction in a real-world cohort and should be considered the new standard of care for secondary care triage. Primary care implementation could potentially improve survival through earlier detection in symptomatic premenopausal women (Supplementary Figure 5); this requires further research alongside investment in sonographer training and quality assurance.

## Dissemination plans

These results have been presented at the International gynaecological cancer society meeting in Dublin, October 2024 and at multiple national meetings. Results will be summarised in lay language and placed on the ROCkeTS trial website. We do not plan to contact participants individually to communicate the results of the study

## Contributor and guarantor information

SS, CD, SM, JD conceptualised and designed the study. SS, SJ, PS, RS-V recruited to the study with collaborators with CR, RO and LS coordinating the study, SM, JD, KS and RA analysed results from the study, DT and TB conducted ultrasound QA, training, BVC provided insight into analysis of ultrasound models, SK, RN, UM and A G-M provided input into study design and conduct. HS provided patient perspectives throughout the study from the grant application, study conduct and interpretation of results. All authors reviewed the results and the manuscript.

JD, RA and KS along with LS and RO have directly accessed and verified the underlying data reported in the manuscript. All authors confirm that they had full access to all the data in the study and accept responsibility to submit for publication

SS is guarantor of paper. The guarantor accepts full responsibility for the work and/or the conduct of the study, had access to the data, and controlled the decision to publish. “The corresponding author attests that all listed authors meet authorship criteria and that no others meeting the criteria have been omitted.”

## Copyright/license for publication

The Corresponding Author has the right to grant on behalf of all authors and does grant on behalf of all authors, a worldwide licence to the Publishers and its licensees in perpetuity, in all forms, formats and media (whether known now or created in the future), to i) publish, reproduce, distribute, display and store the Contribution, ii) translate the Contribution into other languages, create adaptations, reprints, include within collections and create summaries, extracts and/or, abstracts of the Contribution, iii) create any other derivative work(s) based on the Contribution, iv) to exploit all subsidiary rights in the Contribution, v) the inclusion of electronic links from the Contribution to third party material where-ever it may be located; and, vi) licence any third party to do any or all of the above.”

## Patient consent (if applicable) – not applicable

### Competing interests declaration

SS reports a research grant from AoA diagnostics for work with samples collected in this study but not reported within this manuscript. SS reports honoraria from Astra Zeneca, Mercke and GSK and consultancy from GSK and Immunogen, all unrelated to this work. TBo reports grants, personal fees, and travel support from Samsung Medison; travel support from Roche Diagnostics; and personal fees from GE Healthcare; all outside the submitted work. BVC and DT report consultancy work done by KU Leuven to help implementing and testing the IOTA ADNEX model in ultrasound machines by Samsung Medison and GE Healthcare, outside the submitted work. Profs Timmerman and Bourne declare that they are cofounders of a KU Leuven spinout company, Gynaia incorporated in May 2025 (i.e post submission of ROCkeTS manuscript in Dec 2024).

UM stock ownership awarded by University College London (UCL) until October 2021 in Abcodia. UM and AGM report research collaboration contracts with QIMR Berghofer Medical Research Institute, iLOF (intelligent Lab on Fiber), RNA Guardian, Micronoma, Mercy Bioanalytics, Synteny Biotechnology. UM reports research support grants paid to the institution from CleoDx related to early detection of cancer especially ovarian cancer. UM declares membership of the Research Advisory Panel, Yorkshire Cancer Research (UK) and honorarium for membership of Tina’s Wish Scientific Advisory Board (USA). UM holds patent number EP10178345.4 for Breast Cancer Diagnostics.

SK reports honorary role as Ovacome charity trustee. DT, TBo and BVC are IOTA steering group members and developed the IOTA models.

All other authors declare no competing interests.

## Data availability

The dataset generated including deidentified patient data and samples analysed during the study, along with additional material such as protocol, statistical analysis plan is available at Birmingham Clinical Trials Unit, University of Birmingham after date of publication. The dataset is not publicly available but maybe obtained on request to SS, review by Project oversight group, NIHR, ethics approval and after fulfilling all data transfer requirements

## Patient and Public Involvement

The study was supported by a patient co-applicant who played a crucial role throughout the research process. HS provided regular input at key stages of the study, beginning with the application for funding, study design, recruitment and continuing through to the dissemination of findings. Patient advocates from Target Ovarian cancer were involved as partners with the research team to shape the study’s design, develop and review informational materials for the study, and assess the burden of questionnaires from the patient’s perspective.

At the conclusion of the study, HS and Target Ovarian cancer charity contributed feedback on the findings and provided valuable input into the interpretation of results and outcome measures, ensuring that the findings were meaningful from a patient-centred perspective.

They also contributed to the dissemination plan with an online presentation to lay audiences, review of press release of previously published manuscripts from the ROCkeTS study.

### Transparency declaration

The lead author (the manuscript’s guarantor) affirms that the manuscript is an honest, accurate, and transparent account of the study being reported; that no important aspects of the study have been omitted; and that any discrepancies from the study as originally planned (and, if relevant, registered) have been explained.

## Supporting information

Main tables

## Data Availability

The dataset generated including deidentified patient data and samples analysed during the study, along with additional material such as protocol, statistical analysis plan is available at Birmingham Clinical Trials Unit, University of Birmingham after date of publication. The dataset is not publicly available but maybe obtained on request to SS, review by Project oversight group, NIHR, ethics approval and after fulfilling all data transfer requirements. All data produced in the present study are available upon reasonable request to the authors

