## Supplementary material for "Identifying the best diagnostic test for Ovarian cancer in premenopausal women with non-specific symptoms – results from the ROCkeTS prospective, multicentre, cohort study": Main tables

**Rockets premenopausal paper – Figures and Tables for manuscript**

Box 1 – Risk prediction models and scores evaluated in ROCkeTS

Figure 1 – Flowchart describing referral pathways for patients presenting with non-specific symptoms to primary care

Figure 2 – Recruitment of patients in ROCkeTS study

Figure 3A – ROC curve of the index test combinations for the primary outcome in the pre-protocol change cohort

Figure 3B – Calibration plots for ROMA (left), IOTA sRisk (middle) and ADNEX (right) of the primary outcome in Cohort 1.

Table 1 – Participant demographics and clinical characteristics

Table 2 – Histopathology of pelvic masses in women who underwent surgery and/or biopsy (n=837) or identified by follow-up (n=5) in the Combined Cohort

Table 3 – Diagnostic performance statistics of index test combinations for the primary outcome in Cohort 1

**Figure 1: Flowchart describing referral pathways for patients presenting with non-specific symptoms to primary care**

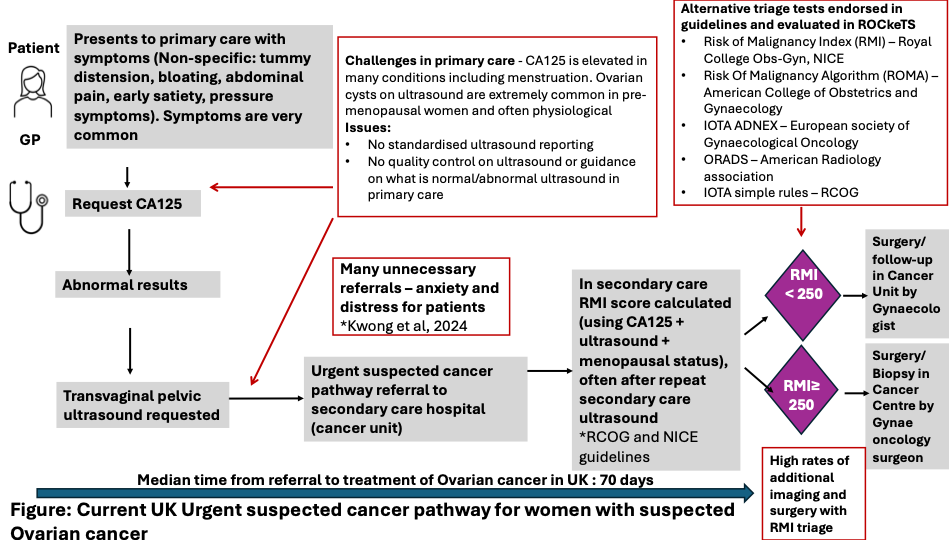

| **Algorithms** | **Explanation** |
| --- | --- |
| **Risk of Malignancy algorithm** | This combines menopausal status, CA125 tumour marker levels and specific ultrasound features to provide a numerical score. Commonly used cut-offs are 200 and 250 to guide management. |
| **Risk of Malignancy Algorithm** | This combines serum levels of tumour markers HE4 and CA125 in an algorithm to provide a numerical score. Thresholds vary by menopausal status and by manufacturer. |
| **IOTA simple rules** | Ultrasound based - This is a classifier based on 5 benign and 5 malignant ultrasound features. Results are expressed as Benign, Malignant or Inconclusive |
| **IOTA ADNEX** | Risk prediction model combining three clinical variables and six ultrasound variables. The model expresses risk in percentage for borderline, stage I cancer, stage II-IV cancer, and secondary metastatic cancer. 3% and 10% are commonly used thresholds. |
| **IOTA SRRisk Model** | A logistic regression model that uses the 10 ultrasound features in IOTA simple rules plus the type of center (oncology center vs non-oncology center) as predictors to calculate the likelihood of malignancy in adnexal masses. |
| **CA125** | Serum CA125 in units U/ml tumour marker level |
| **ORADS** | O-RADS is a clinical support system that provides a standardized lexicon for describing ovarian and adnexal lesions and stratifies them using a numeric score based on morphologic features to indicate risk of malignancy. 0-3 vs 4-5 is equivalent to 10% risk of malignancy |

**Box 1. Risk prediction models and scores evaluated in ROCkeTS study**

**Figure 2: Recruitment of patients in the ROCkeTS study**

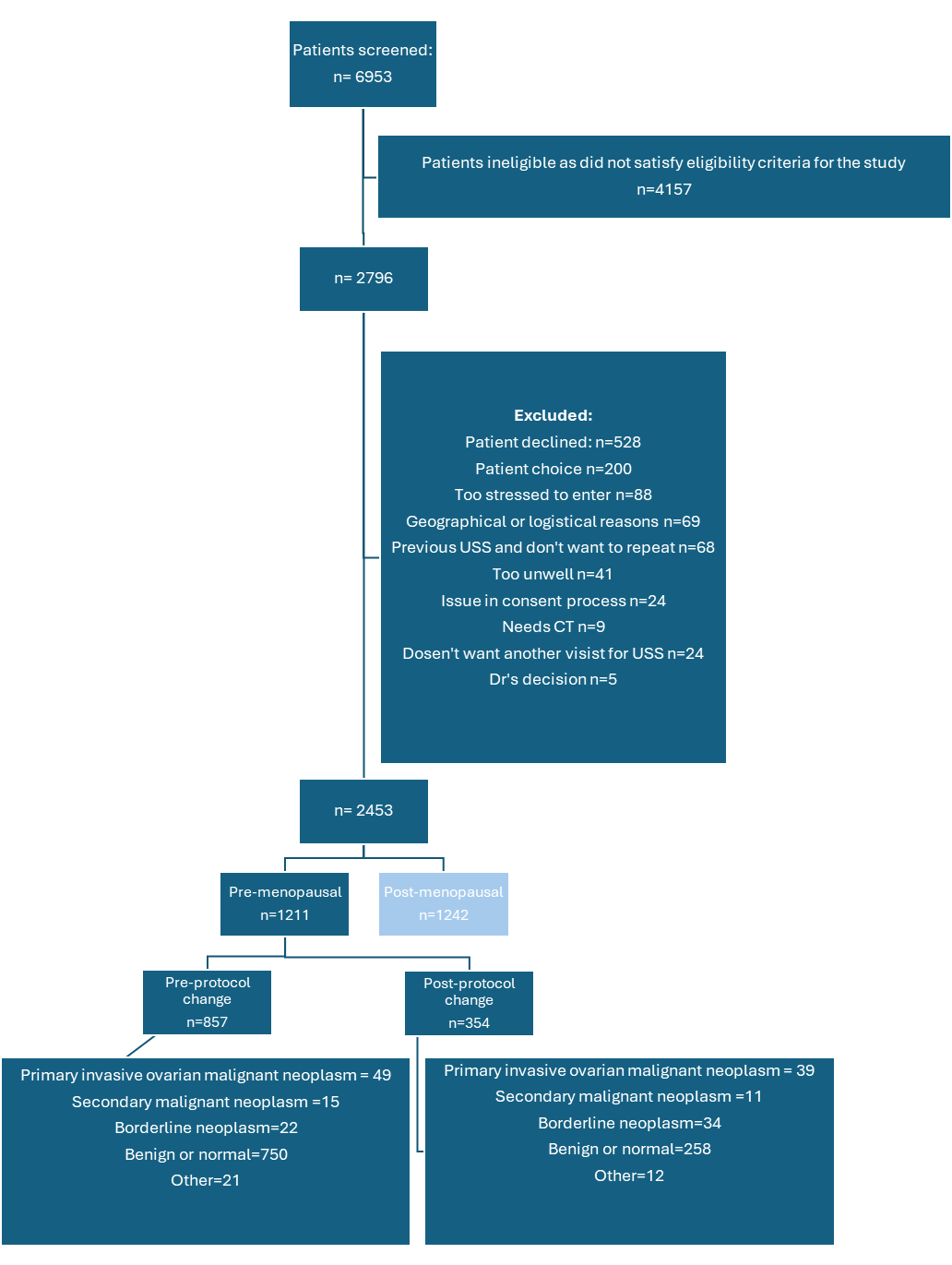

**Table 1. Participant demographics and clinical characteristics**

|  | **Cohort** **1**  n=857 | **Cohort** **2**  n=354 | **Total**  n=1,211 |
| --- | --- | --- | --- |
|  | Data *are n (%) or median (IQR)* | | |
| **Demographics** |  |  |  |
| Age (years) | 44.1 (35.0-48.7) | 40.2 (31.1-47.1) | 43.2 (33.7-48.3) |
| Height (cm) | 164.0 (159.0-168.0) | 164 (160.0-169.0) | 164 (160-169.0) |
| Weight (kg) | 71.6 (62.0-85.4) | 76 (64.0-90.0) | 73 (62.0-86.0) |
| Ethnicity, n (%) |  |  |  |
| White | 732 (85.4%) | 294 (83.1%) | 1,026 (84.7%) |
| Mixed | 18 (2.1%) | 9 (2.5%) | 27 (2.2%) |
| Asian | 51 (5.9%) | 22 (6.4%) | 73 (6.0%) |
| Black | 20 (2.3%) | 10 (2.8%) | 30 (2.5%) |
| Other | 10 (1.2%) | 4 (1.1%) | 14 (1.2%) |
| Never smoked | 476 (55.5%) | 211 (59.6%) | 687 (56.7%) |
| Alcohol (units per week) | 2 (0-9) | 2 (0-6) | 2 (0-8) |
| **Current conditions *** |  |  |  |
| None | 413 (48.2%) | 187 (52.8%) | 600 (49.6%) |
| Fibroids | 137 (16.0%) | 47 (13.3%) | 184 (15.2%) |
| Endometriosis | 135 (15.8%) | 34 (9.6%) | 169 (14.0%) |
| Irritable bowel syndrome | 121 (14.1%) | 35 (9.9%) | 156 (12.9%) |
| Hypertension | 66 (7.7%) | 26 (7.3%) | 92 (7.6%) |
| Arthritis | 59 (6.9%) | 22 (6.2%) | 81 (6.7%) |
| Adhesions | 34 (4.0%) | 11 (3.1%) | 45 (3.7%) |
| Diabetes | 22 (2.6%) | 10 (2.8%) | 32 (2.6%) |
| Uterine polyps | 25 (2.9%) | 4 (1.1%) | 29 (2.4%) |
| Uterine or bladder prolapse | 20 (2.3%) | 7 (2.0%) | 27 (2.2%) |
| High cholesterol | 21 (2.5%) | 11 (3.1%) | 32 (2.6%) |
| **Medical history** |  |  |  |
| Surgical history, n (%) |  |  |  |
| Hysterectomy | 44 (5.1%) | 8 (2.3%) | 52 (4.3%) |
| Ovarian cystectomy | 82 (9.6%) | 34 (9.6%) | 116 (9.6%) |
| Salpingectomy | 33 (3.9%) | 13 (3.7%) | 46 (3.8%) |
| Oophorectomy | 25 (2.9%) | 6 (1.7%) | 31 (2.6%) |
| Previous diagnosis of cancer |  |  |  |
| Breast | 15 (1.8%) | 3 (0.9%) | 18 (1.5%) |
| Other | 20 (2.3%) | 7 (2.0%) | 27 (2.2%) |
| Family cancer history |  |  |  |
| Ovary | 42 (4.9%) | 14 (4.0%) | 56 (4.6%) |
| Breast | 98 (11.4%) | 38 (10.7%) | 136 (11.2%) |
| Colon | 48 (5.6%) | 27 (7.6%) | 75 (6.2%) |
| Uterus | 16 (1.9%) | 9 (2.5%) | 25 (2.1%) |
| Stated sexually active | 604 (70.5%) | 238 (67.2%) | 842 (69.5%) |
| Pregnancies | 2 (0-3) | 1 (0-3) | 1 (0-3) |
| Live births** | 2 (1-2) | 2 (1-3) | 2 (1-3) |
| Vaginal deliveries** | 2 (1-2) | 1 (0-2) | 1 (1-2) |
| Caesarean sections** | 0 (0-1) | 0 (0-1) | 0 (0-1) |
| Trying to get pregnant | 70 (8.2%) | 20 (5.7%) | 90 (7.4%) |
| History of subfertility | 116 (13.5%) | 45 (12.7%) | 161 (13.3%) |
| History of ovarian stimulation | 49 (5.7%) | 19 (5.4%) | 68 (5.6%) |
| Painful periods | 471 (55.0%) | 205 (57.9%) | 676 (55.8%) |
| Change in the nature of periods | 409 (47.7%) | 170 (48.0%) | 579 (47.8%) |
| Change in period pain in last year | 350 (40.8%) | 138 (39.0%) | 488 (40.3%) |
| Irregular periods in last year | 389 (45.4%) | 152 (42.9%) | 541 (44.7%) |
| Heavy periods in last year | 412 (48.1%) | 172 (48.6%) | 584 (48.2%) |
| Currently using contraception | 226 (31.0%) | 117 (37.0%) | 345 (33.0%) |

Cohort 1 are participants recruited up until June 2018, which includes both pre-surgical and conservatively managed patients. Cohort 2 are participants recruited post June 2018 following the study protocol adaptation.

Data completeness is 100% for age, current conditions and medical history; between 96-97% for other demographics, sexually active and gravidity; between 86-87% for gynaecological history.

* Current conditions with less than 20 cases (<2%) were reported in women with adenomyosis, diverticulitis, sexually transmitted infection, epilepsy, heart disease, pelvic Inflammatory disease, vulva pain/vulvodynia and jaundice.

** Live births, vaginal deliveries and Caesarian Sections were counted in participants with one or more pregnancies.

**Table 2. Histopathology of pelvic masses in women who underwent surgery and/or biopsy (n=837) or identified by follow-up (n=5) in the Combined Cohort**

| **Histology** |  | | **n (%)** | |
| --- | --- | --- | --- | --- |
| **Primary invasive malignant neoplasm** | | |  | **88 (10.5%)** |
| Surgery and/or biopsy | Malignant epithelial | *Clear cell* | 7 |  |
|  |  | *Endometrioid* | 14 |  |
|  |  | *High grade serous* | 20 |  |
|  |  | *Low grade serous* | 9 |  |
|  |  | *Mucinous* | 21 |  |
|  |  | *Undifferentiated* | 1 |  |
|  |  | *Not available* | 1 |  |
|  | Germ cell | *Immature teratoma* | 1 |  |
|  |  | *Yolk Sac Tumour* | 1 |  |
|  | Sex cord | *Granulosa cell tumour* | 5 |  |
|  |  | *Not reported* | 1 |  |
|  | Subtype not available |  | 2 |  |
| Follow-up | Not reported* |  | 5 |  |
| **Secondary malignant neoplasm** | | |  | **21 (2.5%)** |
|  |  | *Endometrium* | 11 |  |
|  |  | *Appendix* | 5 |  |
|  |  | *Colon* | 2 |  |
|  |  | *Stomach* | 2 |  |
|  |  | *Breast* | 1 |  |
| **Borderline** | | |  | **58 (6.9%)** |
|  |  | *Mucinous* | 24 |  |
|  |  | *Serous* | 14 |  |
|  |  | *Seromucinous* | 1 |  |
|  |  | *Serous and mucinous* | 1 |  |
|  |  | *Not available* | 18 |  |
| **Benign** | | |  | **601 (71.4%)** |
|  | Benign tumours | *Serous adenofibroma* | 4 |  |
|  |  | *Serous cystadenoma and fibroma* | 99 |  |
|  |  | *Mucinous adenofibroma* | 2 |  |
|  |  | *Mucinous cystadenomas* | 109 |  |
|  |  | *Mature teratoma* | 116 |  |
|  |  | *Brenner tumour* | 2 |  |
|  |  | *Endometrioma* | 142 |  |
|  |  | *Fibroma/fibrothecoma* | 8 |  |
|  |  | *Functional* | 37 |  |
|  | Other pathology | *Endometrioid adenofibroma* | 8 |  |
|  |  | *Uterine fibroid* | 29 |  |
|  |  | *Chronic inflammation/abscess* | 13 |  |
|  |  | *Hydrosalpinx* | 7 |  |
|  |  | *Struma ovarii* | 6 |  |
|  |  | *Seromucinous cystadenoma* | 3 |  |
|  |  | *Mature teratoma & mucinous cystadenoma* | 1 |  |
|  |  | *Mucinous cystadenoma & Brenner tumour* | 2 |  |
|  |  | *Not available* | 13 |  |
| **Normal** |  |  | **-** | **47 (5.6%)** |
| **No histology** |  |  | **-** | **27 (3.2%)** |
| **Total** |  |  | **-** | **842** |

*For 5 patients, histology results were classified as having ovarian cancer based on free text information but did not have structured histology details recorded. For these women, a diagnostic category of “Other” was selected, with free-text entries describing malignancy consistent with ovarian cancer (e.g., “high grade serous carcinoma CK7, WT1, CA125, ER positive, CK20 negative”; “Poorly differentiated cancer – signet ring”; and “The features within the left ovary are of a serous papillary”). However, no additional structured fields were completed to confirm the final histological diagnosis.

**Table 3. Diagnostic performance statistics of index test combinations for the primary outcome in Cohort 1**

| Index test  combination | Threshold | Diagnosis based on  reference standard, n=799 | | Number of  Participants, n (%) | Sensitivity (%)  (95% CI) | Specificity (%)  (95% CI) | C-index  (AUC)  (95% CI) | Positive  predictive value (PPV)  (%)  (95% CI) | Negative predictive value (NPV)  (%)  (95% CI) | Pairwise comparison with RMI 1 (250)  (95% CI), p-value, number of participants |
| --- | --- | --- | --- | --- | --- | --- | --- | --- | --- | --- |
|  |  | OC  n=49 | No OC  n=750 |  |  |  |  |  |  |  |
| RMI 1,  n (%) | Missing | 2 (4.1) | 125 (16.7) |  | | | | | | |
|  | >200 | 23 (46.9) | 29 (3.9) | 672 (84.1) | 48.9  (34.1, 63.9) | 95.4  (93.4, 96.9) | 0.853  (0.792, 0.914) | 44.2  (30.5, 58.7) | 96.1  (94.3, 97.5) | Se: -6.4 (-15.5, 2.7), p=0.2500  Sp: 1.1 (0.1, 2.1), p=0.0156, n=672 |
|  | <200 | 24 (49.0) | 596 (79.5) |  |  |  |  |  |  |  |
|  | >250 | 20 (40.8) | 22 (2.9) |  | 42.6  (28.3, 57.8) | 96.5  (94.7, 97.8) |  | 47.6  (32.0, 63.6) | 95.7  (93.8, 97.2) | * |
|  | <250 | 27 (55.1) | 603 (80.4) |  |  |  |  |  |  |  |
| ROMA,  n (%) | Missing | 1 (2.0) | 48 (6.4) |  | | | | | | |
|  | >7.4% | 43 (87.8) | 372 (49.6) | 750 (93.9) | 89.6  (77.3, 96.5) | 47.0  (43.3, 50.8) | 0.844  (0.769, 0.918) | 10.4  (7.6, 13.7) | 98.5  (96.6, 99.5) | Se: -47.8 (-64.4, -31.2), p<0.0001  Sp: 49.3 (45.0, 53.7), p<0.0001, n=636 |
|  | <7.4% | 5 (10.2) | 330 (44.0) |  |  |  |  |  |  |  |
|  | >11.4% | 38 (77.6) | 189 (25.2) |  | 79.2  (65.0, 89.5) | 73.1  (69.6, 76.3) |  | 16.7  (12.1, 22.2) | 98.1  (96.5, 99.1) | Se: -37.0 (-53.1, -20.8), p<0.0001  Sp: 23.6 (19.6, 27.5), p<0.0001, n=636 |
|  | <11.4% | 10 (20.4) | 513 (68.4) |  |  |  |  |  |  |  |
|  | >12.5% | 37 (75.5) | 164 (21.9) |  | 77.1  (62.7, 88.0) | 68.6  (64.4, 72.5) |  | 18.4  (13.3, 24.5) | 97.0  (94.7, 98.5) | Se: -34.8 (-50.7, -18.8), p<0.0001  Sp: 19.8 (16.0, 23.6), p<0.0001, n=636 |
|  | <12.5% | 11 (22.4) | 358 (47.7) |  |  |  |  |  |  |  |
|  | >13.1% | 36 (73.5) | 149 (19.9) |  | 75.0  (60.4, 86.4) | 78.8  (75.6, 81.7) |  | 19.5  (14.0, 25.9) | 97.9  (96.3, 98.9) | Se: -32.6 (-48.3, -16.9), p=0.0001  Sp: 17.6 (13.9, 21.3), p<0.0001, n=636 |
|  | <13.1% | 12 (24.5) | 553 (73.7) |  |  |  |  |  |  |  |
| ADNEX,  n (%) | Missing | 3 (6.1) | 179 (23.9) |  | | | | | | |
|  | >3.0% | 43 (87.8) | 311 (41.5) | 617 (77.2) | 93.5  (82.1, 98.6) | 45.5  (41.4, 49.7) | 0.891  (0.827, 0.955) | 12.1  (8.9, 16.0) | 98.9  (96.7, 99.8) | Se: -52.2 (-68.8, -35.6), <0.0001  Sp: 50.8 (46.5, 55.1), p<0.0001, n=617 |
|  | <3.0% (Secondary) | 3 (6.1) | 260 (34.7) |  |  |  |  |  |  |  |
|  | >10.0% | 41 (83.7) | 142 (18.9) |  | 89.1  (76.4, 96.4) | 75.1  (71.4, 78.6) |  | 22.4  (16.6, 29.1) | 98.8  (97.3, 99.6) | Se: -47.8 (-64.4, -31.2), p<0.0001  Sp: 21.2 (17.4, 25.0), p<0.0001, n=617 |
|  | <10.0% (Primary) | 5 (10.2) | 429 (57.2) |  |  |  |  |  |  |  |
| IOTA  sRRisk  model,  n (%) | Missing | 2 (4.1) | 129 (17.2) |  | | | | | | |
|  | >3.0% | 41 (83.7) | 231 (30.8) | 668 (83.6) | 87.2  (74.3, 95.2) | 62.8  (58.9, 66.6) | 0.863  (0.799, 0.928) | 15.1  (11.0, 19.9) | 98.5  (96.7, 99.4) | Se: -44.7 (-61.0, -28.3), p<0.0001  Sp: 33.6 (29.5, 37.7), p<0.0001, n=666 |
|  | <3.0% (Secondary) | 6 (12.2) | 390 (52.0) |  |  |  |  |  |  |  |
|  | >10.0% | 39 (79.6) | 149 (19.9) |  | 83.0  (69.2, 92.4) | 76.0  (72.4, 79.3) |  | 20.7  (15.2, 27.2) | 98.3  (96.7, 99.3) | Se: -40.4 (-57.8, -23.1), p<0.0001  Sp: 20.4 (16.6, 24.1), p<0.0001, n=666 |
|  | <10.0% (Primary) | 8 (16.3) | 472 (62.9) |  |  |  |  |  |  |  |
| IOTA  simple  rules,  n (%) | Missing | 2 (4.1) | 124 (16.5) |  | | | | | | |
|  | Malignant | 24 (49.0) | 25 (3.3) | 553 (69.2) | 75.0  (56.6, 88.5) | 95.2  (93.0, 96.9) | 0.851  (0.774, 0.928) | 49.0  (34.4, 63.7) | 98.4  (96.9, 99.3) | Se: -28.1 (-49.1, -7.2), p=0.0117  Sp: 2.3 (-0.1, 4.7), p=0.0576, n=551 |
|  | Benign | 8 (16.3) | 496 (66.1) |  |  |  |  |  |  |  |
|  | Inconclusive | 15 (30.6) | 105 (14.0) |  | | | | | | |
| CA125,  n (%) | Missing | 0 (0.0) | 4 (0.5) |  | | | | | | |
|  | >87 U/ml | 27 (55.1) | 82 (10.9) | 795 (99.5) | 55.1  (40.2, 69.3) | 89.0  (86.5, 91.2) | 0.797  (0.723, 0.872) | 24.8  (17.0, 34.0) | 96.8  (95.2, 98.0) | Se: -10.6 (-21.6, 0.3), p=0.0625  Sp: 6.9 (4.6, 9.2), p<0.0001, n=672 |
|  | <87 U/ml | 22 (44.9) | 664 (88.5) |  |  |  |  |  |  |  |

OC=ovarian cancer; CI=confidence interval; AUC=area under the curve; Se=Sensitivity, Sp=Specificity, CI=confidence interval;

OC present includes primary invasive ovarian malignant neoplasm or diagnosed with ovarian cancer in the last 12 month; OC absent includes benign, normal or absent with ovarian cancer in the last 12 months. Note that 58 (6.8%) had a diagnostic category of other and were not included in the analysis.

**Figure 3A: Receiver operating characteristic (ROC) plot of the index test combinations for the primary outcome in Cohort 1.**

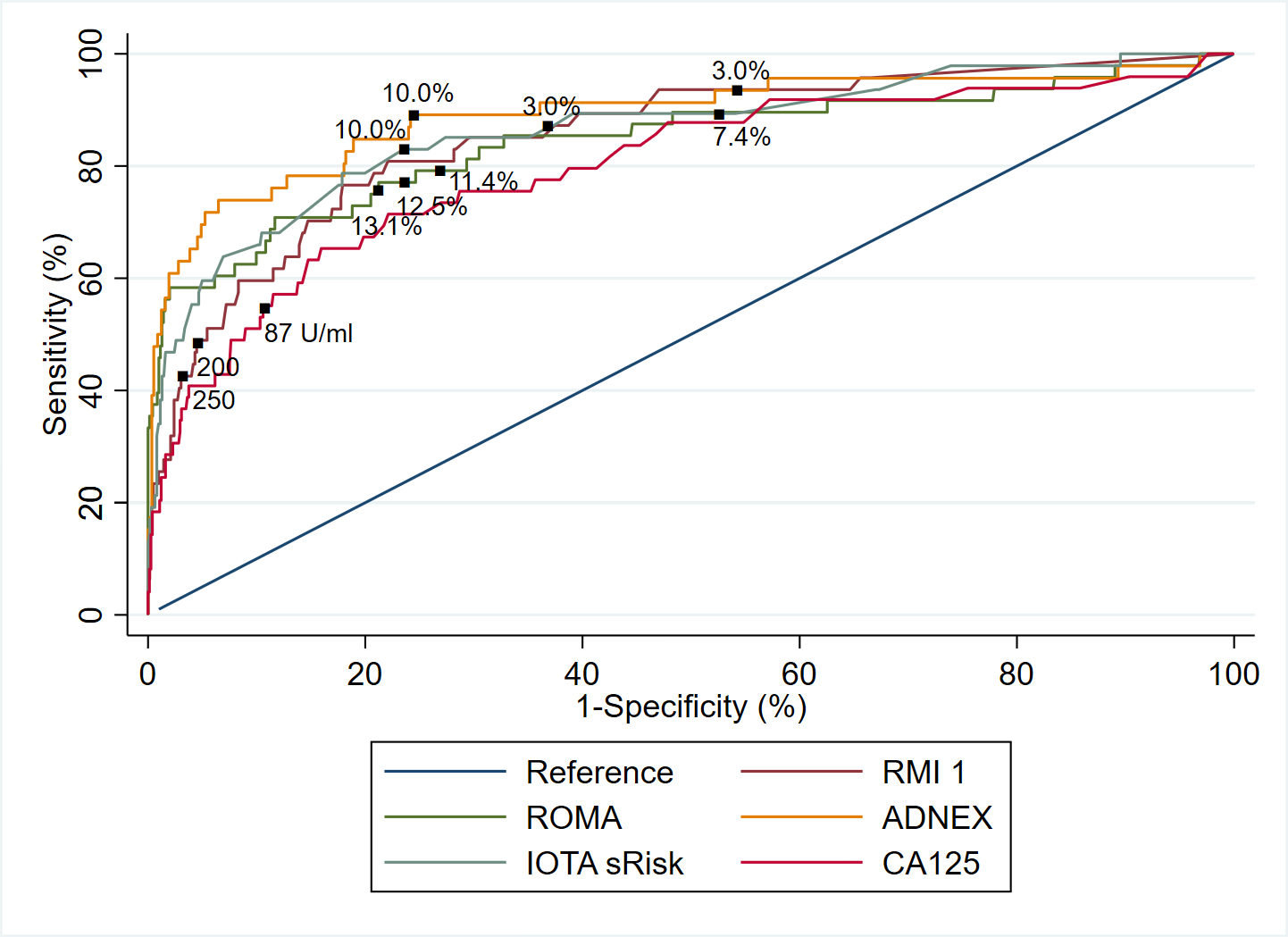

Note that the plot was produced by considering participants with available data for each index test combination separately, such that the number of participants used for each ROC curve varies. IOTA simple rules not included in the ROC plot because some participants received inconclusive results.

**Figure 3B: Calibration plots for ROMA (left), IOTA sRisk (middle) and ADNEX (right) of the primary outcome in Cohort 1.**

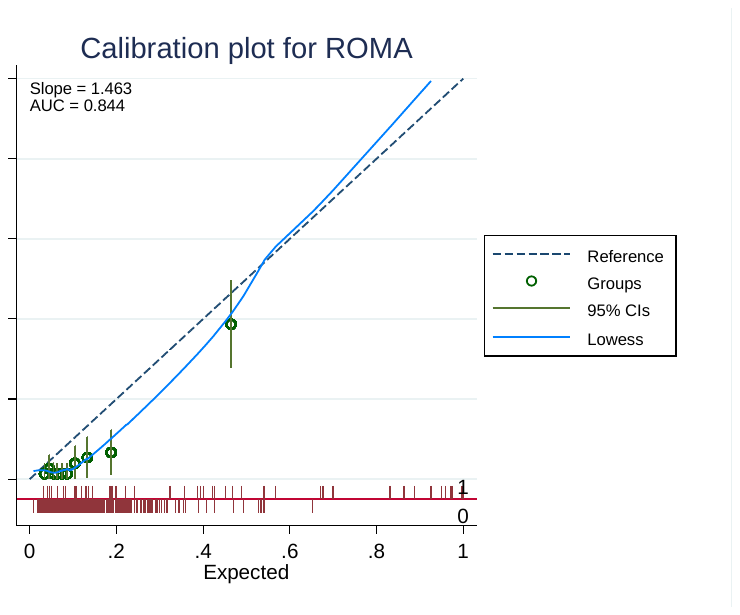

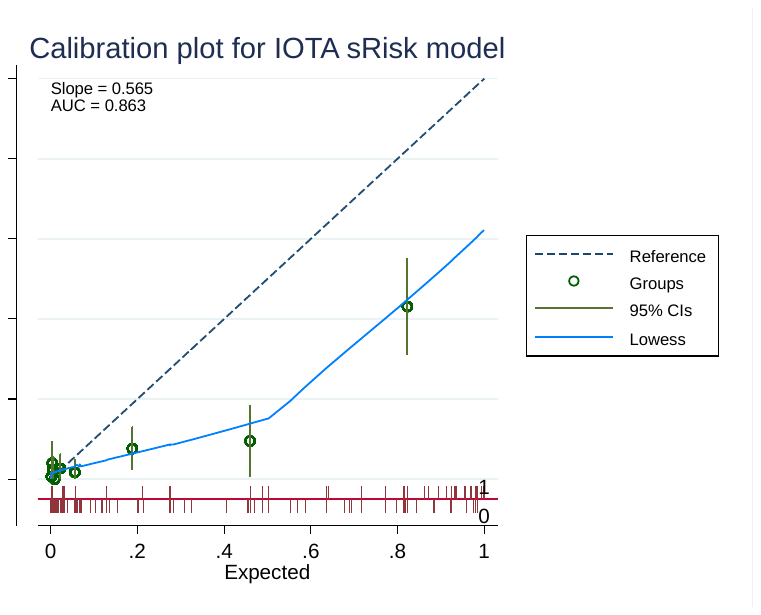

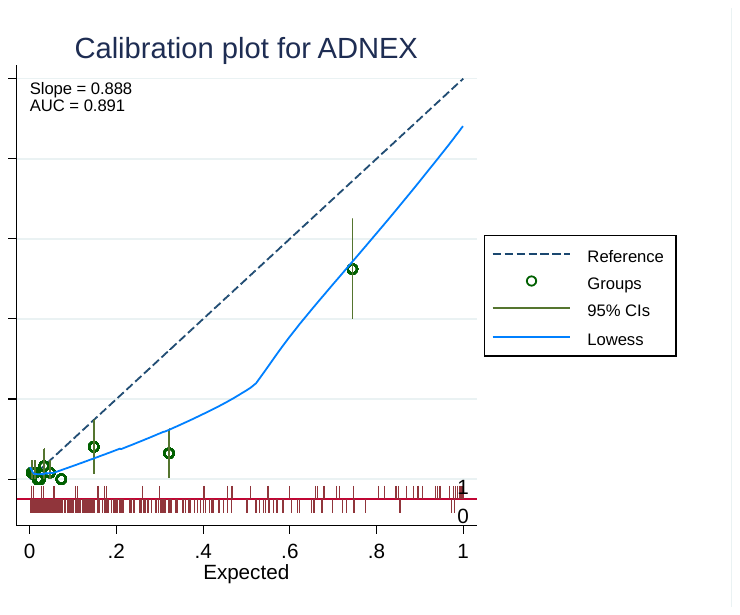

Calibration slope values (95% CIs) (derived from logistic regression model where the outcome is regressed on the predicted log-odds) were ROMA 1.46 (1.11, 1.82), IOTA sRisk 0.89 (0.69, 1.09) and IOTA ADNEX 0.56 (0.43, 0.70).

Note that the calibration slope and plot were only obtained for index test combinations that use a prediction model.
