## Supplementary tables for "Identifying the best diagnostic test for Ovarian cancer in premenopausal women with non-specific symptoms – results from the ROCkeTS prospective, multicentre, cohort study"

Supplementary Material

[APPENDIX B – Reporting Guidelines	2](#_Toc977552262)6

[APPENDIX C – Algorithms for Index Test Combinations and Thresholds](#_Toc635294413)	30

APPENDIX D – ROCKeTs Laboratory Manual	36

### APPENDIX A – Additional Data

**Supplementary Table 1.** Completeness of predictors used in the index test combinations for the Combined Cohorts

| **Component** | **Predictors**  ***Unit or result*** | | **Cohort 1**  n=857 | **Cohort 2**  n=354 | **Total**  n=1,211 | **Total missing** |
| --- | --- | --- | --- | --- | --- | --- |
| Patient  Baseline CRF | Age, y*ears* | Median (IQR) | 44.1  (35.0-48.7) | 40.2  (31.1-47.1) | 43.2  (33.7-48.3) | 0 |
|  | Centre | Cancer unit | 399 (46.6) | 194 (54.8) | 593 (49.0) | 0 |
|  |  | Otherwise | 458 (53.4) | 160 (45.2) | 618 (51.0) |  |
| Blood sample | Serum CA125  *U/ml* | Median (IQR) | 26.0  (14.0-53.0) | 30.0  (16.0-74.0) | 27.0  (15.0-59.0) | 8 |
|  | HE4  *pmol/l* | Median (IQR) | 51.0  (43.0-65.0) | 51.0  (44.0-65.0) | 51.0  (43.0-65.0) | 97 |
| USG Scan | Ultrasound (U) score | 0 | 216 (25.2) | 49 (13.8) | 265 (21.9) | 253 |
|  |  | 1 | 318 (37.1) | 94 (26.6) | 412 (34.0) |  |
|  |  | 3 | 191 (22.3) | 90 (25.4) | 281 (23.2) |  |
|  | Maximum diameter of lesion *mm* | Median (IQR) | 71.0  (49.0-105.0) | 97.0  (66.0-138.0) | 77.0  (52.0-115.0) | 249 |
|  | Maximum diameter of largest solid part  *mm* | Median (IQR) | 0  (0-18.0) | 4.0  (0.0-28.0) | 0  (0-21.0) | 301 |
|  | Number of  cyst locules | *≤10* | 662 (77.3) | 200 (56.5) | 862 (71.2) | 259 |
|  |  | *>10* | 52 (6.1) | 31 (8.8) | 83 (6.9) |  |
|  |  | Not applicable | 7 (0.8) | 0 (0.0) | 7 (0.6) |  |
|  | Number of papillations | 0 | 569 (66.4) | 164 (46.3) | 733 (60.5) | 255 |
|  |  | 1 | 65 (7.6) | 30 (8.5) | 95 (7.8) |  |
|  |  | 2 | 31 (3.6) | 12 (3.4) | 43 (3.6) |  |
|  |  | 3 | 17 (2.0) | 6 (1.7) | 23 (1.9) |  |
|  |  | 4+ | 38 (4.4) | 20 (5.7) | 58 (4.8) |  |
|  |  | Not applicable | 4 (0.5) | 0 (0.0) | 4 (0.3) |  |
|  | Presence of  acoustic shadows | Yes | 145 (16.9) | 55 (15.5) | 200 (16.5) | 254 |
|  | Presence of ascites | Yes | 37 (4.3) | 20 (5.7) | 57 (4.7) | 249 |
|  | M1: Irregular  solid mass | Yes | 27 (3.2) | 19 (5.4) | 46 (3.8) | 259 |
|  | M2: Presence of ascites | Yes | 33 (3.9) | 19 (5.4) | 52 (4.3) | 249 |
|  | M3: At least four papillary structures | Yes | 39 (4.6) | 20 (5.7) | 59 (4.9) | 254 |
|  | M4: Irregular multilocular solid tumour with largest diameter ≥ 100mm | Yes | 54 (6.3) | 31 (8.8) | 85 (7.0) | 260 |
|  | M5: Very strong blood flow (colour score 4) | Yes | 46 (5.4) | 24 (6.8) | 70 (5.8) | 254 |
|  | B1: Unilocular cyst | Yes | 287 (33.5) | 61 (17.2) | 348 (28.7) | 258 |
|  | B2: Presence of solid components where the largest solid component has a largest diameter ≤7mm | Yes | 39 (4.6) | 8 (2.3) | 47 (3.9) | 260 |
|  | B3: Presence of acoustic shadows | Yes | 144 (16.8) | 56 (15.8) | 200 (16.5) | 254 |
|  | B4: Smooth multilocular tumour with largest diameter <100mm | Yes | 142 (16.6) | 24 (6.8) | 166 (13.7) | 259 |
|  | B5: No blood flow (colour score 1) | Yes | 447 (52.2) | 104 (29.4) | 551 (45.5) | 256 |

IQR=Interquartile range.

Cohort 1 are participants recruited up until June 2018, which includes both pre-surgical and conservatively managed patients.

Cohort 2 are participants recruited post June 2018 following the study protocol adaptation.

***Supplementary Table 2.*** Diagnostic performance statistics of index test combinations for the Secondary Outcome in Cohort 1

| **Index test**  **combination** | **Threshold** | **Diagnosis based on**  **reference standard**  n=836 | | **Number of**  **Participants** n (%) | **Sensitivity**  (%) (95% CI) | **Specificity**  (%) (95% CI) | **C-index**  (AUC)  (95% CI) | **Positive**  **predictive**  **value**  (PPV)  (%) (95%CI) | **Negative**  **predictive**  **value**  (NPV)  (%) (95% CI) | **Pairwise comparison with RMI 1^a^**  (250) (95% CI), p-value, number of participants |
| --- | --- | --- | --- | --- | --- | --- | --- | --- | --- | --- |
|  |  | OC  n=86 | No OC  n=750 |  |  |  |  |  |  |  |
| RMI 1,  n (%) | Missing | 4 (4.7) | 125 (16.7) |  | | | | | | |
|  | ≥200 | 32 (37.2) | 29 (3.9) | 707 (84.6) | 39.0  (28.4, 50.4) | 95.4  (93.4, 96.9) | 0.805  (0.752, 0.857) | 52.5 (39.3,65.4) | 92.3  (89.9, 94.2) | Se: -4.9 (-10.8, 1.0), p=0.1250  Sp: 1.1 (0.1, 2.1), p=0.0156, n=707 |
|  | <200 | 50 (58.1) | 596 (79.5) |  |  |  |  |  |  |  |
|  | **≥250** | **28 (32.6)** | **22 (2.9)** |  | **39.0**  **(28.4, 50.4)** | **95.4**  **(93.4, 96.9)** |  | **56.0**  **(41.3, 70.0)** | **91.8**  **(89.4, 93.8)** | * |
|  | **<250** | **54 (62.8)** | **603 (80.4)** |  |  |  |  |  |  |  |
| ROMA,  n (%) | Missing | 3 (3.5) | 48 (6.4) |  | | | | | | |
|  | ≥7.4% | 76 (88.4) | 372 (49.6) | 785 (93.9) | 91.6  (83.4, 96.5) | 47.0  (43.3, 50.8) | 0.814  (0.759, 0.868) | 17.0  (13.6, 20.8) | 97.9  (95.8, 99.2) | Se: -57.0 (-69.1, -44.8), p<0.0001  Sp: 49.3 (45.0, 53.7), p<0.0001, n=669 |
|  | <7.4% | 7 (8.1) | 330 (44.0) |  |  |  |  |  |  |  |
|  | ≥11.4% | 58 (67.4) | 189 (25.2) |  | 69.9  (58.8, 79.5) | 73.1  (69.6, 76.3) |  | 23.5  (18.3, 29.3) | 95.4  (93.2, 97.0) | Se: -35.4 (-47.8, -23.1), p<0.0001  Sp: 23.6 (19.6, 27.5), p<0.0001, n=669 |
|  | <11.4% | 25 (29.1) | 513 (68.4) |  |  |  |  |  |  |  |
|  | ≥12.5% | 57 (66.3) | 164 (21.9) |  | 68.7  (57.6, 78.4) | 68.6  (64.4, 72.5) |  | 25.8  (20.2, 32.1) | 93.2  (90.2, 95.5) | Se: -34.2 (-46.5, -21.9), p<0.0001  Sp: 19.8 (16.0, 23.6), p<0.0001, n=669 |
|  | <12.5% | 26 (30.2) | 358 (47.7) |  |  |  |  |  |  |  |
|  | ≥13.1% | 55 (64.0) | 149 (19.9) |  | 66.3  (55.1, 76.3) | 78.8  (75.6, 81.7) |  | 27.0  (21.0, 33.6) | 95.2  (93.1, 96.8) | Se: -31.6 (-43.8, -19.5), p<0.0001  Sp: 17.6 (13.9, 21.3), p<0.0001, n=669 |
|  | <13.1% | 28 (32.6) | 553 (73.7) |  |  |  |  |  |  |  |
| ADNEX,  n (%) | Missing | 5 (5.8) | 179 (23.9) |  | | | | | | |
|  | ≥3.0% | 71 (82.6) | 311 (41.5) | 652 (78.0) | 87.7  (78.5, 93.9) | 45.5  (41.4, 49.7) | 0.811  (0.751, 0.871) | 18.6  (14.8, 22.9) | 96.3  (93.3, 98.2) | Se: -54.3 (-66.4, -42.2), p<0.0001  Sp: 50.8 (46.5, 55.1), p<0.0001, n=652 |
|  | <3.0% (Secondary) | 10 (11.6) | 260 (34.7) |  |  |  |  |  |  |  |
|  | ≥10.0% | 61 (70.9) | 142 (18.9) |  | 75.3  (64.5, 84.2) | 75.1  (71.4, 78.6) |  | 30.0  (23.8, 36.9) | 95.5  (93.2, 97.3) | Se: -42.0 (-54.0, -30.0), p<0.0001  Sp: 21.2 (17.4, 25.0), p<0.0001, n=652 |
|  | <10.0% (Primary) | 20 (23.3) | 429 (57.2) |  |  |  |  |  |  |  |
| IOTA  sRisk  model,  n (%) | Missing | 4 (4.7) | 129 (17.2) |  | | | | | | |
|  | ≥3.0% | 67 (77.9) | 231 (30.8) | 703 (84.1) | 81.7  (71.6, 89.4) | 62.8  (58.9, 66.6) | 0.814  (0.759, 0.870) | 22.5  (17.9, 27.7) | 96.3  (94.0, 97.9) | Se: -47.6 (-60.1, -35.0), p<0.0001  Sp: 33.6 (29.5, 37.7), p<0.0001, n=701 |
|  | <3.0% (Secondary) | 15 (17.4) | 390 (52.0) |  |  |  |  |  |  |  |
|  | ≥10.0% | 61 (70.9) | 149 (19.9) |  | 74.4  (63.6, 83.4) | 76.0  (72.4, 79.3) |  | 29.0  (23.0, 35.7) | 95.7  (93.6, 97.3) | Se: -40.2 (-53.1, -27.4), p<0.0001  Sp: 20.4 (16.6, 24.1), p<0.0001, n=701 |
|  | <10.0% (Primary) | 21 (24.4) | 472 (62.9) |  |  |  |  |  |  |  |
| IOTA  simple  rules,  n (%) | Missing | 4 (4.7) | 124 (16.5) |  | | | | | | |
|  | Malignant | 33 (38.4) | 25 (3.3) | 576 (68.9) | 60.0  (45.9, 73.0) | 95.2  (93.0, 96.9) | 0.776  (0.710, 0.842) | 56.9  (43.2, 69.8) | 95.8  (93.6, 97.3) | Se: -23.6 (-38.8, -8.5), p=0.0023  Sp: 2.3 (-0.1, 4.7), p=0.0576, n=574 |
|  | Benign | 22 (25.6) | 496 (66.1) |  |  |  |  |  |  |  |
|  | Inconclusive | 27 (31.4) | 105 (14.0) |  | | | | | | |
| CA125,  n (%) | Missing | 0 (0.0) | 4 (0.5) |  | | | | | | |
|  | ≥87 U/ml | 40 (46.5) | 82 (10.9) | 832 (99.5) | 46.5  (35.7, 57.6) | 89.0  (86.5, 91.2) | 0.729  (0.666, 0.791) | 32.8  (24.6, 41.9) | 93.5  (91.5, 95.2) | Se: -11.0 (-19.0, -3.0), p=0.0039  Sp: 6.9 (4.6, 9.2), p<0.0001, n=707 |

OC=ovarian cancer; CI=confidence interval; AUC=area under the curve; Se=Sensitivity, Sp=Specificity, CI=confidence interval.

^a^ Differences in sensitivities and specificities not exact (A – B) for test A and test B, since different numbers of participants are included in each index test analysis, and only those with non-missing index test data for both index tests were included in the calculation of differences.

OC present includes primary invasive ovarian malignant neoplasm, secondary malignant neoplasms, borderline neoplasms, neoplasms of uncertain or unknown behaviour or diagnosed with cancer in the last 12 months; OC absent includes benign, normal or absent with ovarian cancer in the last 12 months.

Note that 21 (2.5%) had a diagnostic category of other and were not included in the analysis.

**Supplementary Figure 1a.** Receiver operating characteristic (ROC) plot of the index test combinations for the Secondary Outcome in Cohort 1
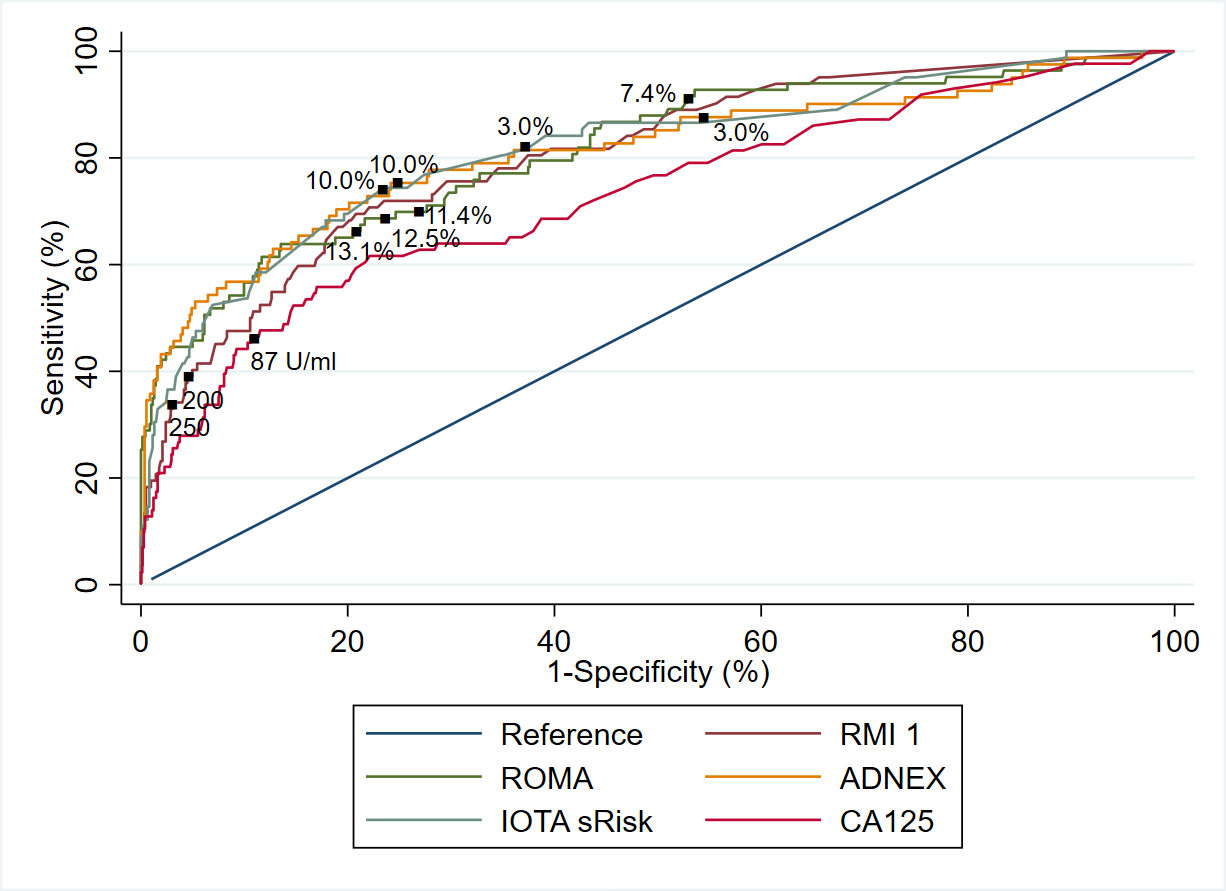

Note that the plot was produced by considering participants with available data for each index test combination separately, such that the number of participants used for each ROC curve varies.

IOTA simple rules not included in the ROC plot because some participants received inconclusive results.

**Supplementary Figure 1b**. Calibration plots for ROMA (left), IOTA sRisk (middle) and ADNEX (right) of the Secondary Outcome in Cohort 1

Calibration slope values (95% CIs) (derived from logistic regression model where the outcome is regressed on the predicted log-odds) were ROMA 1.26 (0.98, 1.54), IOTA ADNEX 0.67 (0.53, 0.80) and IOTA sRisk 0.46 (0.36, 0.55).

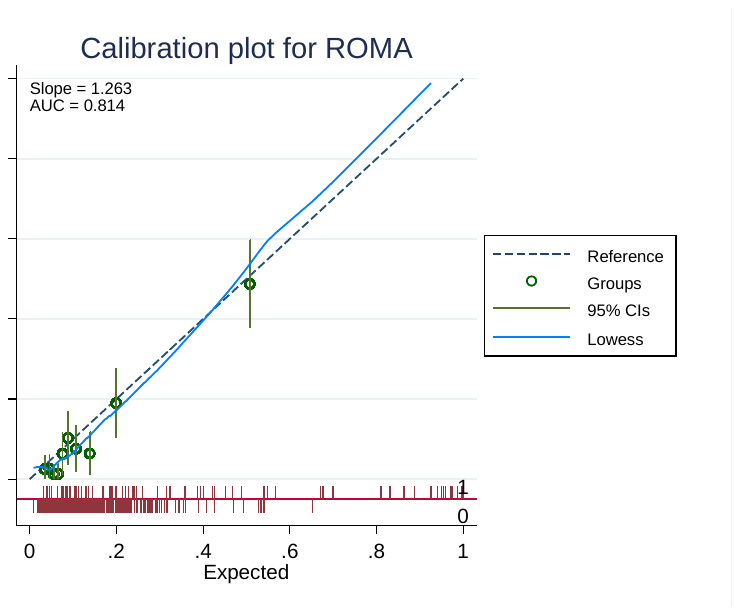

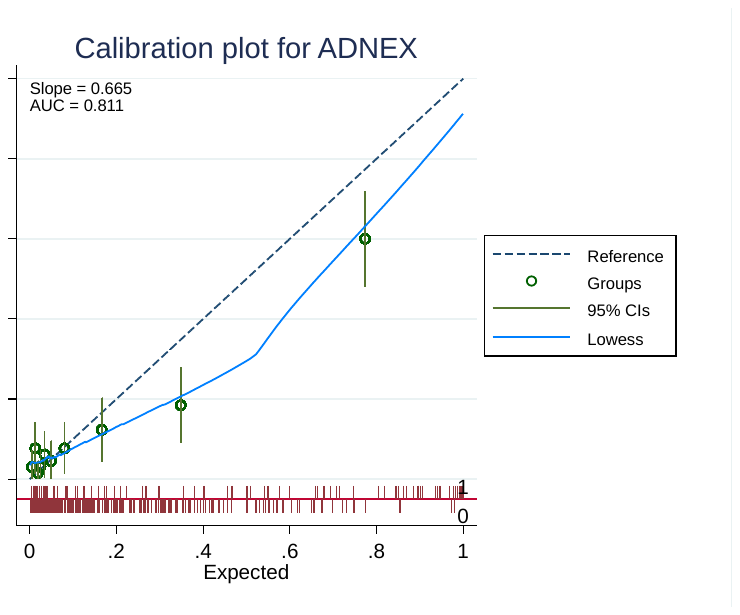

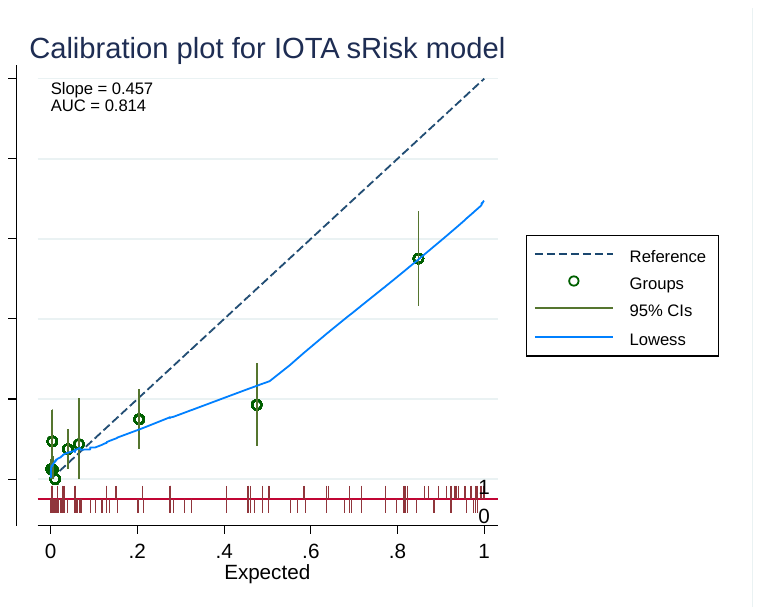

Note that the calibration slope and plot were only obtained for index test combinations that use a prediction model.

***Supplementary Table 3.*** Diagnostic performance statistics of index test combinations for the Primary Outcome in Combined Cohorts

| **Index test**  **combination** | **Threshold** | **Diagnosis based on**  **reference standard**  n=1,096 | | **Number of**  **Participants** n (%) | **Sensitivity**  (%) (95% CI) | **Specificity**  (%) (95% CI) | **C-index**  (AUC)  (95% CI) | **Positive**  **predictive**  **value** (PPV)  (%) (95% CI) | **Negative**  **predictive**  **value** (NPV)  (%) (95% CI) | **Pairwise comparison with RMI 1^a^**  (250) (95% CI), p-value, number of participants |
| --- | --- | --- | --- | --- | --- | --- | --- | --- | --- | --- |
|  |  | OC  n=88 | No OC  n=1,008 |  |  |  |  |  |  |  |
| RMI 1,  n (%) | Missing | 26 (29.5) | 203 (20.1) |  | | | | | | |
|  | ≥200 | 28 (31.8) | 39 (3.9) | 867 (79.1) | 45.2  (32.5, 58.3) | 95.2  (93.4, 96.5) | 0.847  (0.792,  0.901) | 41.8  (29.8, 54.5) | 95.8  (94.1, 97.0) | Se: -4.8 (-11.8, 2.1), p=0.2500  Sp: 1.2 (0.4, 2.1), p=0.0020, n=867 |
|  | <200 | 34 (38.6) | 766 (76.0) |  |  |  |  |  |  |  |
|  | **≥250** | **25 (28.4)** | **29 (2.9)** |  | **40.3**  **(28.1, 53.6)** | **96.4**  **(94.9, 97.6)** |  | **46.3**  **(32.6, 60.4)** | **95.4**  **(93.8, 96.8)** | * |
|  | **<250** | **37 (42.0)** | **776 (77.0)** |  |  |  |  |  |  |  |
| ROMA,  n (%) | Missing | 5 (5.7) | 74 (7.3) |  | | | | | | |
|  | ≥7.4% | 75 (85.2) | 488 (48.4) | 1,017 (92.8) | 90.4  (81.9, 95.7) | 47.8  (44.5, 51.0) | 0.842  (0.787, 0.896) | 13.3  (10.6, 16.4) | 98.2  (96.6, 99.2) | Se: -46.7 (-61.8, -31.6), p<0.001  Sp: 47.5 (43.7, 51.4), p<0.001, n=811 |
|  | <7.4% | 8 (9.1) | 446 (46.2) |  |  |  |  |  |  |  |
|  | ≥11.4% | 66 (75.0) | 241 (23.9) |  | 79.5  (69.2, 87.6) | 74.2  (71.3, 77.0) |  | 21.5  (17.0, 26.5) | 97.6  (96.2, 98.6) | Se: -38.3 (-53.1, -23.5), p<0.001  Sp: 22.1 (18.7, 25.5), p<0.001, n=811 |
|  | <11.4% | 17 (19.3) | 693 (68.8) |  |  |  |  |  |  |  |
|  | ≥12.5% | 63 (71.6) | 208 (20.6) |  | 75.9  (65.3, 84.6) | 77.7  (74.9, 80.4) |  | 23.2  (18.4, 28.7) | 97.3  (95.9, 98.4) | Se: -36.7 (-51.4, -22.0), p<0.001  Sp: 18.4 (15.1, 21.6), p<0.001, n=811 |
|  | <12.5% | 20 (22.7) | 726 (72.0) |  |  |  |  |  |  |  |
|  | ≥13.1% | 61 (69.3) | 188 (18.7) |  | 73.5  (62.7, 82.6) | 79.9  (77.2, 82.4) |  | 24.5  (19.3, 30.3) | 97.1  (95.7, 98.2) | Se: -35.0 (-49.6, -20.4), p<0.001  Sp: 16.1 (13.0, 19.3), p<0.001, n=811 |
|  | <13.1% | 22 (25.0) | 746 (74.0) |  |  |  |  |  |  |  |
| ADNEX,  n (%) | Missing | 28 (31.8) | 265 (26.3) |  | | | | | | |
|  | ≥3.0% | 56 (63.6) | 421 (41.8) | 803 (73.3) | 93.3  (83.8, 98.2) | 43.3  (39.7, 47.0) | 0.886  (0.830, 0.942) | 11.7  (9.0, 15.0) | 98.8  (96.9, 99.7) | Se: -53.3 (-67.6, -39.0), p<0.001  Sp: 52.9 (49.2, 56.6), p<0.001, n=803 |
|  | <3.0% (Secondary) | 4 (4.5) | 322 (31.9) |  |  |  |  |  |  |  |
|  | ≥10.0% | 54 (61.4) | 199 (19.7) |  | 90.0  (79.5, 96.2) | 73.2  (69.9, 76.4) |  | 21.3  (16.5, 26.9) | 98.9  (97.6, 99.6) | Se: -50.0 (-64.3, -35.7), p<0.001  Sp: 23.0 (19.7, 26.3), p<0.001, n=803 |
|  | <10.0% (Primary) | 6 (6.8) | 544 (54.0) |  |  |  |  |  |  |  |
| IOTA  sRisk  model,  n (%) | Missing | 27 (30.7) | 212 (21.0) |  | | | | | | |
|  | ≥3.0% | 54 (61.4) | 336 (33.3) | 857 (78.2) | 88.5  (77.8, 95.3) | 57.8  (54.3, 61.2) | 0.857  (0.802, 0.912) | 13.8  (10.6, 17.7) | 98.5  (96.9, 99.4) | Se: -47.5 (-61.7, -33.4), p<0.001  Sp: 38.5 (34.9, 42.2), p<0.001, n=855 |
|  | <3.0% (Secondary) | 7 (8.0) | 460 (45.6) |  |  |  |  |  |  |  |
|  | ≥10.0% | 51 (58.0) | 221 (21.9) |  | 83.6  (71.9, 91.8) | 72.2  (69.0, 75.3) |  | 18.8  (14.3, 23.9) | 98.3  (96.9, 99.2) | Se: -42.6 (-57.5, -27.8), p<0.001  Sp: 24.1 (20.7, 27.4), p<0.001, n=855 |
|  | <10.0% (Primary) | 10 (11.4) | 575 (57.0) |  |  |  |  |  |  |  |
| IOTA  simple  rules,  n (%) | Missing | 26 (29.6) | 203 (20.1) |  | | | | | | |
|  | Malignant | 32 (36.4) | 41 (4.1) | 692 (63.1) | 76.2  (60.5, 87.9) | 93.7  (91.5, 95.4) | 0.849  (0.784, 0.915) | 43.8  (32.2, 55.9) | 98.4  (97.0, 99.2) | Se: -31.0 (-48.8, -13.1), p=0.0010  Sp: 3.5 (1.3, 5.8), p=0.0011, n=690 |
|  | Benign | 10 (11.4) | 609 (60.4) |  |  |  |  |  |  |  |
|  | Inconclusive | 20 (22.7) | 155 (15.4) |  | | | | | | |
| CA125,  n (%) | Missing | 0 (0.0) | 6 (0.6) |  | | | | | | |
|  | ≥87 U/ml | 49 (55.7) | 121 (12.0) | 1,090 (99.5) | 55.7  (44.7, 66.3) | 87.9  (85.7, 89.9) | 0.785  (0.728, 0.842) | 28.8  (22.1, 36.3) | 95.8  (94.3, 97.0) | Se: -12.9 (-22.9, -2.9), p=0.0078  Sp: 7.7 (5.6, 9.8), p<0.001, n=867 |
|  | <87 U/ml | 39 (44.3) | 881 (87.4) |  |  |  |  |  |  |  |

OC=ovarian cancer; CI=confidence interval; AUC=area under the curve; Se=Sensitivity, Sp=Specificity, CI=confidence interval;

^a^Differences in sensitivities and specificities not exact (A – B) for test A and test B, since different numbers of participants are included in each index test analysis, and only those with non-missing index test data for both index tests were included in the calculation of differences.

OC present includes primary invasive ovarian malignant neoplasm or diagnosed with ovarian cancer in the last 12 month; OC absent includes benign, normal or absent with ovarian cancer in the last 12 months.

Note that 115 (9.5%) had a diagnostic category of other and were not included in the analysis.

**Supplementary Figure 2a.** Receiver operating characteristic (ROC) plot of the index test combinations for the Primary Outcome in the Combined Cohorts

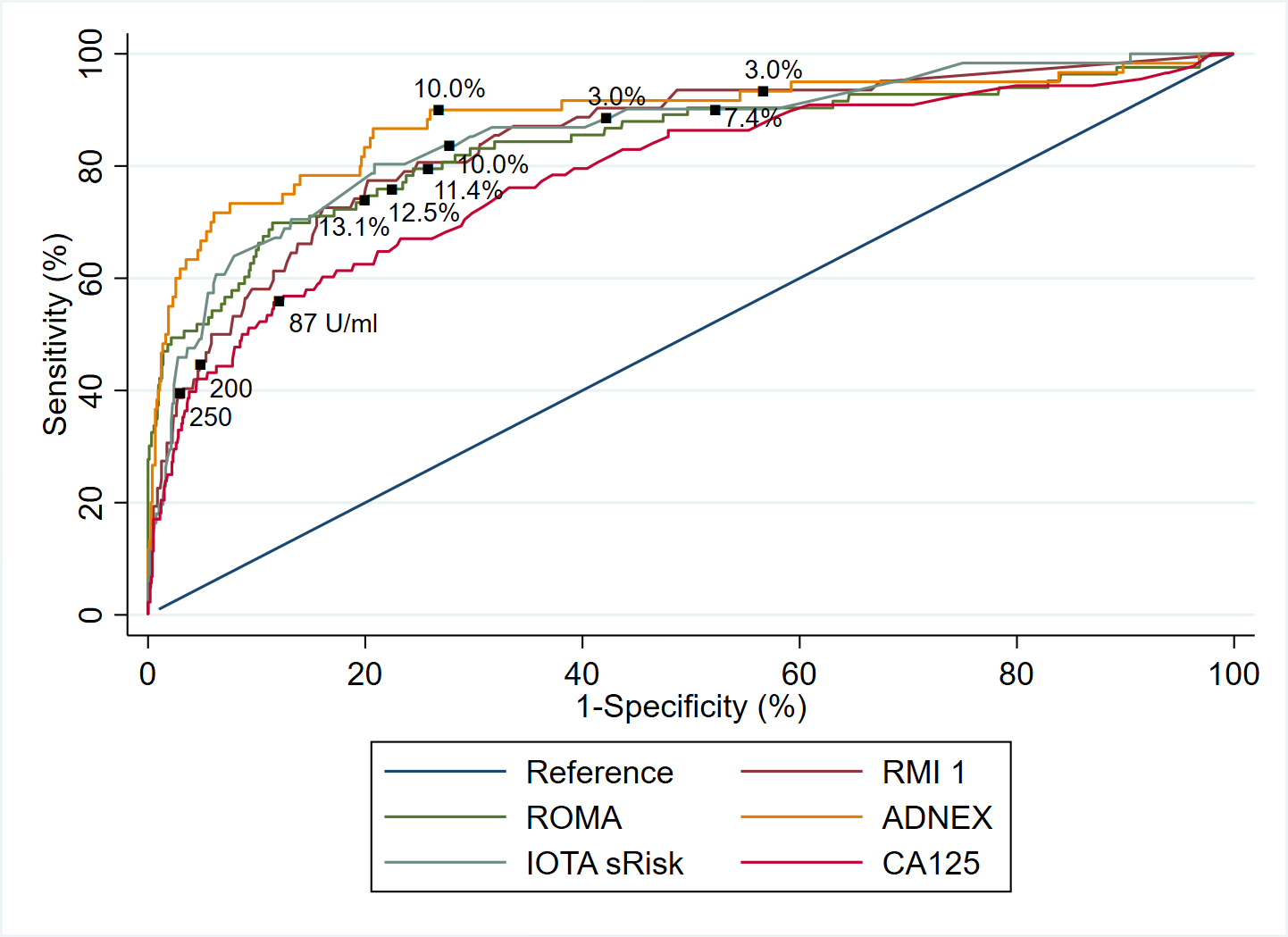

Note that the plot was produced by considering participants with available data for each index test combination separately, such that the number of participants used for each ROC curve varies.

IOTA simple rules not included in the ROC plot because some participants received inconclusive results.

**Supplementary Figure 2b.** Calibration plots for ROMA (left), IOTA sRisk (middle) and ADNEX (right) of the Primary Outcome in Combined Cohort 1

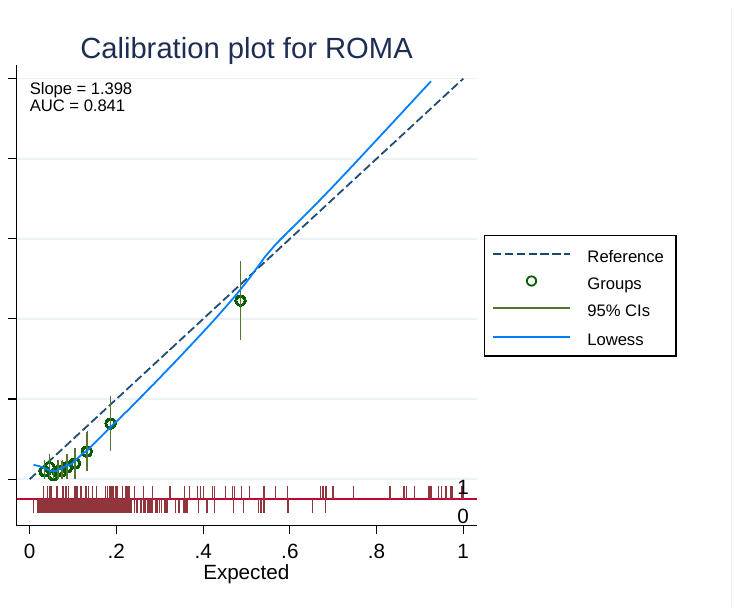

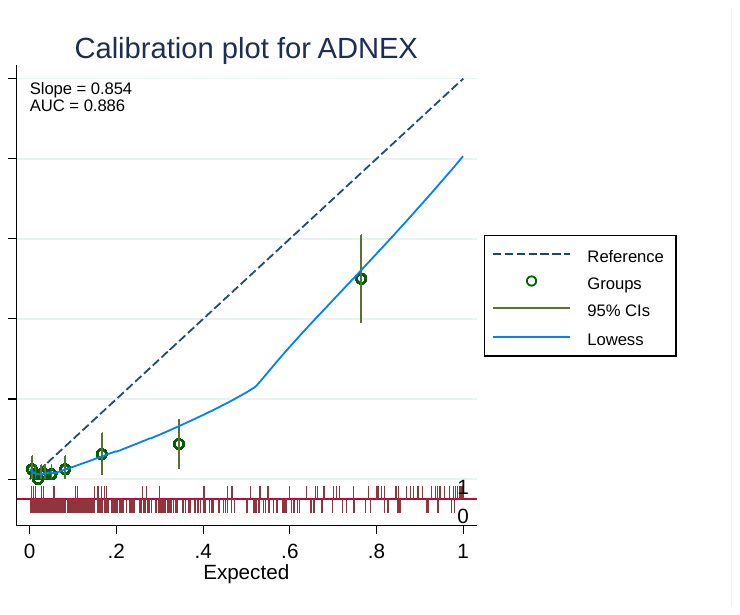

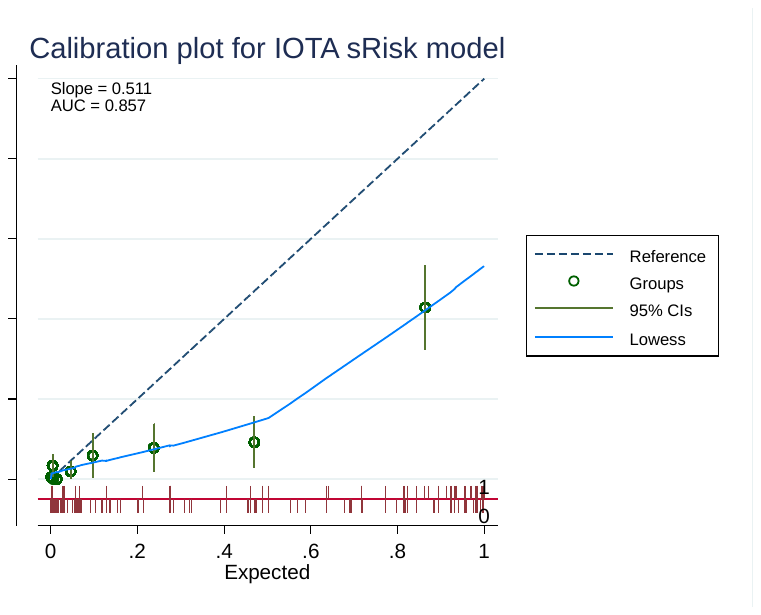

Primary outcome analysis in combined cohorts.

Calibration slope values (95% CIs) (derived from logistic regression model where the outcome is regressed on the predicted log-odds) were 1.40 (1.13, 1.67), 0.85 (0.68, 1.02) and 0.51 (0.40, 0.62) for ROMA, ADNEX and IOTA sRisk models respectively.

Note that the calibration slope and plot were only obtained for index test combinations that use a prediction model.

**Supplementary Table 4.** Diagnostic performance statistics of index test combinations for the Primary Outcome in Combined Cohorts with imputation of missing data.

| **Index test**  **combination** | **Threshold**  n=1,096 | **Sensitivity**  (%) (95% CI) | **Specificity**  (%) (95% CI) | **C-index**  (AUC)  (95% CI) | **Positive**  **predictive**  **value**  (PPV)  (%) (95% CI) | **Negative**  **predictive**  **value**  (NPV)  (%) (95% CI) | **Pairwise comparison with RMI 1^a^**  (250) (95% CI), p-value, number of participants |
| --- | --- | --- | --- | --- | --- | --- | --- |
| RMI 1,  n (%) | Missing |  | | | | | |
|  | ≥200 | 48.2  (37.2, 59.1) | 94.5  (93.0, 96.1) | 0.840  (0.788, 0.893) | 43.5  (33.2, 53.8) | 95.4  (94.1, 96.7) | Se: -3.7 (-18.4, 11.1), p=0.627  Sp: 1.5 (-0.4, 3.4), p=0.120 |
|  | <200 |  |  |  |  |  |  |
|  | **≥250** | **44.5**  **(33.6, 55.4)** | **96.0**  **(94.7, 97.4)** |  | **49.5**  **(37.8, 61.2)** | **95.2**  **(93.9, 96.5)** | * |
|  | **<250** |  |  |  |  |  |  |
| ROMA,  n (%) | Missing |  | | | | | |
|  | ≥7.4% | 90.0  (83.5, 96.6) | 47.8  (44.5, 51.0) | 0.838  (0.783, 0.894) | 13.1  (10.4, 15.8) | 98.2  (97.0, 99.4) | Se: -45.5 (-58.1, -32.9), p<0.001  Sp: 48.3 (44.9, 51.7), p<0.001 |
|  | <7.4% |  |  |  |  |  |  |
|  | ≥11.4% | 79.0  (70.1, 88.0) | 74.3  (71.5, 77.0) |  | 21.1  (16.7, 25.6) | 97.6  (96.5, 98.7) | Se: -34.5 (-48.5, -20.5), p<0.001  Sp: 21.8 (18.7, 24.8), p<0.001 |
|  | <11.4% |  |  |  |  |  |  |
|  | ≥12.5% | 75.5  (66.0, 85.0) | 77.7  (75.1, 80.4) |  | 22.8  (17.9, 27.7) | 97.3  (96.2, 98.5) | Se: -31.0 (-45.3, -16.7), p<0.001  Sp: 18.3 (15.4, 21.2), p<0.001 |
|  | <12.5% |  |  |  |  |  |  |
|  | ≥13.1% | 73.1  (63.3, 82.9) | 79.8  (77.2, 82.3) |  | 24.0  (18.8, 29.2) | 97.1  (96.0, 98.3) | Se: -28.6 (-43.1, -14.1), p<0.001  Sp: 16.2 (13.4, 19.1), p<0.001 |
|  | <13.1% |  |  |  |  |  |  |
| ADNEX,  n (%) | Missing |  | | | | | |
|  | ≥3.0% | 93.3  (86.9, 99.8) | 43.9  (40.4, 47.4) | 0.884  (0.828, 0.941) | 11.2  (8.4, 13.9) | 98.9  (97.8, 100.0) | Se: -48.8 (-61.3, -36.4), p<0.001  Sp: 52.1 (48.4, 55.9), p<0.001 |
|  | <3.0% (Secondary) |  |  |  |  |  |  |
|  | ≥10.0% | 89.2  (80.9, 97.4) | 73.6  (70.5, 76.7) |  | 20.3  (15.4, 25.3) | 98.9  (98.0, 99.8) | Se: -44.7 (-58.1, -31.2), p<0.001  Sp: 22.4 (19.0, 25.8), p<0.001 |
|  | <10.0% (Primary) |  |  |  |  |  |  |
| IOTA  sRisk  model,  n (%) | Missing |  | | | | | |
|  | ≥3.0% | 89.5  (82.2, 96.7) | 57.2  (53.6, 60.9) | 0.862  (0.814, 0.911) | 15.4  (12.2, 18.7) | 98.4  (97.3, 99.5) | Se: -45.0 (-57.6, -32.3), p<0.001  Sp: 38.8 (35.0, 42.6), p<0.001 |
|  | <3.0% (Secondary) |  |  |  |  |  |  |
|  | ≥10.0% | 83.9  (75.1, 92.7) | 71.6  (68.4, 74.7) |  | 20.5  (16.1, 24.8) | 98.1  (97.0, 99.2) | Se: -39.4 (-52.9, -25.9), p<0.001  Sp: 24.5 (21.0, 27.9), p<0.001 |
|  | <10.0% (Primary) |  |  |  |  |  |  |
| IOTA  simple  rules,  n (%) | Missing |  | | | | | |
|  | Malignant | 75.8  (63.0, 88.7) | 93.4  (91.3, 95.5) | 0.846  (0.782, 0.910) | 45.0  (33.5, 56.6) | 98.2  (97.1, 99.2) | Se: -31.3 (-47.5, -15.2), p<0.001  Sp: 2.6 (0.2, 5.1), p=0.036 |
|  | Benign |  |  |  |  |  |  |
|  | Inconclusive |  | | | | | |
| CA125,  n (%) | Missing |  | | | | | |
|  | ≥87 U/ml | 55.7  (45.2, 66.2) | 87.9  (85.9, 90.0) | 0.785  (0.728, 0.842) | 28.7  (21.9, 35.6) | 95.8  (94.5, 97.1) | Se: -11.2 (-26.1, 3.8), p=0.142  Sp: 8.1 (5.7, 10.5), p<0.001 |
|  | <87 U/ml |  |  |  |  |  |  |

OC=ovarian cancer; CI=confidence interval; AUC=area under the curve; Se=Sensitivity, Sp=Specificity, CI=confidence interval;

^a^ Differences in sensitivities and specificities not exact (A – B) for test A and test B, since different numbers of participants are included in each index test analysis, and only those with non-missing index test data for both index tests were included in the calculation of differences.

Calibration slope values (95% CIs) (derived from logistic regression model where the outcome is regressed on the predicted log-odds) were 1.38 (1.10, 1.65), 0.87 (0.70, 1.04) and 0.53 (0.42, 0.63) for ROMA, ADNEX and IOTA sRisk models respectively.

Note that the calibration slope and plot were only obtained for index test combinations that use a prediction model.

**Supplementary Table 5.** Diagnostic performance statistics of index test combinations for the Secondary Outcome in Combined Cohorts

| **Index test**  **combination** | **Threshold** | **Diagnosis based on**  **reference standard**  n=1,178 | | **Number of**  **Participants** n (%) | **Sensitivity**  (%) (95% CI) | **Specificity**  (%) (95% CI) | **C-index**  (AUC)  (95% CI) | **Positive**  **predictive**  **value**  (PPV)  (%) (95%CI) | **Negative**  **predictive**  **value**  (NPV)  (%) (95% CI) | **Pairwise comparison with RMI 1^a^**  (250) (95% CI), p-value, number of participants |
| --- | --- | --- | --- | --- | --- | --- | --- | --- | --- | --- |
|  |  | OC  n=170 | No OC  n=1,008 |  |  |  |  |  |  |  |
| RMI 1,  n (%) | Missing | 45 (26.5) | 203 (20.1) |  | | | | | | |
|  | ≥200 | 47 (27.6) | 39 (3.9) | 930 (78.9) | 37.6  (29.1, 46.7) | 95.2  (93.4, 96.5) | 0.797  (0.753, 0.840) | 54.7  (43.5, 65.4) | 90.8  (88.6, 92.6) | Se: -3.2 (-7.1, 0.7), p=0.1250  Sp: 1.2 (0.4, 2.1), p=0.0020, n=930 |
|  | <200 | 78 (45.9) | 766 (76.0) |  |  |  |  |  |  |  |
|  | **≥250** | **43 (25.3)** | **29 (2.9)** |  | **34.4**  **(26.1, 43.4)** | **96.4**  **(94.9, 97.6)** |  | **59.7**  **(47.5, 71.1)** | **90.4**  **(88.3, 92.3)** | * |
|  | **<250** | **82 (48.2)** | **776 (77.0)** |  |  |  |  |  |  |  |
| ROMA,  n (%) | Missing | 18 (10.6) | 74 (7.3) |  | | | | | | |
|  | ≥7.4% | 131 (77.1) | 488 (48.4) | 1,086 (92.1) | 86.2  (79.7, 91.2) | 47.8  (44.5, 51.0) | 0.788  (0.744, 0.832) | 21.2  (18.0, 24.6) | 95.5  (93.2, 97.2) | Se: -49.6 (-60.3, -38.8), p<0.001  Sp: 47.5 (43.7, 51.4), p<0.001, n=864 |
|  | <7.4% | 21 (12.4) | 446 (46.2) |  |  |  |  |  |  |  |
|  | ≥11.4% | 104 (61.2) | 241 (23.9) |  | 68.4  (60.4, 75.7) | 74.2  (71.3, 77.0) |  | 30.1  (25.3, 35.3) | 93.5  (91.5, 95.2) | Se: -32.7 (-43.3, -22.2), p<0.001  Sp: 22.1 (18.7, 25.5), p<0.001, n=864 |
|  | <11.4% | 48 (28.2) | 693 (68.8) |  |  |  |  |  |  |  |
|  | ≥12.5% | 100 (58.8) | 208 (20.6) |  | 65.8  (57.7, 73.3) | 77.7  (74.9, 80.4) |  | 32.5  (27.3, 38.0) | 93.3  (91.3, 95.0) | Se: -31.9 (-42.3, -21.4), p<0.001  Sp: 18.4 (15.1, 21.6), p<0.001, n=864 |
|  | <12.5% | 52 (30.6) | 726 (72.0) |  |  |  |  |  |  |  |
|  | ≥13.1% | 97 (57.1) | 188 (18.7) |  | 63.8  (55.6, 71.4) | 79.9  (77.2, 82.4) |  | 34.0  (28.6, 39.9) | 93.1  (91.2, 94.8) | Se: -30.1 (-40.4, -19.7), p<0.001  Sp: 16.1 (13.0, 19.3), p<0.001, n=864 |
|  | <13.1% | 55 (32.4) | 746 (74.0) |  |  |  |  |  |  |  |
| ADNEX,  n (%) | Missing | 47 (27.6) | 265 (26.3) |  | | | | | | |
|  | ≥3.0% | 111 (65.3) | 421 (41.8) | 866 (73.5) | 90.2  (83.6, 94.9) | 43.3  (39.7, 47.0) | 0.824  (0.779, 0.869) | 20.9  (17.5, 24.6) | 96.4  (93.8, 98.1) | Se: -56.1 (-65.7, -46.5), p<0.001  Sp: 52.9 (49.2, 56.6), p<0.001, n=866 |
|  | <3.0% (Secondary) | 12 (7.1) | 322 (31.9) |  |  |  |  |  |  |  |
|  | ≥10.0% | 98 (57.6) | 199 (19.7) |  | 79.7  (71.5, 86.4) | 73.2  (69.9, 76.4) |  | 33.0  (27.7, 38.7) | 95.6  (93.6, 97.1) | Se: -45.5 (-55.1, -35.9), p<0.001  Sp: 23.0 (19.7, 26.3), p<0.001, n=866 |
|  | <10.0% (Primary) | 25 (14.7) | 544 (54.0) |  |  |  |  |  |  |  |
| IOTA  sRisk  model,  n (%) | Missing | 45 (26.5) | 212 (21.0) |  | | | | | | |
|  | ≥3.0% | 106 (62.3) | 336 (33.3) | 921 (78.2) | 84.8  (77.3, 90.6) | 57.8  (54.3, 61.2) | 0.816  (0.772, 0.859) | 24.0  (20.1, 28.2) | 96.0  (93.9, 97.6) | Se: -50.0 (-60.2, -39.8), p<0.001  Sp: 38.5 (34.9, 42.2), p<0.001, n=918 |
|  | <3.0% (Secondary) | 19 (11.2) | 460 (45.6) |  |  |  |  |  |  |  |
|  | ≥10.0% | 96 (56.5) | 221 (21.9) |  | 76.8  (68.4, 83.9) | 72.2  (69.0, 75.3) |  | 30.3  (25.3, 35.7) | 95.2  (93.2, 96.8) | Se: -41.9 (-52.5, -31.4), p<0.001  Sp: 24.1 (20.7, 27.4), p<0.001, n=918 |
|  | <10.0% (Primary) | 29 (17.1) | 575 (57.0) |  |  |  |  |  |  |  |
| IOTA  simple  rules,  n (%) | Missing | 44 (25.9) | 203 (20.1) |  | | | | | | |
|  | Malignant | 53 (31.2) | 41 (4.1) | 734 (62.3) | 63.1  (51.9, 73.4) | 93.7  (91.5, 95.4) | 0.784  (0.731, 0.837) | 56.4  (45.8, 66.6) | 95.2  (93.2, 96.7) | Se: -25.3 (-38.0, -12.6), p=0.0001  Sp: 3.5 (1.3, 5.8), p=0.0011, n=731 |
|  | Benign | 31 (18.2) | 609 (60.4) |  |  |  |  |  |  |  |
|  | Inconclusive | 42 (24.7) | 155 (15.4) |  | | | | | | |
| CA125,  n (%) | Missing | 1 (0.6) | 6 (0.6) |  | | | | | | |
|  | ≥87 U/ml | 80 (47.1) | 121 (12.0) | 1,171 (99.4) | 47.3  (39.6, 55.1) | 87.9  (85.7, 89.9) | 0.723  (0.678, 0.769) | 39.8  (33.0, 46.9) | 90.8  (88.8, 92.6) | Se: -10.4 (-16.6, -4.2), p=0.0002  Sp: 7.7 (5.6, 9.8), p<0.001, n=930 |
|  | <87 U/ml | 89 (52.4) | 881 (87.4) |  |  |  |  |  |  |  |

OC=ovarian cancer; CI=confidence interval; AUC=area under the curve; Se=Sensitivity, Sp=Specificity, CI=confidence interval;

^a^Differences in sensitivities and specificities not exact (A – B) for test A and test B, since different numbers of participants are included in each index test analysis, and only those with non-missing index test data for both index tests were included in the calculation of differences.

OC present includes primary invasive ovarian malignant neoplasm, secondary malignant neoplasms, borderline neoplasms, neoplasms of uncertain or unknown behaviour or diagnosed with cancer in the last 12 months; OC absent includes benign, normal or absent with cancer in the last 12 months.

Note that 33 (2.7%) had a diagnostic category of other and were not included in the analysis.

**Supplementary Figure 3a.** Receiver operating characteristic (ROC) plot of the index test combinations for the Secondary Outcome in the Combined Cohorts

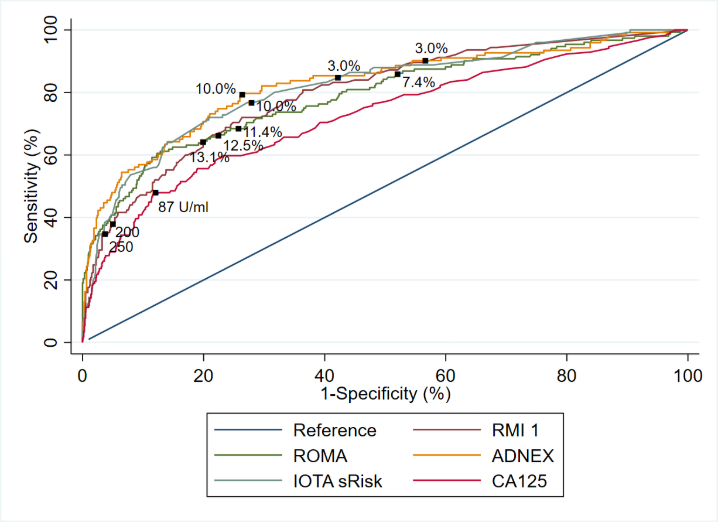

Note that the plot was produced by considering participants with available data for each index test combination separately, such that the number of participants used for each ROC curve varies.

IOTA simple rules not included in the ROC plot because some participants received inconclusive results.

**Supplementary Figure 3b.** Calibration plots for ROMA (left), IOTA sRisk (middle) and ADNEX (right) of the Secondary Outcome in Combined Cohorts

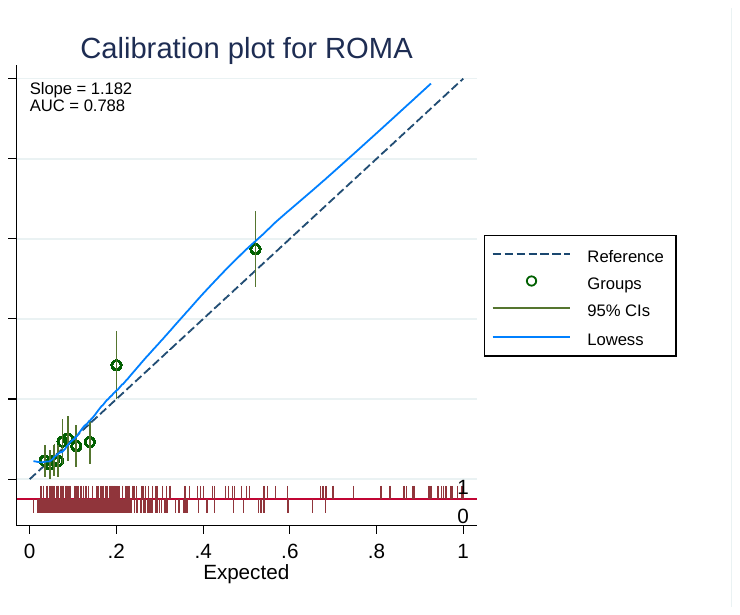

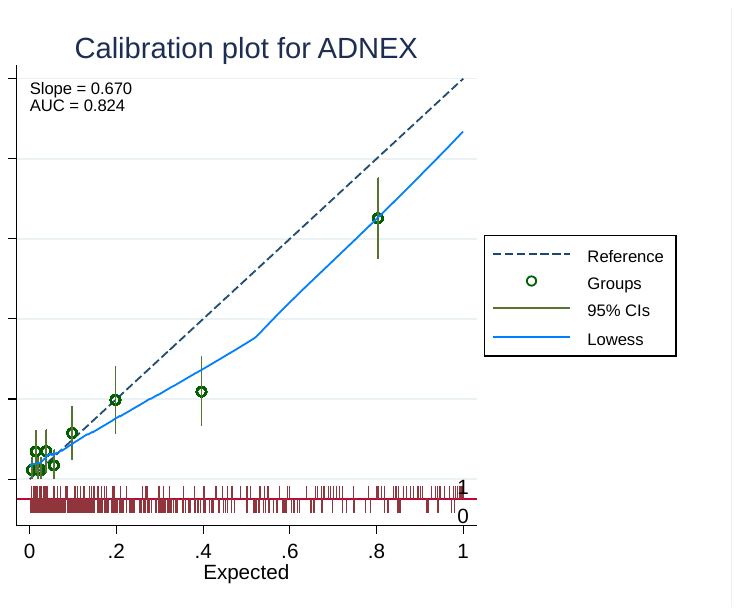

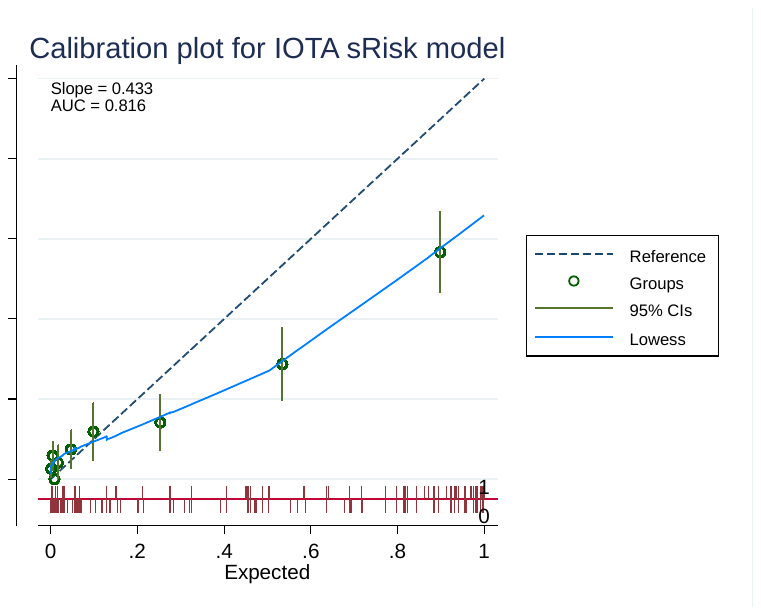

Calibration slope values (95% CIs) (derived from logistic regression model where the outcome is regressed on the predicted log-odds) were 1.18 (0.97, 1.40), 0.67 (0.56, 0.78) and 0.43 (0.36, 0.51) for ROMA, ADNEX and IOTA sRisk models respectively.

Note that the calibration slope and plot were only obtained for index test combinations that use a prediction model.

**Supplementary Table 6.** Diagnostic performance statistics of index test combinations for the Secondary Outcome in Cohort 1 with imputation for missing data.

| **Index test**  **combination** | **Threshold**  n=1,178 | **Sensitivity**  (%) (95% CI) | **Specificity**  (%) (95% CI) | **C-index**  (AUC)  (95% CI) | **Positive**  **predictive**  **value**  (PPV)  (%) (95% CI) | **Negative**  **predictive**  **value**  (NPV)  (%) (95% CI) | **Pairwise comparison with RMI 1^a^**  (250) (95% CI), p-value, number of participants |
| --- | --- | --- | --- | --- | --- | --- | --- |
| RMI 1,  n (%) | Missing |  | | | | | |
|  | ≥200 | 39.9  (32.2, 47.5) | 94.5  (93.0, 96.0) | 0.784  (0.741, 0.827) | 54.9  (45.5, 64.2) | 90.3  (88.5, 92.1) | Se: -3.4 (-13.8, 7.0), p=0.520  Sp: 1.5 (-0.4, 3.4), p=0.115 |
|  | <200 |  |  |  |  |  |  |
|  | **≥250** | **36.5**  **(28.9, 44.0)** | **96.0**  **(94.7, 97.3)** |  | **60.5**  **(50.5, 70.6)** | **90.0**  **(88.1, 91.8)** | * |
|  | **<250** |  |  |  |  |  |  |
| ROMA,  n (%) | Missing |  | | | | | |
|  | ≥7.4% | 85.3  (79.5, 91.0) | 47.7  (44.5, 50.9) | 0.780  (0.736, 0.825) | 21.6  (18.4, 24.7) | 95.0  (93.0, 97.1) | Se: -48.8 (-58.3, -39.3), p<0.001  Sp: 48.3 (44.9, 51.7), p<0.001 |
|  | <7.4% |  |  |  |  |  |  |
|  | ≥11.4% | 67.2  (59.7, 74.7) | 74.2  (71.4, 77.0) |  | 30.5  (25.7, 35.3) | 93.1  (91.2, 94.9) | Se: -30.7 (-41.4, -20.1), p<0.001  Sp: 21.8 (18.7, 24.9), p<0.001 |
|  | <11.4% |  |  |  |  |  |  |
|  | ≥12.5% | 64.5  (56.9, 72.2) | 77.7  (75.0, 80.4) |  | 32.8  (27.7, 38.0) | 92.9  (91.1, 94.6) | Se: -28.1 (-38.8, -17.3), p<0.001  Sp: 18.3 (15.3, 21.2), p<0.001 |
|  | <12.5% |  |  |  |  |  |  |
|  | ≥13.1% | 62.6  (54.9, 70.4) | 79.8  (77.3, 82.4) |  | 34.4  (28.9, 39.8) | 92.7  (90.9, 94.5) | Se: -26.2 (-37.0, -15.4), p<0.001  Sp: 16.2 (13.3, 19.0), p<0.001 |
|  | <13.1% |  |  |  |  |  |  |
| ADNEX,  n (%) | Missing |  | | | | | |
|  | ≥3.0% | 90.3  (85.1, 95.6) | 43.8  (40.4, 47.3) | 0.826  (0.781, 0.870) | 20.1  (16.7, 23.4) | 96.7  (94.8, 98.5) | Se: -53.9 (-63.0, -44.8), p<0.001  Sp: 52.2 (48.5, 55.9), p<0.001 |
|  | <3.0% (Secondary) |  |  |  |  |  |  |
|  | ≥10.0% | 79.4  (72.2, 86.7) | 73.5  (70.3, 76.6) |  | 31.9  (26.6, 37.1) | 95.8  (94.2, 97.4) | Se: -43.0 (-53.3, -32.6), p<0.001  Sp: 22.5 (19.1, 25.9), p<0.001 |
|  | <10.0% (Primary) |  |  |  |  |  |  |
| IOTA  sRisk  model,  n (%) | Missing |  | | | | | |
|  | ≥3.0% | 84.5  (78.4, 90.6) | 56.9  (53.5, 60.2) | 0.810  (0.767, 0.853) | 24.8  (21.2, 28.5) | 95.6  (93.8, 97.4) | Se: -48.1 (-57.5, -38.6), p<0.001  Sp: 39.1 (35.6, 42.6), p<0.001 |
|  | <3.0% (Secondary) |  |  |  |  |  |  |
|  | ≥10.0% | 76.1  (68.7, 83.5) | 71.3  (68.1, 74.4) |  | 30.9  (26.3, 35.5) | 94.7  (92.9, 96.4) | Se: -39.7 (-50.0, -29.4), p<0.001  Sp: 24.7 (21.4, 28.0), p<0.001 |
|  | <10.0% (Primary) |  |  |  |  |  |  |
| IOTA  simple  rules,  n (%) | Missing |  | | | | | |
|  | Malignant | 61.3  (50.4, 72.1) | 93.3  (91.4, 95.3) | 0.773  (0.718, 0.827) | 55.8  (45.5, 66.1) | 94.6  (92.8, 96.4) | Se: -24.8 (-37.5, -12.1), p<0.001  Sp: 2.7 (0.3, 5.0), p=0.028 |
|  | Benign |  |  |  |  |  |  |
|  | Inconclusive |  | | | | | |
| CA125,  n (%) | Missing |  | | | | | |
|  | ≥87 U/ml | 47.3  (39.7, 54.9) | 87.9  (85.9, 89.9) | 0.723  (0.677, 0.768) | 39.8  (33.0, 46.6) | 90.8  (89.0, 92.6) | Se: -10.8 (-21.5, -0.2), p=0.046  Sp: 8.1 (5.7, 10.5), p<0.001 |
|  | <87 U/ml |  |  |  |  |  |  |

OC=ovarian cancer; CI=confidence interval; AUC=area under the curve; Se=Sensitivity, Sp=Specificity, CI=confidence interval;

^a^ Differences in sensitivities and specificities not exact (A – B) for test A and test B, since different numbers of participants are included in each index test analysis, and only those with non-missing index test data for both index tests were included in the calculation of differences.

Calibration slope values (95% CIs) (derived from logistic regression model where the outcome is regressed on the predicted log-odds) were 1.14 (0.93, 1.35), 0.69 (0.57, 0.80) and 0.42 (0.35, 0.50) for ROMA, ADNEX and IOTA sRisk models respectively.

Note that the calibration slope and plot were only obtained for index test combinations that use a prediction model.

**Supplementary Table 7.** Diagnostic accuracy of Secondary Outcome definition with borderlines grouped with benign in Combined Cohorts

| **Index test**  **combination** | **Threshold** | Diagnosis based on  reference standard, n=1,178 | | **Number of**  **Participants** n (%) | **Sensitivity**  (%) (95% CI) | **Specificity**  (%) (95% CI) | **C-index**  (AUC)  (95% CI) | **Positive**  **predictive**  **value**  (PPV)  (%) (95%CI) | **Negative**  **predictive**  **value**  (NPV)  (%) (95% CI) | **Pairwise comparison with RMI 1^a^**  (250) (95% CI), p-value, number of participants |
| --- | --- | --- | --- | --- | --- | --- | --- | --- | --- | --- |
|  |  | OC  n=114 | No OC  n=1,064 |  |  |  |  |  |  |  |
| RMI 1,  n (%) | Missing | 32 (28.1) | 216 (20.3) |  | | | | | | |
|  | ≥200 | 33 (28.9) | 53 (5.0) | 930 (78.9) | 40.2  (29.6, 51.7) | 93.8  (91.9, 95.3) | 0.790  (0.735, 0.845) | 38.4  (28.1, 49.5) | 94.2  (92.4, 95.7) | Se: -4.9 (-10.8, 1.0), p=0.1250  Sp: 1.2 (0.3, 2.0), p=0.0020, n=930 |
|  | <200 | 49 (43.0) | 795 (74.7) |  |  |  |  |  |  |  |
|  | **≥250** | **29 (25.4)** | **43 (4.0)** |  | **35.4**  **(25.1, 46.7)** | **94.9**  **(93.2, 96.3)** |  | **40.3**  **(28.9, 52.5)** | **93.8**  **(92.0, 95.3)** | * |
|  | **<250** | **53 (46.5)** | **805 (75.7)** |  |  |  |  |  |  |  |
| ROMA,  n (%) | Missing | 9 (7.9) | 83 (7.8) |  | | | | | | |
|  | ≥7.4% | 95 (83.3) | 524 (49.2) | 1,086 (92.1) | 90.5  (83.2, 95.3) | 46.6  (43.4, 49.8) | 0.823  (0.773, 0.872) | 15.3  (12.6, 18.4) | 97.9  (96.1, 99.0) | Se: -51.9 (-65.0, -38.9), p<0.001  Sp: 47.4 (43.6, 51.2), p<0.001, n=864 |
|  | <7.4% | 10 (8.8) | 457 (43.0) |  |  |  |  |  |  |  |
|  | ≥11.4% | 79 (69.3) | 266 (25.0) |  | 75.2  (65.9, 83.1) | 72.9  (70.0, 75.6) |  | 22.9  (18.6, 27.7) | 96.5  (94.9, 97.7) | Se: -39.0 (-51.7, -26.2), p<0.001  Sp: 22.0 (18.6, 25.3), p<0.001, n=864 |
|  | <11.4% | 26 (22.8) | 715 (67.2) |  |  |  |  |  |  |  |
|  | ≥12.5% | 76 (66.7) | 232 (21.8) |  | 72.4  (62.8, 80.7) | 76.4  (73.6, 79.0) |  | 24.7  (20.0, 29.9) | 96.3  (94.7, 97.5) | Se: -37.7 (-50.4, -25.0), p<0.001  Sp: 18.4 (15.2, 21.6), p<0.001, n=864 |
|  | <12.5% | 29 (25.4) | 749 (70.4) |  |  |  |  |  |  |  |
|  | ≥13.1% | 73 (64.0) | 212 (19.9) |  | 69.5  (59.8, 78.1) | 78.4  (75.7, 80.9) |  | 25.6  (20.6, 31.1) | 96.0  (94.4, 97.3) | Se: -35.1 (-47.6, -22.5), p<0.001  Sp: 16.3 (13.2, 19.4), p<0.001, n=864 |
|  | <13.1% | 32 (28.1) | 769 (72.3) |  |  |  |  |  |  |  |
| ADNEX,  n (%) | Missing | 34 (29.8) | 278 (26.1) |  | | | | | | |
|  | ≥3.0% | 70 (61.4) | 462 (43.4) | 866 (73.5) | 87.5  (78.2, 93.8) | 41.2  (37.8, 44.8) | 0.816  (0.756, 0.877) | 13.2  (10.4, 16.3) | 97.0  (94.6, 98.6) | Se: -52.5 (-64.7, -40.3), p<0.001  Sp: 53.4 (49.8, 57.1), p<0.001, n=866 |
|  | <3.0% (Secondary) | 10 (8.8) | 324 (30.5) |  |  |  |  |  |  |  |
|  | ≥10.0% | 64 (56.1) | 233 (21.9) |  | 80.0  (69.6, 88.1) | 70.4  (67.0, 73.5) |  | 21.5  (17.0, 26.7) | 97.2  (95.5, 98.4) | Se: -45.0 (-57.2, -32.8), p<0.001  Sp: 24.3 (21.0, 27.6), p<0.001, n=866 |
|  | <10.0% (Primary) | 16 (14.0) | 553 (52.0) |  |  |  |  |  |  |  |
| IOTA  sRisk  model,  n (%) | Missing | 33 (28.9) | 224 (21.1) |  | | | | | | |
|  | ≥3.0% | 68 (59.6) | 374 (35.2) | 921 (78.2) | 84.0  (74.1, 91.2) | 55.5  (52.0, 58.9) | 0.807  (0.750, 0.863) | 15.4  (12.1, 19.1) | 97.3  (95.4, 98.5) | Se: -48.1 (-60.3, -36.0), p<0.001  Sp: 39.3 (35.7, 42.9), p<0.001, n=918 |
|  | <3.0% (Secondary) | 13 (11.4) | 466 (43.8) |  |  |  |  |  |  |  |
|  | ≥10.0% | 62 (54.4) | 255 (24.0) |  | 76.5  (65.8, 85.2) | 69.6  (66.4, 72.7) |  | 19.6  (15.3, 24.4) | 96.9  (95.1, 98.1) | Se: -40.7 (-53.2, -28.3), p<0.001  Sp: 25.1 (21.7, 28.4), p<0.001, n=918 |
|  | <10.0% (Primary) | 19 (16.7) | 585 (55.0) |  |  |  |  |  |  |  |
| IOTA  simple  rules,  n (%) | Missing | 32 (28.1) | 215 (20.2) |  | | | | | | |
|  | Malignant | 41 (36.0) | 53 (5.0) | 734 (62.3) | 69.5  (56.1, 80.8) | 92.1  (89.9, 94.1) | 0.808  (0.748, 0.868) | 43.6  (33.4, 54.2) | 97.2  (95.6, 98.3) | Se: -32.2 (-46.7, -17.7), p<0.001  Sp: 3.7 (1.4, 6.0), p=0.0010, n=731 |
|  | Benign | 18 (15.8) | 622 (58.5) |  |  |  |  |  |  |  |
|  | Inconclusive | 23 (20.2) | 174 (16.4) |  | | | | | | |
| CA125,  n (%) | Missing | 0 (0.0) | 7 (0.7) |  | | | | | | |
|  | ≥87 U/ml | 57 (50.0) | 144 (13.5) | 1,171 (99.4) | 50.0  (40.5, 59.5) | 86.4  (84.2, 88.4) | 0.730  (0.674, 0.785) | 28.4  (22.2, 35.1) | 94.1  (92.5, 95.5) | Se: -11.0 (-19.0, -3.0), p=0.0039  Sp: 7.8 (5.8, 9.8), p<0.001, n=930 |
|  | <87 U/ml | 57 (50.0) | 913 (85.8) |  |  |  |  |  |  |  |

OC=ovarian cancer; CI=confidence interval; AUC=area under the curve; Se=Sensitivity, Sp=Specificity, CI=confidence interval;

^a^ Differences in sensitivities and specificities not exact (A – B) for test A and test B, since different numbers of participants are included in each index test analysis, and only those with non-missing index test data for both index tests were included in the calculation of differences.

OC present includes primary invasive ovarian malignant neoplasm, secondary malignant neoplasms or diagnosed with cancer in the last 12 months; OC absent includes benign, normal, borderline neoplasms, neoplasms of uncertain or unknown behaviour or absent with cancer in the last 12 months.

Note that 33 (2.7%) had a diagnostic category of other and were not included in the analysis.

**Supplementary Table 8.** Diagnostic performance statistics of ORADS in the Primary Outcome in Combined Cohorts.

| **Index test**  **combination** | **Threshold** | **Diagnosis based on**  **reference standard**  n=1,096 | | **Number of**  **Participants** n (%) | **Sensitivity**  (%) (95% CI) | **Specificity**  (%) (95% CI) | **C-index**  (AUC)  (95% CI) | **Positive**  **predictive**  **value**  (PPV)  (%) (95% CI) | **Negative**  **predictive**  **value**  (NPV)  (%) (95% CI) | **Pairwise comparison with RMI 1^a^**  (250) (95% CI), p-value, number of participants |
| --- | --- | --- | --- | --- | --- | --- | --- | --- | --- | --- |
|  |  | OC  n=88 | No OC  n=1,008 |  |  |  |  |  |  |  |
| RMI 1,  n (%) | Missing | 26 (29.5) | 203 (20.1) |  | | | | | | |
|  | ≥200 | 28 (31.8) | 39 (3.9) | 867 (79.1) | 45.2  (32.5, 58.3) | 95.2  (93.4, 96.5) | 0.847  (0.792, 0.901) | 41.8  (29.8, 54.5) | 95.8  (94.1, 97.0) | Se: -4.8 (-11.8, 2.1), p=0.2500  Sp: 1.2 (0.4, 2.1), p=0.0020, n=867 |
|  | <200 | 34 (38.6) | 766 (76.0) |  |  |  |  |  |  |  |
|  | **≥250** | **25 (28.4)** | **29 (2.9)** |  | **40.3**  **(28.1, 53.6)** | **96.4**  **(94.9, 97.6)** |  | **46.3**  **(32.6, 60.4)** | **95.4**  **(93.8, 96.8)** | * |
|  | **<250** | **37 (42.0)** | **776 (77.0)** |  |  |  |  |  |  |  |
| ADNEX,  n (%) | Missing | 28 (31.8) | 265 (26.3) |  | | | | | | |
|  | ≥3.0% | 56 (63.6) | 421 (41.8) | 803 (73.3) | 93.3  (83.8, 98.2) | 43.3  (39.7, 47.0) | 0.886  (0.830, 0.942) | 11.7  (9.0, 15.0) | 98.8  (96.9, 99.7) | Se: -53.3 (-67.6, -39.0), p<0.001  Sp: 52.9 (49.2, 56.6), p<0.001, n=803 |
|  | <3.0% (Secondary) | 4 (4.5) | 322 (31.9) |  |  |  |  |  |  |  |
|  | ≥10.0% | 54 (61.4) | 199 (19.7) |  | 90.0  (79.5, 96.2) | 73.2  (69.9, 76.4) |  | 21.3  (16.5, 26.9) | 98.9  (97.6, 99.6) | Se: -50.0 (-64.3, -35.7), p<0.001  Sp: 23.0 (19.7, 26.3), p<0.001, n=803 |
|  | <10.0% (Primary) | 6 (6.8) | 544 (54.0) |  |  |  |  |  |  |  |
| ORADS,  n (%) | Missing | 24 (27.3) | 94 (9.3) |  | | | | | | |
|  | ≥4 | 52 (59.1) | 163 (16.2) | 978 (89.2) | 81.3  (69.5, 89.9) | 82.2  (79.5, 84.6) | 0.850  (0.795, 0.905) | 24.2  (18.6, 30.5) | 98.4  (97.3, 99.2) | Se: -43.5 (-58.3, 28.8), p<0.001  Sp: 16.9 (13.9, 19.9), p<0.001, n=859 |
|  | <4 | 12 (13.6) | 751 (74.5) |  |  |  |  |  |  |  |

OC=ovarian cancer; CI=confidence interval; AUC=area under the curve; Se=Sensitivity, Sp=Specificity, CI=confidence interval;

^a^Differences in sensitivities and specificities not exact (A – B) for test A and test B, since different numbers of participants are included in each index test analysis, and only those with non-missing index test data for both index tests were included in the calculation of differences.

OC present includes primary invasive ovarian malignant neoplasm or diagnosed with ovarian cancer in the last 12 month; OC absent includes benign, normal or absent with ovarian cancer in the last 12 months. Note that 115 (9.5%) had a diagnostic category of other and were not included in the analysis.

**Supplementary Table 9. Diagnostic performance statistics of ORADS in the Secondary Outcome in Combined Cohorts.**

| **Index test**  **combination** | **Threshold** | **Diagnosis based on**  **reference standard**  n=1,178 | | **Number of**  **Participants** n (%) | **Sensitivity**  (%) (95% CI) | **Specificity**  (%) (95% CI) | **C-index**  (AUC)  (95% CI) | **Positive**  **predictive**  **value**  (PPV)  (%) (95% CI) | **Negative**  **predictive**  **value**  (NPV)  (%) (95% CI) | **Pairwise comparison with RMI 1^a^**  (250) (95% CI), p-value, number of participants |
| --- | --- | --- | --- | --- | --- | --- | --- | --- | --- | --- |
|  |  | OC  n=170 | No OC  n=1,008 |  |  |  |  |  |  |  |
| RMI 1,  n (%) | Missing | 45 (26.5) | 203 (20.1) |  | | | | | | |
|  | ≥200 | 47 (27.6) | 39 (3.9) | 930 (78.9) | 37.6  (29.1, 46.7) | 95.2  (93.4, 96.5) | 0.797  (0.753, 0.840) | 54.7  (43.5, 65.4) | 90.8  (88.6, 92.6) | Se: -3.2 (-7.1, 0.7), p=0.1250  Sp: 1.2 (0.4, 2.1), p=0.0020, n=930 |
|  | <200 | 78 (45.9) | 766 (76.0) |  |  |  |  |  |  |  |
|  | **≥250** | **43 (25.3)** | **29 (2.9)** |  | **34.4**  **(26.1, 43.4)** | **96.4**  **(94.9, 97.6)** |  | **59.7**  **(47.5, 71.1)** | **90.4**  **(88.3, 92.3)** | * |
|  | **<250** | **82 (48.2)** | **776 (77.0)** |  |  |  |  |  |  |  |
| ORADS,  n (%) | Missing | 38 (22.4) | 94 (9.3) |  | | | | | | |
|  | ≥4 | 91 (53.5) | 163 (16.2) | 1,046 (88.8) | 68.9  (60.3, 76.7) | 82.2  (79.5, 84.6) | 0.790  (0.745, 0.835) | 35.8  (29.9, 42.1) | 94.8  (93.0, 96.3) | Se: -37.6 (-48.0, -27.2), p<0.001  Sp: 16.9 (13.9, 19.9), p<0.001, n=922 |
|  | <4 | 41 (24.1) | 751 (74.5) |  |  |  |  |  |  |  |

OC=ovarian cancer; CI=confidence interval; Se=Sensitivity, Sp=Specificity, CI=confidence interval;

^a^Differences in sensitivities and specificities not exact (A – B) for test A and test B, since different numbers of participants are included in each index test analysis, and only those with non-missing index test data for both index tests were included in the calculation of differences.

OC present includes primary invasive ovarian malignant neoplasm, secondary malignant neoplasms, borderline neoplasms, neoplasms of uncertain or unknown behaviour or diagnosed with cancer in the last 12 months; OC absent includes benign, normal or absent with cancer in the last 12 months. Note that 21 (2.5%) had a diagnostic category of other and were not included in the analysis.

**Supplementary Figure 4. Implementation of IOTA ADNEX ultrasound triage in secondary care (modified from ESGO 2023).**
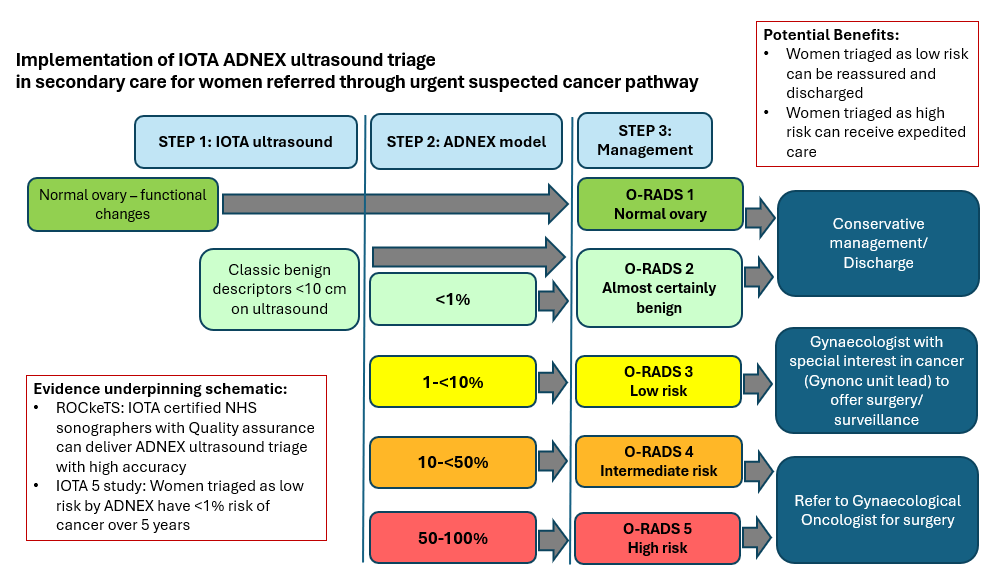

**Supplementary Figure 5. Implementation of IOTA ADNEX in primary care for women with symptoms (proposed future research).**

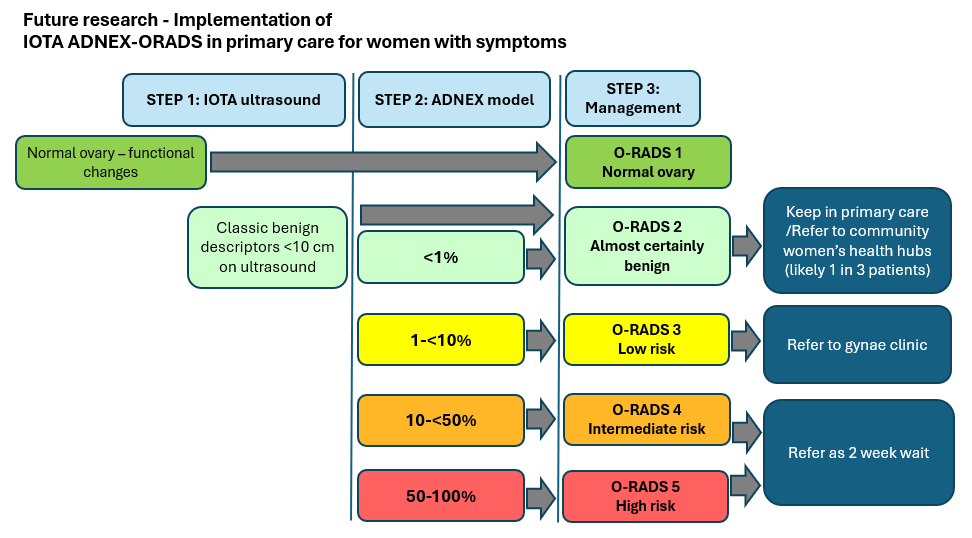

### APPENDIX B – Reporting Guidelines

**Supplementary Table 10.** Standards for the Reporting of Diagnostic Accuracy Studies (STARD) guidelines.

|  | **Section & Topic** | **No** | **Item** | **Reported on**  **page #** |
| --- | --- | --- | --- | --- |
|  | **TITLE OR ABSTRACT** |  |  |  |
|  |  | **1** | Identification as a study of diagnostic accuracy using at least one measure of accuracy  (such as sensitivity, specificity, predictive values, or AUC) | 2 |
|  | **ABSTRACT** |  |  |  |
|  |  | **2** | Structured summary of study design, methods, results, and conclusions  (for specific guidance, see STARD for Abstracts) | 2 |
|  | **INTRODUCTION** |  |  |  |
|  |  | **3** | Scientific and clinical background, including the intended use and clinical role of the index test | 8-9 |
|  |  | **4** | Study objectives and hypotheses | 9 |
|  | **METHODS** |  |  |  |
|  | *Study design* | **5** | Whether data collection was planned before the index test and reference standard were performed (prospective study) or after (retrospective study) | 10 |
|  | *Participants* | **6** | Eligibility criteria | 10,11 |
|  |  | **7** | On what basis potentially eligible participants were identified  (such as symptoms, results from previous tests, inclusion in registry) | 10 |
|  |  | **8** | Where and when potentially eligible participants were identified (setting, location and dates) | 10,16 |
|  |  | **9** | Whether participants formed a consecutive, random or convenience series | 10 |
|  | *Test methods* | **10a** | Index test, in sufficient detail to allow replication | 11-13 |
|  |  | **10b** | Reference standard, in sufficient detail to allow replication | 13 |
|  |  | **11** | Rationale for choosing the reference standard (if alternatives exist)  Histology is gold standard reference standard. Follow-up for patients managed without surgery was also used | 13 |
|  |  | **12a** | Definition of and rationale for test positivity cut-offs or result categories  of the index test, distinguishing pre-specified from exploratory | 12 |
|  |  | **12b** | Definition of and rationale for test positivity cut-offs or result categories  of the reference standard, distinguishing pre-specified from exploratory | 13 |
|  |  | **13a** | Whether clinical information and reference standard results were available to the performers/readers of the index test | 13 |
|  |  | **13b** | Whether clinical information and index test results were available  to the assessors of the reference standard | NA |
|  | *Analysis* | **14** | Methods for estimating or comparing measures of diagnostic accuracy | 14 |
|  |  | **15** | How indeterminate index test or reference standard results were handled. | 14 |
|  |  | **16** | How missing data on the index test and reference standard were handled | 15 |
|  |  | **17** | Any analyses of variability in diagnostic accuracy, distinguishing pre-specified from exploratory | 11, 15 |
|  |  | **18** | Intended sample size and how it was determined | 15 |
|  | **RESULTS** |  |  |  |
|  | *Participants* | **19** | Flow of participants, using a diagram | Figure 2 |
|  |  | **20** | Baseline demographic and clinical characteristics of participants | Table 1 |
|  |  | **21a** | Distribution of severity of disease in those with the target condition | 16, Table 2 |
|  |  | **21b** | Distribution of alternative diagnoses in those without the target condition | 16-7 |
|  |  | **22** | Time interval and any clinical interventions between index test and reference standard | 13 |
|  | *Test results* | **23** | Cross tabulation of the index test results (or their distribution)  by the results of the reference standard | Table 3 |
|  |  | **24** | Estimates of diagnostic accuracy and their precision (such as 95% confidence intervals) | Table 3 |
|  |  | **25** | Any adverse events from performing the index test or the reference standard | NA |
|  | **DISCUSSION** |  |  |  |
|  |  | **26** | Study limitations, including sources of potential bias, statistical uncertainty, and generalisability | 23 |
|  |  | **27** | Implications for practice, including the intended use and clinical role of the index test | 21-2, 25 |
|  | **OTHER INFORMATION** |  |  |  |
|  |  | **28** | Registration number and name of registry | 3 |
|  |  | **29** | Where the full study protocol can be accessed | 10 |
|  |  | **30** | Sources of funding and other support; role of funders | 6 |

**Supplementary Table 11.** Reporting of a multivariable prediction model for Individual Prognosis Or Diagnosis (TRIPOD) guidelines.

| **Section/Topic** | | | **Checklist Item** | **Page** |
| --- | --- | --- | --- | --- |
| **Title and abstract** | | | | |
| Title | 1 | D;V | Identify the study as developing and/or validating a multivariable prediction model, the target population, and the outcome to be predicted. | 1 |
| Abstract | 2 | D;V | Provide a summary of objectives, study design, setting, participants, sample size, predictors, outcome, statistical analysis, results, and conclusions. | 2 |
| **Introduction** | | | | |
| Background and objectives | 3a | D;V | Explain the medical context (including whether diagnostic or prognostic) and rationale for developing or validating the multivariable prediction model, including references to existing models. | 8-9 |
|  | 3b | D;V | Specify the objectives, including whether the study describes the development or validation of the model or both. | 9 |
| **Methods** | | | | |
| Source of data | 4a | D;V | Describe the study design or source of data (e.g., randomized trial, cohort, or registry data), separately for the development and validation data sets, if applicable. | 10 |
|  | 4b | D;V | Specify the key study dates, including start of accrual; end of accrual; and, if applicable, end of follow-up. | 16 |
| Participants | 5a | D;V | Specify key elements of the study setting (e.g., primary care, secondary care, general population) including number and location of centres. | 10, 16 |
|  | 5b | D;V | Describe eligibility criteria for participants. | 10-11 |
|  | 5c | D;V | Give details of treatments received, if relevant. | na |
| Outcome | 6a | D;V | Clearly define the outcome that is predicted by the prediction model, including how and when assessed. | 13-14 |
|  | 6b | D;V | Report any actions to blind assessment of the outcome to be predicted. | 13 |
| Predictors | 7a | D;V | Clearly define all predictors used in developing or validating the multivariable prediction model, including how and when they were measured. | 11, Supplementary material Appendix C |
|  | 7b | D;V | Report any actions to blind assessment of predictors for the outcome and other predictors. | 13 |
| Sample size | 8 | D;V | Explain how the study size was arrived at. | 15 |
| Missing data | 9 | D;V | Describe how missing data were handled (e.g., complete-case analysis, single imputation, multiple imputation) with details of any imputation method. | 15 |
| Statistical analysis methods | 10c | V | For validation, describe how the predictions were calculated. | 14, Supplementary material Appendix C |
|  | 10d | D;V | Specify all measures used to assess model performance and, if relevant, to compare multiple models. | 14 |
|  | 10e | V | Describe any model updating (e.g., recalibration) arising from the validation, if done. | NA |
| Risk groups | 11 | D;V | Provide details on how risk groups were created, if done. | NA |
| Development vs. validation | 12 | V | For validation, identify any differences from the development data in setting, eligibility criteria, outcome, and predictors. | NA |
| **Results** | | | | |
| Participants | 13a | D;V | Describe the flow of participants through the study, including the number of participants with and without the outcome and, if applicable, a summary of the follow-up time. A diagram may be helpful. | 16, Figure 2 |
|  | 13b | D;V | Describe the characteristics of the participants (basic demographics, clinical features, available predictors), including the number of participants with missing data for predictors and outcome. | Table 1, Supplementary table 1 |
|  | 13c | V | For validation, show a comparison with the development data of the distribution of important variables (demographics, predictors and outcome). | NA |
| Model performance | 16 | D;V | Report performance measures (with CIs) for the prediction model. | Table 3 |
| Model-updating | 17 | V | If done, report the results from any model updating (i.e., model specification, model performance). | NA |
| **Discussion** | | | | |
| Limitations | 18 | D;V | Discuss any limitations of the study (such as nonrepresentative sample, few events per predictor, missing data). | 23 |
| Interpretation | 19a | V | For validation, discuss the results with reference to performance in the development data, and any other validation data. | NA |
|  | 19b | D;V | Give an overall interpretation of the results, considering objectives, limitations, results from similar studies, and other relevant evidence. | 21 |
| Implications | 20 | D;V | Discuss the potential clinical use of the model and implications for future research. | 21-2, 24 |
| **Other information** | | | | |
| Supplementary information | 21 | D;V | Provide information about the availability of supplementary resources, such as study protocol, Web calculator, and data sets. | 10, Supplementary material |
| Funding | 22 | D;V | Give the source of funding and the role of the funders for the present study. | 6 |

### APPENDIX C – Algorithms for Index Test Combinations and Thresholds

#### IOTA simple rules

Many adnexal masses have a typical USG appearance and can therefore be easily correctly classified even by relatively inexperienced USG examiners. IOTA group have established simple rules based on a number of clearly defined USG features that can guide examiners. Using these simple rules no risk estimates are produced, but tumours are classified as benign, malignant or unclassifiable. The simple rules consist of five USG features of malignancy (M-features), and five USG features suggestive of a benign mass (B-features). A mass is classified as malignant if at least one M-feature and none of the B-features are present, and vice versa. If no B- or M-features are present, or if both B- and M-features are present, then the rules are considered inconclusive (unclassifiable mass) and a different diagnostic method should be used. In the ROCkeTS study, inconclusive results were reported, but excluded from any further analysis.

#### B- and M- features.

| **Rules for predicting a malignant tumor (M-rules)** | | | **Rules for predicting a benign tumor (B-rules)** | | |
| --- | --- | --- | --- | --- | --- |
| M1 | Irregular solid tumour | □ | B1 | Unilocular | □ |
| M2 | Presence of ascites | □ | B2 | Presence of solid components where the largest | □ |
| M3 | At least four papillary structures | □ |  | Solid component has a largest diameter < 7 mm |  |
| M4 | Irregular multilocular solid tumour with largest | □ | B3 | Presence of acoustic shadows | □ |
|  | Diameter ≥ 100 mm |  | B4 | Smooth multilocular tumour with largest diameter < 100 mm | □ |
| M5 | Very strong blood flow (colour score 4) | □ | B5 | No blood flow (colour score 1) | □ |

#### ADNEX

The ADNEX model uses the following predictors, with measurement unit between parentheses and reference letter between square brackets:

[A] patient age (years)

[B] serum CA125 (U/mL)

[C] maximum diameter of lesion (mm)

[D] maximum diameter of largest solid part (mm)

[E] number of cyst locules (>10=1, <10=0)

[F] number of papillations (0 papillary structure=0, 1 papillary structure=1, 2 papillary structures=2, and 3 papillary structures=3 and 4+ papillary structures=4)

[G] presence of acoustic shadows (yes=1, no=0)

[H] presence of ascites (yes=1, no=0)

[I] centre (cancer centre=1, otherwise=0)

The linear predictors z1 to z4 contain model coefficients for each predictor for the prediction of borderline vs. benign tumours (z1), stage I cancer vs. benign tumours (z2), stage II-IV cancer vs. benign tumours (z3), and secondary metastatic cancer vs. benign tumours (z4). The linear predictors are as follows:

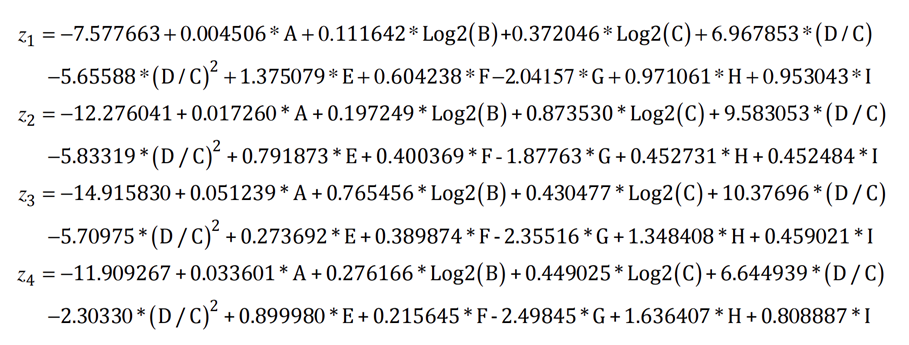

The probabilities of benign tumour P(Benign) is computed as follows:

$$P\left( Benign \right)=\frac{1}{1+\exp\left( z_{1} \right)+\exp\left( z_{2} \right)+\exp\left( z_{3} \right)+exp(z_{4})}$$

Thus, the probability of a malignancy is $1-P(\mathrm{Benign})$

IOTA ADNEX model is integrated into ultrasound machines from GE Healthcare USA/Austria, Samsung Healthcare Seoul South Korea, Canon Japan and Mindray China. IOTA ADNEX model is also available through Gynaia. https://gynaia.com/

#### Components and predictors used in the index test combinations.

| **Component** | **Predictors**  ***Unit or result*** | **RMI 1** | **ROMA** | **IOTA ADNEX** | **IOTA simple rules** | **IOTA sRisk model** | **CA125** |
| --- | --- | --- | --- | --- | --- | --- | --- |
| Patient Baseline CRF | Age *Years* |  |  | X |  |  |  |
|  | Menopausal status  *Ordinal: Pre=1, Post=3* | X |  |  |  |  |  |
|  | Mode of presentation  *Dichotomous: 1=Referral from cancer unit, 0=other* |  |  | X |  | X |  |
| Blood sample | Serum CA125 *U/ml* | X | X | X |  |  | X |
|  | HE4 *pmol/l* |  | X |  |  |  |  |
| USG Scan | Ultrasound (U) score  *Ordinal score of 0 for a U score of 0, 1 for a U score of 1 and 3 for a U score of 2-5.* | X |  |  |  |  |  |
|  | Maximum diameter of lesion *mm* |  |  | X |  |  |  |
|  | Maximum diameter of largest solid part *mm* |  |  | X |  |  |  |
|  | Number of cyst locules  *Dichotomous: 1=>10, 0=≤10* |  |  | X |  |  |  |
|  | Number of papillations  *Ordinal score of 0 for 0 papillary structure, 1 for 1 papillary structure, 2 for 2 papillary structures and 3 for 3 papillary structures and 4 for 4+ papillary structures.* |  |  | X |  |  |  |
|  | Presence of acoustic shadows  *Dichotomous: 1=Yes, 0=No* |  |  | X |  |  |  |
|  | Presence of ascites  *Dichotomous: 1=Yes, 0=No* |  |  | X |  |  |  |
|  | M1: Irregular solid mass  *Dichotomous: 1=Yes, 0=No* |  |  |  | X | X |  |
|  | M2: Presence of ascites  *Dichotomous: 1=Yes, 0=No* |  |  |  | X | X |  |
|  | M3: At least four papillary structures  *Dichotomous: 1=Yes, 0=No* |  |  |  | X | X |  |
|  | M4: Irregular multilocular solid tumour with largest diameter ≥ 100mm *Dichotomous: 1=Yes, 0=No* |  |  |  | X | X |  |
|  | M5: Very strong blood flow (colour score 4) *Dichotomous: 1=Yes, 0=No* |  |  |  | X | X |  |
|  | B1: Unilocular cyst  *Dichotomous: 1=Yes, 0=No* |  |  |  | X | X |  |
|  | B2: Presence of solid components where the largest solid component has a largest diameter ≤7mm  *Dichotomous: 1=Yes, 0=No* |  |  |  | X | X |  |
|  | B3: Presence of acoustic shadows  *Dichotomous: 1=Yes, 0=No* |  |  |  | X | X |  |
|  | B4: Smooth multilocular tumour with largest diameter <100mm  *Dichotomous: 1=Yes, 0=No* |  |  |  | X | X |  |
|  | B5: No blood flow (colour score 1)  *Dichotomous: 1=Yes, 0=No* |  |  |  | X | X |  |

#### Additional index test combinations.

| **Other index test algorithms** | **Detail** |
| --- | --- |
| **RMI 1**  ***U*** × ***M*** × **CA125** | **Ultrasound (U)**: where a total ultrasound score of 0 made U=0, a score of 1 made U=1, and a score of 2-5 made U=3  **Menopausal status (M):** premenopausal status made M=1 and postmenopausal status made M=3  **Serum CA125:** CA125 U/ml applied directly to the calculation |
| **ROMA**  **Premenopausal:** Predictive Index (PI) = −12.0 + (2.38 × LN(**HE4**)) + (0.0626 × LN(**CA125**))  **Postmenopausal:** PI = −8.09 + (1.04 × LN(**HE4**)) + 0.732 × (LN(**CA125**))  **Predicted probability** = exp(PI)/[1 + exp(PI)] × 100 | **HE4** in units pmol/l  **Serum CA125** in units U/ml |
| **IOTA sRisk Model**  Predictive Index= −0.97 − 3.41(**B1**) − 2.25(**B2**) – 1.66 (**B3**) − 2.75(**B4**) – 1.86(**B5**) + 2.19(**M1**) + 2.65(**M2**) + 1.53(**M3**) + 0.98(**M4**) + 1.55(**M5**) + 0.92(**C**)  **Predicted probability** = exp(PI)/[1 + exp(PI)] × 100 | **B1-B5** and **M1-M5** are respectively the five USG features of malignancy and benign mass that form the simple rules. These are dichotomous variables where Yes=1 and 0=No.  **Centre (C)**: cancer unit made C=1 and otherwise made C=0. |
| **CA125**  No calculation | **Serum CA125** in units U/ml, threshold applied directly to the index test. |

#### ORADS

Calculation of ORADS lexicon score from IOTA variables recorded on the Ultrasound Case report form, as previously described in Timmerman S, et al. doi: 10.1001/jamaoncol.2022.5969. PMID: 36520422; PMCID: PMC9856950. Threshold - ORADS 4 and above correspond to $\geq$10% risk of malignancy.

| **O-RADS Lexicon** | **IOTA Terms / Variables corresponding to the O-RADS lexicon** |
| --- | --- |
| **O-RADS 2** | |
| 2a: Simple cyst | Locality = unilocular; irregular = no; echogenicity = anechoic; lesion largest diameter <10 cm |
| 2a1: ≤3 cm | Lesion largest diameter ≤3 cm |
| 2a2: >3 – 5 cm | Lesion largest diameter > to ≤5 cm |
| 2a3: > 5 but < 10 cm | Lesion largest diameter >5 to <10 cm |
| **2b: Classic benign lesions** | |
| 2b1: Typical hemorrhagic cyst <10 cm | Subjective assessment: hemorrhagic cyst, probably or certainly benign; lesion largest diameter <10 cm |
| 2b2: Typical dermoid cyst <10 cm | Subjective assessment: dermoid cyst, probably or certainly benign; lesion largest diameter <10 cm |
| 2b3: Typical endometrioma <10 cm | Subjective assessment: endometrioma, probably or certainly benign; lesion largest diameter <10 cm |
| Peritoneal inclusion cyst | Subjective assessment: peritoneal inclusion cyst, probably or certainly benign |
| Hydrosalpinx | Subjective assessment: hydrosalpinx, probably or certainly benign |
| 2c: Nonsimple unilocular cyst, smooth inner margin | Locality = unilocular; irregular = no; lesion largest diameter <10 cm |
| 2c1: ≤3 cm | Lesion largest diameter ≤3 cm |
| 2c2: >3 but <10 cm | Lesion largest diameter >3 to <10 cm |
| **O-RADS 3** | |
| 3a: Unilocular cyst ≥10 cm (simple or nonsimple) | Locality = unilocular; irregular = no; lesion largest diameter ≥10 cm |
| 3b: Typical dermoid cysts, endometrioma, hemorrhagic cysts ≥10 cm | |
| 3b1: Typical hemorrhagic cyst ≥10 cm | Subjective assessment: hemorrhagic cyst, probably or certainly benign; lesion largest diameter ≥10 cm |
| 3b2: Typical dermoid cyst ≥10 cm | Subjective assessment: dermoid cyst, probably or certainly benign; lesion largest diameter ≥10 cm |
| 3b3: Typical endometrioma ≥10 cm | Subjective assessment: endometrioma, probably or certainly benign; lesion largest diameter ≥10 cm |
| 3c: Unilocular cyst, any size with irregular wall <3 mm height | Locality = unilocular; irregular = yes |
| 3d: Multilocular cyst <10 cm, smooth inner wall, color score 1–3 | Locality = multilocular; irregular = no; color score < 4; lesion largest diameter <10 cm |
| 3e: Solid smooth, any size, color score = 1 | Locality = solid; irregular = no; color score = 1 |
| **O-RADS 4** | |
| 4a: Multilocular cyst, no solid component | |
| 4a1: ≥10 cm, smooth inner wall, color score 1–3 | Locality = multilocular; irregular = no; color score < 4; lesion largest diameter ≥10 cm |
| 4a2: Any size, smooth inner wall, color score = 4 | Locality = multilocular; irregular = no; color score = 4 |
| 4a3: Any size; irregular inner wall, irregular septation, or both; color score = any | Locality = multilocular; irregular = yes |
| 4b: Unilocular cyst with solid component, any size, 0-3 papillary projections, color score = any | Locality = unilocular, solid; No. of papillary projections <4 |
| 4c: Multilocular cyst with solid component, any size, color score = 1–2 | Locality = multilocular, solid; color score <3 |
| 4d: Solid smooth, any size, color score = 2–3 | Locality = solid; irregular = no; color score = 2 or 3 |
| **O-RADS 5** | |
| 5a: Unilocular cyst, any size, ≥4 papillary projections, color score = any | Locality = unilocular, solid; No. of papillary projections ≥4 |
| 5b: Multilocular cyst with solid component, any size, color score = 3–4 | Locality = multilocular, solid; color score >2 |
| 5c: Solid smooth, any size, color score = 4 | Locality = solid; irregular = no; color score = 4 |
| 5d: Solid irregular, any size, color score = any | Locality = solid; irregular = yes |
| 5e: Ascites, peritoneal nodules, or both | Ascites = yes or metastases = yes |

### APPENDIX D – ROCKeTs Laboratory Manual

Laboratory Sample Handling Manual

**Staff Responsibilities**

All personnel involved in taking blood samples for the ROCkeTS study must sign the site *Trial Signature & Delegation Log* kept by the Principal Investigator in the ROCkeTS Site File. Personnel involved in obtaining blood samples as part of routine care do not need to sign this log.

**Sample Collection Schedule**

For ROCkeTS, blood samples need to be taken after consent when the participant is registered in to the study. This is the only time point for collection.

**Serum:** Draw a sufficient amount of blood to yield the necessary serum volume. Invert tube 5-10 times to activate clotting. Allow blood to clot at room temperature for 30 minutes. NOTE: Avoid hemolysis.

**Specimen Collection Tubes**

- Red-top tube, plastic
- This tube is a plastic Vacutainer containing a clot activator but no anticoagulants, preservatives, or separator material. It is used for collection of serum for selected laboratory tests as indicated.
- Invert the tube to activate the clotting; let stand for 30-60 minutes before centrifuging.
- Centrifuge the blood sample at the end of the clotting time (30-60 minutes) in a horizontal rotor (swing-out head) for ≤1300 RCF for 10 minutes at room temperature.
- If the blood is not centrifuged immediately after the clotting time (30 to 60 minutes at room temperature), the tubes should be refrigerated (4°C) for no longer than 4 hours.
- PIPETTE off serum into 1 ml labelled plastic cryovials and freeze (fill 5 cryovials with serum, if 5 cryovials is not attainable a minimum of 3 cryovials is required). Do not freeze Vacutainer® tubes. Please ensure you do not pipette red blood cells – this can be done by leaving a small amount of serum in the tube. This process should be completed within 1 hour of centrifugation.
- Place all aliquots upright in a specimen box or rack in an -40°C or colder freezer. All specimens should remain at -40°C or colder prior to shipping. The samples should not be thawed prior to shipping.
- Samples will be transported to the NHS Gateshead labs, via a courier organised by the BCTU.

**SPECIMEN COLLECTION AND PREPARATION FOR ANALYSIS**

**Specimen Conditions**

Do not use specimens with the following conditions:

Heat-inactivated

Pooled

Grossly hemolyzed

Obvious microbial contamination

For accurate results, serum specimens should be free of fibrin, red blood cells, and other particulate matter. Serum specimens from patients receiving anticoagulant or thrombolytic therapy may contain fibrin due to incomplete clot formation.

Use caution when handling patient specimens to prevent cross contamination.

Use of disposable pipettes or pipette tips is recommended.

For optimal results, inspect all specimens for bubbles. Remove bubbles with an applicator stick before analysis. Use a new applicator stick for each specimen to prevent cross contamination. Complete the label provided with following information and label tube:

Patient study number

Patient initials

Date sample taken

Vial number i.e. 1, 2, 3 etc. Ensure the Sample log is completed (stored in the site file) with the following information:

Date and time blood sample taken

Date and time serum sample was frozen

Serum stored in vial box number

Date collected by courier

Any notes about the sample

Serum must be transported on dry ice.
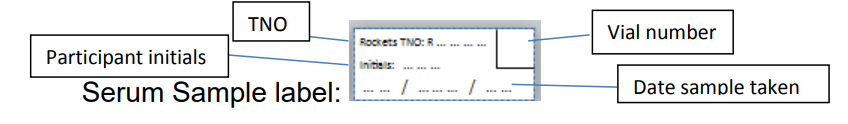

**Consumables:**

Consumable requests should be sent to ROCkeTS consumables listed below. Please leave plenty of time for resupply. Remember to quote the trial ID (rockets) in all correspondence.

**Envelope label**

Rockets Consumables

BCTU

Public Health Building

University of Birmingham

Vincent Drive

Birmingham

B15 2TT

### APPENDIX E – ROCkeTS collaborators

**Regional Study Centre Team**

**Belfast City Hospital**

Nagar H (PI & imaging lead), McAlister C, Clarke, P, O’Donnell A, Cunningham W, McAllister J, McClisker S, McClean S, Dadebo B, Laverly G

**Birmingham City Hospital**

Sundar S (PI), Parker R (Associate PI), Khan H (Imaging lead), Butler L, Gammon B, Samuel-Oparah U, Orme A, Marsden L, Smith G, Cartwright J, Storistreet D, Goddard H, Williams A, Bruten E, Devonport D, Pilsworth Z

**Birmingham Women's Hospital**

Abedin P (PI), Balogun M (Imaging lead), McCooty S, Qureshi N, Chana P, Beale F, Galloway A, Iqbal G, Carden N, McPake C

**East Surrey Hospital**

Jermy K (PI & Imaging lead), Weller S, Maher S, Summers G, Nicks H, Knight H, Habibi R

**Guy's Hospital**

Sayasheh A (PI and Imaging lead), Abdelbar A (Associate PI), Debattista L, D'Alessandro V, Bilbert-Jones H, Khaula M, Ijeomah-Orgi M, Worthington M, Fitzpatrick-Greening M, Lombardi S, Ng L, Shipa B, Zielonka A, Jadhau A, Barrett S, Love R, Borley J, Mohamoud N

**Hinchingbrooke Hospital**

Majmudar T (PI & Imaging lead), Mackenzie C (Associate PI), Palombo C, Baker TA, Adebayo A, Wilde L, Nosib H, Miller S, Webb D, Perkins L, Plaza S, Goss V, Donnelly S, Osmanska A, Kurian R, Lam R, Calcada R, Marco-Illana E

**James Cook University Hospital**

Hebblethwaite N (PI), Exley K (Imaging lead), Peatman S, Kane J, Hebbron K, Alexander H, Harwood H, Cuthbert H, Hodges M, Mcgajjar J, Wright L, Dale M, Chadwick V, Naseem S, Iqbal N, Proctor C

**Liverpool Women's Hospital**

McDonald RD (PI), Hamer M (Imaging lead), Robinson-Jones A, Pearritt S, Corlett P, Wray J, Drury J, Heathcote L, Sutton V, Coppin D, Cooke K, Bolderson J, Bia C, Sawan S, Davies M, Lowe A, Hamlett H, Houghton F, Beasley A, Robinson-Jones A, Rice E, Bell S

**Norfolk & Norwich University Hospital**

Duncan T (PI), Ames V (Imaging lead), Archer D, Gibbins T, Turner S, Nieto J, Borbos N, Turnbull H, Anderson S, French K, Hunter N, High L, Dann A, Licence V, Websdale C, Darby H, Malone E, Walton S, Schofield E, Platt J, Cooper A, Cook J, Cornwell M, Ashgar M, Walter S, Macnab W, Kellett J, Halliwell-Bass S, Knapp S, McElhinney S

**Northampton General Hospital**

Gnanachandran C (PI & Imaging lead), Alawad H (Associate PI), Kariyadil B, Jose S, Kempa A, Woolhouse C, Duncan A, Bussey R, Campey L, Hall K, Dudgeon L, Hitchcock R, Polnik M, Stockham LJ, Al Husain H, Grantham G

**Nottingham City Hospital**

son K (PI & Imaging lead), Coleridge S/ Naskretski A (Associate PI), Dennis S, Gibbins T, Williamson K, Nunns D, Abu J, Hammond R, Juliana A, Golding J, Cope J, Mills S, Gan C, Wrigmy S, Warren C, Ward H, Wilson G

**Peterborough General Hosptial**

Ramsay B (PI), Moshy R (Imaging lead), Adebayo A, Palombo C, Woodhouse S, Barter E, Baker TA, Butcher D, Goodyear P, O’Sullivan S, O’Herlihy S, Collins H, Sidlow J, Weatherburn A, Steachan S, Diaz S, Austin M, Penart-Buck F, Dunn S, Adams L, Bhayani J

**Princess Anne Hospital**

Rosello N (PI), Johnson S (Imaging lead), Benson L, Wood J, Lowry J, Smith L, Barton S

**Queen Elizabeth Hospital, Gateshead**

Hughes T (PI & Imaging lead), Pearce L, McCormick W

**Royal Blackburn Hospital**

Willett M (PI)

**Royal Hallamshire Hospital**

Abdi S (PI & Imaging lead), Duffy S, Bullivant E, Taylor F, Waller C, Jobling N, Tidy J, Palmer J, Gillespie A, Senbeto S, Sutcliffe A, Johnson K, Murtagh L, Lally B

**Royal Victoria Infirmary**

Russell M (PI), Maddison J (Imaging lead), Kimber A, Graham J, Conner D, Murtha V, Dunn E, Lim CP, Russell M, Chalhoub T, Birtles JO, Davies M, Galeon MC, Lowes J, Narayansingh G, Fenn A, Gallgher I, Brown K, Hoh J

**The Royal London Hospital St Bartholomew's Hospital**

Manchanda R (PI & Imaging lead), Aswat S, Robbani S, Dzumbunu F, Chandrasekaran D, Gaba F, Lawrence A, Sahdev A, Hillman P

**Royal Sussex County Hospital**

Kaushik S (PI), Baron S (Associate PI), Vitta L (Imaging lead), Herbertson R, Lyttle AJ, Laltho R, Larsen-Disney P, Newman N, Curry J, Heron H, Porges A, McLennan C, Frattaroli p, Temegan J, O’Neill F, Whitfield C, Lavender P, , Dailey N, Drews F, Langford K, Fellich V

**Royal Preston Hospital**

Keating P/Wood N (PI), Butcher T (Imaging lead), Young A, Cornthwaite S, Swan A, Martyniak A, Brunton M, Sutton V, Turner E, Ellel K, Antrobus P, Leach M, Musa N, Ardern N, Prashar S, Jones K, Brar N, Cook A, Patel S, Gardner A, Panchal K, Speirs R

**University Hospital of North Durham**

Sengupta P (PI), Kent R (Imaging lead), Deur J, Downey L, Sen S, Atkinson V, Bodnar S, wook M, Walton E, Arava U, Rathaparchi S, Damigos E, Kay A, Potts K, Chatt R, Jennings J, Baggett A, Beukenholdt R, Bainbridge V, Clark S, Nemeth Z, Humphries C, Stamp K, Brown E, O’Brien J, Hobson S

**Sheffield Teaching Hospitals**

Palmer J (Imaging lead)

**University Hospital of Wales**

Sharma A (PI), Sinha A (Imaging lead), Noble N, Wadmore C, Holland R, Pugh N, Lim K, Rzyska E, Shamsunsin L, Gnanachandrana C, Kendall J, Price C, Cloudsdale R, McNee D, Smith C, Heirene L, James R

**Walsall Manor Hospital**

Ghazal F (PI), Rai H (Imaging lead), Davies J, Mhembere P, Botfield L, Fletcher J, Darby S, Lenehan F, Richardson L, Thomas T, Hannon J, Cogings P

**Watford General Hospital**

Radhikavikram S (PI), Malhotra V (Imaging lead), Walker E, Markwell K, Zhao X

### APPENDIX F – Statistical Analysis Plan

Refining Ovarian Cancer Test Accuracy Scores: A test accuracy study to validate new risk scores in women with symptoms of suspected ovarian cancer protocol

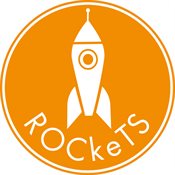

Trial Registration: ISRCTN **17160843**

#### Statistical Analysis Plan

| SAP Version Number | Protocol Version Number |
| --- | --- |
| 0.1 | 10.0 |

| Name of Author: | Ridhi Agarwal | Role: | Trial Statistician | Affiliation: | Institute of Applied Health Research  University of Birmingham |
| --- | --- | --- | --- | --- | --- |
| Signature of Author: | 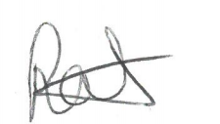 | Date: | 19/05/2022 |  |  |
| Name of Reviewer: | Yemisi Takwoingi on behalf of Jon Deeks | Role: | Senior Statistician | Affiliation: | Institute of Applied Health Research  University of Birmingham |
| Signature of Reviewer: | 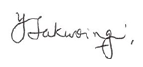 | Date: | 19/05/2022 |  |  |
| Name of Chief Investigator: | Sudha Sundar | Role: | Chief Investigator | Affiliation: |  |
| Signature of Chief Investigator: | 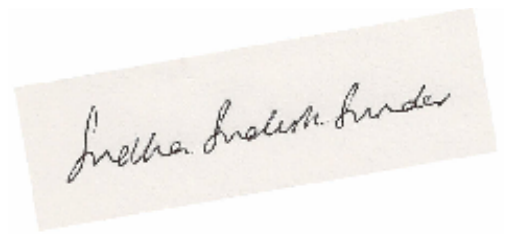 | Date: | 19/05/2022 |  |  |
| **This Statistical Analysis Plan has been approved by:** | | | | | |
| Name of Approver: | Yemisi Takwoingi on behalf of Jon Deeks | Role: | Senior Statistician | Affiliation: | Institute of Applied Health Research  University of Birmingham |
| Signature of Approver: | 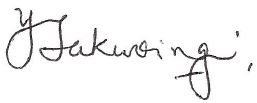 | Date: | 19/05/2022 |  |  |

Statistical Analysis Plan (SAP) Amendments.

| **SAP version number** | **SAP section number** | **Description of and reason for change** | **Timing of change with respect to interim analysis/ final analysis/ database lock** | **Blind Reviewer** |
| --- | --- | --- | --- | --- |
| 1.0 |  |  |  | Name: |
|  |  |  |  | Signature: |
|  |  |  |  | Date: |
|  |  |  |  | Name: |
|  |  |  |  | Signature: |
|  |  |  |  | Date: |
|  |  |  |  | Name: |
|  |  |  |  | Signature: |
|  |  |  |  | Date: |

Statistical analysis plan abbreviations and definitions.

| **Abbreviations & Definitions** | |
| --- | --- |
| **Abbreviation / Acronym** | **Meaning** |
| BCTU | Birmingham Clinical Trials Unit |
| CONSORT | Consolidated Standards of Reporting Trials |
| DMC | Data Monitoring Committee |
| FDD | Final Definitive Diagnosis |
| FNAC | Fine Needle Aspiration Cytology |
| ISRCTN | International Standard Randomised Controlled Trial Number |
| ITT | Intention to Test |
| MDT | Multidisciplinary Team |
| RTE | Shear and strain wave elastography |
| SAE | Serious Adverse Event |
| SAP | Statistical Analysis Plan |
| SUSAR | Suspected Unexpected Serious Adverse Reaction |
| TSC | Trial Steering Committee |
| US | Ultrasound |
| **Term** | **Definition** |
| International Standard Randomised Controlled Trial Number | A clinical trial registry |
| Protocol | Document that details the rationale, objectives, design, methodology and statistical considerations of the study |
| Randomisation | The process of assigning trial subjects to intervention or control groups using an element of chance to determine the assignments in order to reduce bias. |
| Statistical Analysis Plan | Pre-specified statistical methodology documented for the trial, either in the protocol or in a separate document. |

#### 1. Introduction

This document is the Statistical Analysis Plan (SAP) for the ROCkeTS study, and should be read in conjunction with the current trial protocol. This SAP details the proposed analyses and presentation of the data for the main paper(s) reporting the results for the ROCkeTS study.

The results reported in these papers will follow the strategy set out here. Subsequent analyses of a more exploratory nature will not be bound by this strategy, though they are expected to follow the broad principles laid down here. The principles are not intended to curtail exploratory analysis (e.g. to decide cut-points for categorisation of continuous variables), nor to prohibit accepted practices (e.g. transformation of data prior to analysis), but they are intended to establish rules that will be followed, as closely as possible, when analysing and reporting data.

Any deviations from this SAP will be described and justified in the final report or publication of the trial (Supplementary Table 30). The analysis will be carried out by an appropriately qualified statistician, who should ensure integrity of the data during their data cleaning processes.

#### 2. Background and rationale

The background and rationale for the study are outlined in detail in the protocol. In brief, ovarian cancer (OC) has an annual incidence of 6500 women and causes 4400 deaths in the UK; the lifetime risk of developing OC is 1 in 54.1 OC is predominantly a disease of older, postmenopausal women, however 1000 women under 50 will be diagnosed with OC annually.^1^

National Institute for Health and Care Excellence (NICE) guidelines, in 2011 recommended sequential testing using serum Ca125 followed by pelvic ultrasound (USG) in women (particularly aged >50) presenting to primary care with symptoms suggestive of OC. However the symptoms are very common^2,3^ with abdominal bloating alone being documented in 16-30% of women presenting to General Practitioners (GPs).^4^ Diagnostic challenges are considerable given (1) the low incidence of OC (a GP sees a woman with OC once in 3-5 years) (2) the low positive predictive value (PPV) of symptoms (only 1 in 400-600 symptomatic women have OC)^5,6^ and (3) the lack of clear diagnostic pathways. Furthermore, NICE guidelines do not detail what USG abnormalities should prompt referral.

Use of this NICE guidance is extremely variable. A survey of 258 GPs report that the majority would refer patients on the basis of raised Ca125 even if the USG was normal.^7^ An audit at City Hospital, Birmingham of 448 referrals through the 2 week wait clinics, reveals that 90% of referrals for suspected OC do not follow guidance.^8^ Referrals were heterogeneous in symptoms, in what the GPs considered were abnormal levels of Ca125 or abnormal USG. Two thirds of women referred were premenopausal. Ovarian masses are common in premenopausal women and up to 10% of women will undergo surgery during their lifetime for the presence of an ovarian mass.^8^

There is substantial literature on tests and scoring systems in women with ovarian masses presenting to secondary care. However, this literature is often derived in women with advanced stage cancer. Literature is limited in studies of women with symptoms presenting to primary care, and no published studies of blood tests or USG in women with symptoms in primary care exist.

It is important to stratify patients into premenopausal and postmenopausal women. Whilst the risk of OC is higher in postmenopausal women, two thirds of patients referred through the rapid access pathway are premenopausal women. Current recommended best practice for premenopausal women (Royal College of Obstetricians and Gynaecologists (RCOG) guidance)^8^ with a complex ovarian mass is to assess risk of malignancy using the Risk of Malignancy Index (RMI), 200 threshold, even though the score was derived for postmenopausal women and the use of logistic regression models in a different population is flawed.

#### 3. Study aim

- To externally validate risk prediction models that estimate the probability of having OC for postmenopausal and premenopausal women with symptoms suggestive of OC.
- To identify optimal risk thresholds for the models that can guide patient management.

#### 4. Trial methods

##### 4.1. Trial design

ROCkeTS is a single arm prospective cohort diagnostic accuracy study to evaluate existing diagnostic tools (risk prediction models and scores) for diagnosing postmenopausal and pre-menopausal women with ovarian cancer. For premenopausal women, ROCkeTS has now been amended in protocol v5 with recruitment focussed on women undergoing surgery/biopsy for suspected OC or adnexal mass due to low prevalence of true positives in premenopausal women referred through rapid access clinics. See SAP Appendix B for study schema.

Postmenopausal and premenopausal women referred to secondary care with symptoms of suspected ovarian are recruited from 20-30 sites within the UK. Additional requirement for premenopausal women is they must be undergoing surgery/biopsy for adnexal mass. Patients referred as outpatients; either as 2-week referrals, USG clinics, routine GP referrals or cross specialty referrals. Patients referred as inpatients through emergency presentations to secondary care.

##### 4.2. Trial tests

The reference standard will be histology of tissues taken from patients who proceed to surgery/biopsy or assessment of wellbeing using a structured questionnaire as follow-up at 12 months after presentation for patients who do not undergo surgery. This does not apply to premenopausal women recruited following amendment in protocol v5 as they will all undergo surgery/biopsy for suspected ovarian cancer or adnexal mass.

The patient data includes symptom questionnaire, blood sample and USG scan, which will be used in the diagnostic tools identified in Phases 1 and 2 of the ROCkeTS study for diagnosing postmenopausal and premenopausal women with OC. The diagnostic tools are split into two lists of clinical interest and academic interest.

Clinical list:

- RMI 1 (Thresholds: 200, 250)
- ROMA (Thresholds: 13.1% for premenopausal and 27.7% for postmenopausal, 12.5% for premenopausal and 14.4% for postmenopausal, 7.4% for premenopausal and 25.3% for postmenopausal, 11.4% for premenopausal and 29.9% for postmenopausal)
- UCL model
- ADNEX (Primary threshold: 10%, secondary threshold: 3%)
- IOTA simple rules (see SAP Appendix C for more details)

Academic list:

- IOTA sRisk model (Primary threshold: 10%, secondary threshold: 3%)
- CA125 (Thresholds: 87 IU/ml for premenopausal and 35 IU/ml for postmenopausal)

##### 4.3. Primary outcome measure

Key outcome measures are the accuracy of the diagnostic tools in terms of their discrimination ability (e.g. sensitivity, specificity) and calibration (observed versus predicted probabilities) for diagnosing primary invasive ovarian malignant neoplasms versus benign, and the identification of thresholds to guide patient management decisions.

##### 4.4. Secondary outcome measures

The accuracy of the diagnostic tools as a binary outcome of:

- Primary invasive and secondary ovarian malignant neoplasms versus benign (borderline will be considered with benign).
- Primary invasive, secondary and borderline ovarian malignant neoplasms versus benign.
- Primary invasive, secondary and borderline ovarian malignant neoplasms versus benign in the subset of patients that were assessed in one-stop clinics.
- Primary invasive, secondary and borderline ovarian malignant neoplasms versus benign where ultrasound was performed by those who had passed IOTA Quality assurance.

##### 4.5. Timing of outcome assessments

The schedule of trial procedures and CRF completion is given in SAP Appendix A.

##### 4.6. Randomisation

N/A

##### 4.7. Sample size

Due to the expected difference in performance in pre and postmenopausal women, separate sample sizes have been computed.

For postmenopausal women: Performance of RMI 1 is assumed to be 70% sensitive and 90% specific. A sample size of n=1299 will have 90% power to detect a 13% difference in sensitivity between the preferred ROCkeTS test (83% sensitivity) and current practice test: RMI 1 at threshold 250 (70% sensitivity) at the 5% significance level, allowing for a moderate positive correlation between test errors of 65%.

For premenopausal women: Performance of RMI 1 is assumed to be sensitivity of 72% and a specificity of 46%. To assess the specificity of the index tests with RMI 1, a sample size of 575 (546 before 5% drop-outs) women will have 90% power to detect at the 5% significance level a difference of 10% from a baseline of 46% specificity using the Alonzo method, assuming uncorrelated test errors and prevalence of 2.5%.

To compare the sensitivity of the index tests with RMI 1, recruitment of premenopausal women will now focus on those more likely to have OC – ie those on the surgical pathway until we have 105 (accounting for 5% drop-outs) women identified as having OC (a minimum of 100 events has been suggested for external validation of prognostic models9). At a prevalence of 14.9%, we predict that we will require 705 patients. However, we only expect 80% of recruits to undergo surgery, which reduces prevalence to 11.9%, meaning we will require a maximum of 880 patients.

##### 4.8. Framework

The objective of this study is to assess the accuracy of the diagnostic tools for diagnosing premenopausal and postmenopausal women with OC. This study will externally validate the diagnostic tools identified from Phases 1 and 2 of the ROCkeTS project on newly presenting premenopausal and postmenopausal patients with symptoms of OC.

##### 4.9. Interim analyses and stopping guidance

N/A

##### 4.10. Internal Pilot Progression Rules

N/A

##### 4.11. Timing of final analysis

The analysis for post-menopausal group will occur once:

- participants that do not undergo surgery have been followed up at a minimum of 12 months
- or participants that undergo surgery/biopsy have their histology results recorded
- and all patient data has been entered onto the trial database and validated as being ready for analysis.

The analysis for pre-menopausal group will occur once:

- participants that do not undergo surgery (pre-menopausal women recruited before amendment in protocol v5) have been followed up at a minimum of 12 months
- participants that undergo surgery/biopsy (pre-menopausal women recruited following amendment in protocol v5) have their histology results recorded
- and all patient data has been entered onto the trial database and validated as being ready for analysis.

##### 4.12. Timing of other analyses

N/A

##### 4.13. Study comparisons

The diagnostic performance of the following diagnostic tools will be compared against that of the comparator test – the existing standard risk prediction score RMI 1 at a threshold of 250.

- RMI 1 at a threshold of 200
- ROMA (Thresholds: 13.1% for premenopausal and 27.7% for postmenopausal, 12.5% for premenopausal and 14.4% for postmenopausal, 7.4% for premenopausal and 25.3% for postmenopausal, 11.4% for premenopausal and 29.9% for postmenopausal)
- ADNEX (threshold: 3%, 10%)
- IOTA simple rules (see SAP Appendix C for more details)
- IOTA sRisk model (Primary threshold: 10%, secondary threshold: 3%)
- CA125 (Thresholds: 87 IU/ml for premenopausal and 35 IU/ml for postmenopausal)

#### 5. Statistical Principles

##### 5.1. Confidence intervals and p-values

All confidence intervals presented will be 95% and two-sided. All applicable statistical tests will be two-sided and p-values will be reported.

##### 5.2. Adjustments for multiplicity

Adjustment for multiple testing will be made using the Bonferroni correction.

##### 5.3. Analysis populations

The analysis will be based on per protocol. Only participants who fulfil the inclusion and exclusion criteria (Section 3.2 of the protocol) and the definition of adherence (Section 5.4) will be used.

##### 5.4. Definition of adherence

Where histology is unavailable, participants should be followed up for the full 12 months.

The patient data outlined in Section 5.1 of protocol should be collected within three months of whichever of the following occurred first: presentation or at the point the first set of data was collected.

If a USG recording IOTA variables has been performed within 120 days prior to the surgery (if surgery is performed) then that scan can be used to provide information for ROCkeTS.

##### 5.5. Handing protocol deviations

A protocol deviation is defined as a failure to adhere to the protocol such as errors in applying the inclusion/exclusion criteria, the incorrect test being given, incorrect data being collected or measured, follow-up visits outside the visit window or missed follow-up visits. A strict definition of per protocol principle will be applied and will include all participants as per protocol population described in Sections 5.3 and 5.4 of the SAP. This does not include those participants who have specifically withdrawn consent for the use of their data in the first instance.

##### 5.6. Unblinding

This is an unblinded study.

#### 6. Trial population

##### 6.1. Recruitment

The number of patients screened, recruited and ineligible will be reported. The number of patients recruited will be summarised overall and by site. A STARD flow diagram will be produced to describe the participant flow through the study in the premenopausal and postmenopausal groups. This will include information on the number (with reasons) of losses to follow-up, withdrawals and participants excluded/ineligible.

##### 6.2. Baseline characteristics

The premenopausal and postmenopausal groups will be described in terms of demographics and clinical characteristics separately and by presence/absence of OC, where the latter will be defined in two ways. Firstly, according to primary outcome of primary invasive malignant neoplasms versus benign in Surgery CRF and secondly, by Outcome CRF with the question if the participant has been diagnosed with ovarian cancer in the last 12 months. Categorical data will be summarised by number of participants and percentages. Continuous data will be summarised by the mean and standard deviation if deemed to be normally distributed or median and interquartile range if data appears skewed. The number of patients with missing data will also be tabulated.

The characteristics reported will include, but not limited to:

- Tabulation of mode of presentation against reason for referral (available only for GP referrals in registration form).
- Tabulation of the diagnostic category (found in surgery CRF) against mode of presentation.
- Tabulation of the symptoms questionnaire and its severity.
- Summary of the serum tumour markers (After entry into Rockets in Registration form).

#### 7. Test(s)

##### 7.1. Description of the test(s)

The index tests and reference standard used are described in Section 4.2 of this document and in Appendix E.

##### 7.2. Adherence to allocated test

A tabulation of the adherence stated in section 5.4 will be produced by postmenopausal and premenopausal groups (proportions and percentages) and by recruiting site.

#### 8. Protocol deviations and violations

Numbers and percentages by postmenopausal and premenopausal groups will be tabulated for the protocol deviations and violations.

#### 9. Analysis methods

##### 9.1. Covariate adjustment

The accuracy of the diagnostic tools identified in Section 4.2 will be assessed for diagnosing postmenopausal and premenopausal women with OC. Details of the patient data that will be adjusted for in these tools is given in Appendix E.

##### 9.2. Distributional assumptions and outlying responses

Distributional assumptions (e.g. normality of continuous variables) will be assessed visually prior to analysis and outlying data will be queried; although the combination/transformation of the patient data used in each diagnostic tool in the *Algorithms for Index Test Combinations and Thresholds* section will be followed.

##### 9.3. Handling missing data

Results of all index tests/patient data and reference standard will be recorded. Patterns of missingness will be investigated in the premenopausal and postmenopausal groups by tabulating the predictors and reference standard results. This analysis will provide information on predictors that may be difficult to obtain in practice. If participants have missing data, then pairwise comparisons of the diagnostic tools will be made. As sensitivity analysis, multiple imputation by chained equations (MICE)10 will be used to impute missing values in order to avoid bias and make best use of the data, by replacing missing values with plausible values based on the distribution of the observed data. The number of imputed datasets that will be created will be determined by the percentage of participants that have at least one candidate predictor missing.

##### 9.4. Data manipulations

The Statistician will derive and manipulate all responses from the raw data recorded in the database according to the combination/transformation of patient data proposed for each diagnostic tool in the *Algorithms for Index Test Combinations and Thresholds* section.

##### 9.5. Analysis methods – primary outcome(s)

For the primary analysis, we will validate the diagnostic tools identified in Section 4.2 using the premenopausal and postmenopausal groups for diagnosing the target condition of primary invasive ovarian malignant neoplasms versus benign. We will report estimates of sensitivity, specificity, c-index (area under Receiver operating curve (ROC) curve), PPV and Negative Predictive value (NPV) and for risk prediction models a calibration plot.

A ROC plot will be created including the following models defined in Section 4.2: Trial tests with the thresholds labelled:

- RMI 1 (Thresholds: 200, 250)
- ROMA (Thresholds: 13.1% for premenopausal and 27.7% for postmenopausal, 12.5% for premenopausal and 14.4% for postmenopausal, 7.4% for premenopausal and 25.3% for postmenopausal, 11.4% for premenopausal and 29.9% for postmenopausal)
- UCL model
- ADNEX (Primary threshold: 10%, secondary threshold: 3%)
- IOTA sRisk model (Primary threshold: 10%, secondary threshold: 3%)
- CA125 (Thresholds: 87 IU/ml for premenopausal and 35 IU/ml for postmenopausal)

The performance of:

- RMI 1 (Thresholds: 200)
- ROMA (Thresholds: 13.1% for premenopausal and 27.7% for postmenopausal, 12.5% for premenopausal and 14.4% for postmenopausal, 7.4% for premenopausal and 25.3% for postmenopausal, 11.4% for premenopausal and 29.9% for postmenopausal)
- ADNEX (Primary threshold: 10%, secondary threshold: 3%)
- IOTA simple rules (see SAP Appendix C for more details)
- IOTA sRisk model (Primary threshold: 10%, secondary threshold: 3%)
- CA125 (Thresholds: 87 IU/ml for premenopausal and 35 IU/ml for postmenopausal)

will be compared to the existing RMI 1 model at a threshold of 250. The difference in sensitivity and specificity (and their corresponding 95% confidence intervals) will be assessed using McNemar’s test. The difference (including 95% CI) and p-value will be reported from this test.

The risk prediction models will produce a predicted risk of OC for all the individuals in our study. Therefore, we will compare the observed outcome from histology or at 12 months follow-up with this predicted risk. The calibration (in terms of calibration slope) and discrimination (e.g. c-index) will be evaluated for the models identified. Discrimination performance will be compared to the existing RMI 1 model.

For risk prediction models, the calibration will be shown visually by grouping women into deciles ordered by predicted risk and considering the agreement between the mean predicted risk and the observed events in each decile. The value for the calibration slope should ideally be one signifying perfect agreement between the predicted probabilities and the observed probabilities. A calibration slope < 1 indicates that a model over-predicts while a calibration slope > 1 indicates under prediction.

##### 9.6. Analysis methods – secondary outcomes

In a similar manner to the primary analysis, we will validate the accuracy of the diagnostic tools for the binary outcomes of:

- primary invasive and secondary ovarian malignant neoplasms versus benign (borderline will be considered with benign).
- primary invasive, secondary and borderline ovarian malignant neoplasms versus benign.
- primary invasive, secondary and borderline ovarian malignant neoplasms versus benign in the subset of patients that were assessed in one-stop clinics.
- primary invasive, secondary and borderline ovarian malignant neoplasms versus benign where ultrasound was performed by those who had passed IOTA Quality assurance.

##### 9.7. Analysis methods – exploratory outcomes and analyses

Exploratory analysis includes:

- Tabulation of the patients’ anxiety (anxiety questionnaire) at presentation and at 12 months for patients who had surgery.
- Tabulation of participants diagnosed with OC that had surgery, chemotherapy, surgery and chemotherapy or palliation by pre-menopausal and postmenopausal (Outcome CRF).
- Tabulation of participant outcome at 12 months by mode of presentation.
- Assess the predictive value of the symptoms from the symptoms questionnaire for predicting OC through logistic regression modelling. Odds ratios (95% CI) for each symptom to diagnose OC will be calculated.

Exploratory outcomes that will be tabulated will be presented using simple summary statistics by premenopausal and postmenopausal groups i.e. numbers and percentages for binary data and means (or medians) and standard deviations (or inter-quartile ranges) for continuous normal (or non-normal) data.

##### 9.8. Safety data

A table listing study related serious adverse events (SAEs), as defined in Section 6.1 of the protocol, will be provided according to their relationship of “Definite”, “Probable” and “Possible”. The number and percentage of participants experiencing any SAEs will be presented by test group (i.e. premenopausal or postmenopausal) and according to their severity level and seriousness of the event. For each patient, only maximum severity experienced for each type of adverse event will be displayed.

##### 9.9. Planned subgroup analyses

Subgroup analyses will be limited to primary outcome on all model external validations. The following will assess the difference in the C-statistic:

- between participants that have surgery versus participants that have biopsy. This will be investigated by creating an indicator variable for patients that either had surgery or biopsy and comparing/testing the area under the ROC curve between the two groups using the roccomp test in Stata.^11^
- between participants that have surgery and versus those that are followed up for 12 months participants. This will be investigated by creating an indicator variable (surgery or not) and comparing/testing the area under the ROC curve between the two groups.
- between high volume recruiting sites (County Durham & Darlington, West Hertfordshire (Watford General), Princess Anne (Southampton), University of Wales (Cardiff), East Surrey and Northampton) versus other organisations. This will be investigated by creating an indicator variable (high volume or not) and comparing/testing the area under the ROC curve between the two groups.
- between one-stop clinics (Southampton, Cardiff, Northampton, Surrey) versus other organisations. This will be investigated by creating an indicator variable (one-stop clinic or not) and comparing/testing the area under the ROC curve between the two groups.

Further subgroup analyses include:

- Is the time to diagnosis quicker in one stop clinics than other organisations in women referred through rapid access routes? The average number of days to diagnosis will be compared between one stop clinics and other organisations.
- Does the accuracy differ between participants who come through the rapid access route (found in the Registration form) versus other routes? This will be investigated by creating an indicator variable (rapid access or not) and comparing/testing the area under the ROC curve between the two groups.
- Does accuracy differ between those that pass the quality assurance check on IOTA rules versus those without a quality assurance check? This will be investigated on IOTA simple rules and ADNEX risk prediction models by creating an indicator variable (quality assurance pass or not) and comparing/testing the area under the ROC curve between the two groups.
- The accuracy of the diagnostic tools by the different routes into secondary care (routine referral from GP, two week wait referral from GP, presentation via A&E, referral from cancer unit at RMI 1>250 and referral from other speciality). This will be investigated by creating an indicator variable for the different routes and comparing/testing the area under the ROC curve between the groups.

#### 9.10. Sensitivity analyses

As sensitivity analysis, multiple imputation by chained equations (MICE)10 will be used to impute missing values in order to avoid bias and make best use of the data, by replacing missing values with plausible values based on the distribution of the observed data. The number of imputed datasets that will be created will be determined by the percentage of participants that have at least one candidate predictor missing.

#### 10. Analysis of sub-randomisations

N/A

#### 11. Health economic analysis

Resource usage for each of the diagnostics will be broken down and displayed along with their unit costs alongside the outcomes for each pathway. The resource usage will include the different types of tests administered, the number of inpatient and outpatient consultations, and any operative procedures undertaken. This approach will help to show which are the major cost drivers for each of the diagnostic pathways and will be collected as part of the clinical CRF.

#### 12. Statistical software

Statistical analysis will be undertaken in the following statistical software package: Stata version 17.

#### SAP Appendix A: Deviations from SAP

N/A

#### SAP Appendix B: Trial schema

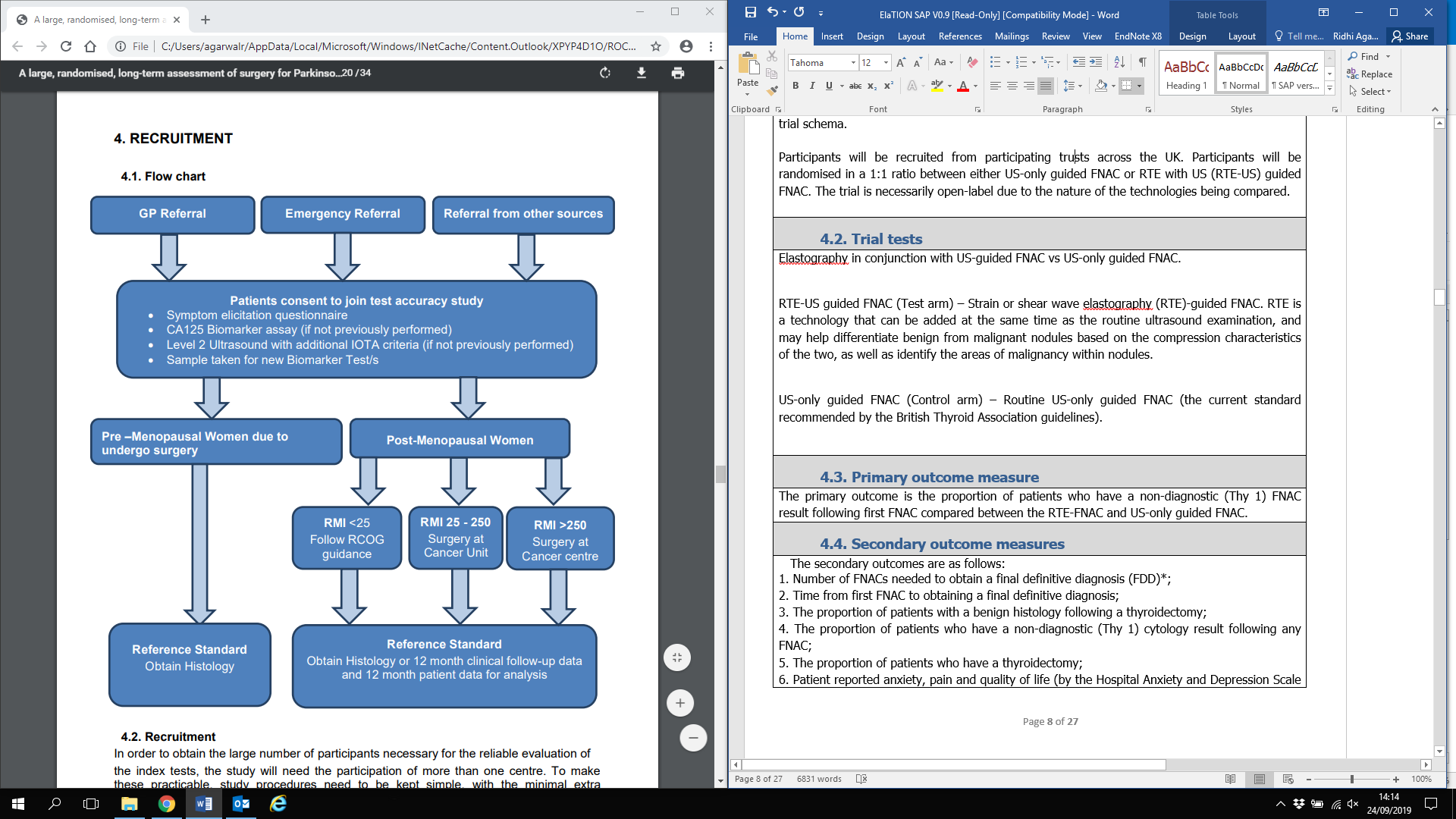

#### SAP Appendix C: IOTA simple rules

IOTA group have established simple rules based on a number of clearly defined USG features that can guide examiners. Using these simple rules no risk estimates are produced, but tumours are classified as benign, malignant or unclassifiable. The simple rules consist of five USG features of malignancy (M-features) and five USG features suggestive of a benign mass (B-features). These features with corresponding USG images are shown in this image. A mass is classified as malignant if at least one M-feature and none of the B-features are present, and vice versa. If no B- or M-features are present, or if both B- and M-features are present, then the rules are considered inconclusive (unclassifiable mass) and a different diagnostic method should be used.

#### SAP Appendix D: Schedule of assessments

See study protocol 2.14: Schedule of Events & CRF completion.

#### SAP Appendix E: List of covariates

Details of the combination of patient data in the diagnostics tools is provided in the *Algorithms for Index Test Combinations and Thresholds* section.
